# Subtyping and staging of Alzheimer’s disease from routine structural MRI with PHASE–AD

**DOI:** 10.64898/2026.06.26.26356678

**Authors:** Hannah Baumeister, Falk Lüsebrink, Luca Kleineidam, Niels Hansen, Matthias Schmid, Alexis Moscoso, Antoine Leuzy, Sophie E. Mastenbroek, Colin Groot, Frederic Brosseron, Alfredo Ramirez, Lukas Preis, Daria Gref, Eike J. Spruth, Maria Gemenetzi, Slawek Altenstein, Klaus Fliessbach, Okka Kimmich, Björn H. Schott, Ayda Rostamzadeh, Wenzel Glanz, Enise I. Incesoy, Michaela Butryn, Daniel Janowitz, Boris-Stephan Rauchmann, Mihovil Mladinov, Alice Grazia, Sebastian Sodenkamp, Tony Stöcker, Stefan Hetzer, Peter Dechent, Sophia Stoecklein, the Alzheimer’s Disease Repository Without Borders Investigators, the Alzheimer’s Disease Neuroimaging Initiative, the DELCODE study group, Oliver Peters, Julian Hellmann-Regen, Josef Priller, Anja Schneider, Jens Wiltfang, Katharina Buerger, Robert Perneczky, Stefan Teipel, Christoph Laske, Annika Spottke, Michael Wagner, Frank Jessen, Emrah Düzel, David Berron

## Abstract

Structural MRI is routinely acquired in the clinical assessment of Alzheimer’s disease, yet quantitative morphometric indices derived from these scans remain largely confined to research settings. Here we present PHASE–AD — a framework that translates such indices into clinically interpretable classifications of atrophy subtype and stage that jointly capture atrophy progression while accounting for inter-individual atrophy heterogeneity. PHASE–AD is trained on MRI scans from 8,415 participants and robustly captures limbic-predominant and hippocampal-sparing atrophy subtypes that were identified across seven independent datasets. Two cross-validation schemes revealed high robustness across different field strength and scanner manufacturer configurations. Atrophy classifications were associated with diverging clinical profiles and tau accumulation patterns. In prospective designs mirroring contemporary AD trials, they stratified longitudinal cognitive trajectories and outperformed semi-quantitative visual MRI assessments as a clinically established comparator. These findings support the integration of automated atrophy subtyping and staging into clinical practice and pharmacological trials.

## Introduction

Structural magnetic resonance imaging (MRI) is a key neuroimaging modality used in the clinical assessment of Alzheimer’s disease (AD), which is the leading cause of dementia.^1,2^ Beyond its central use in differential diagnosis and, more recently, in safety monitoring for amyloid-targeting treatments (ATTs, ref.^3^), structural MRI enables the *in vivo* assessment of regional neurodegeneration through spatial patterns of brain atrophy.^4^ In routine clinical practice, brain atrophy is most commonly evaluated through qualitative visual inspection or semi-quantitative rating scales, such as the medial temporal atrophy scale (MTA, ref.^5^).^6,7^ Meanwhile, an extensive body of research supports the use of quantitative indices of brain morphometry, including regional volumetric or cortical thickness estimates, for improved diagnostic sensitivity and prognostic accuracy relative to these reader-dependent, ordinal assessment tools.^8–11^ Despite this evidence, the integration of quantitative brain morphometry in clinical workflows has remained limited; in a European survey of neuroimaging centers, only a small proportion of clinicians reported routine use of such measures in their assessments.^7^

Various factors likely contribute to this translational gap, and the current study addresses three that we consider among the most critical. First, the computational demands and long processing times of established neuroimaging pipelines are likely incompatible with time-constrained clinical workflows. As an example, FreeSurfer — one of the most widely used tools for deriving morphometric indices from MRI — requires several hours of processing time per scan.^12^ Second, integrating morphometric estimates across a large number of cortical and subcortical regions of interest (ROIs) presents a substantial interpretive burden that may exceed what is feasible in routine clinical practice. Finally, and most fundamentally, the lack of validated norms severely limits the interpretability of individual morphometric estimates. Establishing robust norms is methodologically challenging, as morphometric estimates are systematically influenced by acquisition-related factors, including scanner manufacturer and magnetic field strength, as well as participant variables, such as age, sex, and head size.^13^ The combined effect of these factors typically requires very large reference samples to reliably characterize normative variation. While previous efforts have been made to establish normative reference frameworks for brain morphometry from large-scale datasets, most do not generalize to unseen sites lacking local samples of healthy controls from which to derive site-specific normative values, as is typical of small radiological centers and clinics.^14–17^

For these reasons, a disease-specific composite measure of brain structure relative to a robust population norm, rapidly deliverable from a single structural MRI scan, would be of substantial clinical value. Crucially, such an approach must account for the fact that atrophy in AD does not progress uniformly across individuals. Both hypothesis- and data-driven modeling frameworks have converged on considerable inter-individual heterogeneity in the spatiotemporal patterns of brain atrophy in AD.^18^ Distinct atrophy subtypes have been observed not only across clinical variants of the disease, such as between typical amnestic and atypical non-amnestic presentations, but also within the amnestic phenotype itself.^19–25^ Despite growing evidence for divergent atrophy cascades in AD, no universal framework exists for classifying individuals along these trajectories from a single out-of-sample MRI acquisition while demonstrating utility in relevant applied settings.

While the exact nature of brain atrophy heterogeneity in AD remains unclear and prior findings appear highly method-dependent, a previous review of the literature proposed that the primary axis of variation runs between limbic-predominant and hippocampal-sparing extremes.^18^ In line with this prior model, earlier work from our group identified distinct limbic-predominant and hippocampal-sparing atrophy subtypes in two memory clinic-based AD cohorts.^19^ The underlying model was built on the Subtype and Stage Inference (SuStaIn, ref.^23^) algorithm and jointly infers distinct atrophy cascades represented in the provided training set, as well as each individual’s position within these cascades, yielding per-scan classifications of both atrophy subtype and stage. In the present study, we seek to extend this approach into PHASE–AD (Progression and Heterogeneity Analysis from Structural MRI in Early AD) — a generalizable atrophy subtyping and staging framework directly applicable to the raw structural MRI data routinely acquired in clinical dementia care, research settings, and pharmacological trials. To this end, we evaluate the replicability of our prior findings across seven independent and diverse samples using an updated methodology with reduced processing demands and model requirements. Moreover, we aim to establish PHASE–AD as a universal subtyping and staging model that both reproduces these sample-specific results and robustly classifies unseen data. We further formally characterize the demographic, cognitive, and clinical correlates of PHASE–AD atrophy subtype and stage classifications through meta-analyses across samples. Finally, we evaluate the translational value of PHASE–AD for two application contexts of current relevance. First, we assess the utility of atrophy subtype and stage for estimating regional tau burden. Tau PET — the only modality offering spatially resolved information on tau burden *in vivo* — is costly and largely inaccessible outside of specialized research and tertiary care settings. PET-free proxies of tau burden are of direct clinical relevance in the context of ATTs, as trial data for donanemab have shown the largest treatment effects in individuals with low-to-intermediate relative to high tau burden.^26,27^ Second, we investigate the prognostic information provided by PHASE–AD. Accurate prognosis of an individual’s clinical trajectory is not only crucial to personalizing clinical management, but also central to trial design strategies aimed at increasing statistical power through sample homogenization, enrichment, and stratification. Prognostic utility is therefore evaluated under two scenarios selected in reference to recently completed phase 3 ATT trials. The first scenario is defined in a sample of individuals with mild symptomatic AD using the Clinical Dementia Rating Sum of Boxes (CDR–SB, ref.^28,29^) as the outcome over 180 weeks (cf., the CLARITY AD trial of lecanemab; ref.^30^). The second is defined in a sample of individuals with preclinical AD, using the Preclinical Alzheimer’s Cognitive Composite 5 (PACC–5, ref.^31^) as the outcome over 240 weeks (cf., the Anti-Amyloid Treatment in Asymptomatic Alzheimer’s Study [A4] trial of solanezumab; ref.^32^). In the mild symptomatic AD scenario, the prognostic utility of PHASE–AD classifications was additionally benchmarked against MTA scores as a clinically established measure of AD-related atrophy. Given the increasing clinical availability of plasma p-tau^217^ as an accessible and accurate biomarker of AD pathology, we additionally examined whether the utility of PHASE–AD for both applications is preserved independently of this marker.^33^

## Methods

### Participants

We analyzed data from 8,415 participants with available T_1_-weighted MRI scans acquired at 1.5T or 3T from five international cohort studies. Given the established influence of MRI field strength on morphometric atrophy markers, we stratified the dataset by cohort study and field strength.^13^ This included the following samples using 3T MRI: the DZNE Longitudinal Cognitive Impairment and Dementia study (DELCODE-3T, *N* = 911; ref.^34^), the Alzheimer’s Disease Neuroimaging Initiative (ADNI-3T, *N* = 1,052, phases GO, 2, and 3), the National Alzheimer’s Coordinating Center Uniform Data Set (NACC-3T, *N* = 3,801), and the A4 study, combined with its Longitudinal Evaluation of Amyloid Risk and Neurodegeneration Extension (A4/LEARN-3T, *N* = 1,237, all included MRI scans acquired prior to treatment initiation). In addition, the following samples using 1.5T MRI were included: NACC-1.5T (*N* = 657), ADNI-1.5T (*N* = 606, phase 1), and the Alzheimer’s Disease Repository Without Borders (ARWIBO-1.5T, *N* = 151).

The cohort-specific inclusion criteria are reported in the Supplementary Methods. In addition, inclusion was limited to participants who were at least 60 years old at the time of MRI and either cognitively unimpaired (CU) or diagnosed with mild cognitive impairment (MCI) or dementia of the Alzheimer type (DAT) within one year of the scan. If a participant met these criteria at multiple MRI time points, we selected the earliest eligible scan.

#### Sub-samples

The full dataset was partitioned into several sub-samples for different analyses. First, biological AD samples were defined for each cohort as Aβ-positive participants, supplemented by Aβ-negative CU participants as a reference for atrophy marker normalization. These samples were used to evaluate atrophy cascades independently of cognitive impairment not attributable to AD pathology. Second, cohort-wise Aβ-positive samples were used to assess whether the meta-analytically derived clinical profiles associated with PHASE– AD classifications could be recovered in biologically defined AD. Third, a tau PET sample comprising participants with available tau PET and a nested sample additionally requiring plasma p-tau_217_ was used to examine tau burden along the identified atrophy cascades. Within the tau PET sub-sample, an ATT-eligible tau PET sample was further defined, approximating ATT eligibility criteria (MCI or DAT diagnosis, MMSE ≥ 19, no APOE ε4 homozygosity; ref.^35^). Finally, two longitudinal samples were defined for prognostic analyses, each restricted to Aβ-positive participants with at least two assessments of the outcome of interest (see section **Neuropsychological assessment**). The preclinical AD longitudinal sample comprised participants from A4 (LEARN participants were excluded due to the Aβ-positivity restriction). The mild symptomatic AD longitudinal sample comprised Aβ-positive participants from ADNI diagnosed with MCI or DAT and an MMSE ≥ 19 to ensure dementia severity was mild. A nested sample of the mild symptomatic AD longitudinal sample with available MTA scores was additionally considered for comparison with visual atrophy ratings.

### Alzheimer’s disease biomarkers

#### Amyloid-β status

Biomarkers of Aβ pathology were available for a subset of participants (*n* = 3,887; 46.19%). Aβ status was defined using PET or CSF measures, depending on cohort and data availability (Supplementary Methods). Aβ biomarkers were considered only if acquired within one year of the included MRI scan. Data on Aβ status were omitted from the NACC-1.5T and ARWIBO-1.5T samples because sample sizes were insufficient for statistical inference.

#### Tau PET

Tau PET was used to assess regional tau burden *in vivo* and was available in A4/LEARN-3T (*n* = 229; all using ^18^F-Flortaucipir), ADNI-1.5T/-3T (*n* = 177; using ^18^F-Flortaucipir), and NACC-3T (*n* = 54; *n* = 17 using ^18^F-Flortaucipir, *n* = 27 using ^18^F-MK-6240, *n* = 10 using ^18^F-PI-2620). Tau PET data were included if acquired within one year of the corresponding MRI scan or within one year of another MRI scan during which participants met this study’s inclusion criteria. In addition, tau PET data were only included if Aβ-positivity was established within one year of the scan. Among these 460 participants, 61 participants lacked a tau PET scan within one year of their index MRI and a second MRI acquisition temporally proximate to an available tau-PET scan was used instead (mean ± *SD* interval between MRI acquisitions: 4.93 ± 2.97 years). Regional tau burden was harmonized across tracers onto the CenTauR scale as previously described.^36^ SUVR images were generated using the inferior cerebellar cortex as the reference region, and tracer-specific SUVRs were converted to CenTauR units using the joint propagation model mapping equations, which place 0 at the mean of young Aβ-negative CU individuals and 100 at the mean of typical DAT patients without requiring a reference tracer. CenTauR values were derived for the universal (global cortical) CenTauR region and for four predefined subregions (mesial temporal, meta-temporal, temporo-parietal, and frontal) defined within the universal mask. Tau burden was additionally stratified into low, medium, and high categories using universal ROI CenTauR cut-points of 16.1 and 68.7, corresponding to thresholds established in the TRAILBLAZER-ALZ clinical trial program (Moscoso, Leuzy, Raket, et al., *in revision*).^27^

#### Plasma p-tau_217_

Plasma p-tau_217_ biomarkers acquired within one year of an included MRI scan were available in ADNI-1.5T (*n* = 105, 17.33%), ADNI-3T (n = 297, 28.23%), and A4/LEARN-3T (n = 601, 48.59%). In ADNI samples, ratios of p-tau_217_/Aβ_42_ were established using chemiluminescent immunoassays (RUO IVD kits) on the Fujirebio Lumipulse G1200 platform. In A4/LEARN-3T, the concentration of p-tau_217_ was established using an electrochemiluminescence (ECL) immunoassay on the Meso Scale Discovery (MSD) Sector S 600 MM platform at the Lilly Clinical Diagnostics Laboratory.

### Neuropsychological assessment

For the cross-sectional meta-analytic evaluation of clinical and cognitive correlates of PHASE–AD classifications, neuropsychological instruments capturing distinct cognitive constructs were selected. Global cognition was measured with the Mini-Mental State Examination (MMSE, ref.^37^). Sum scores from the Functional Activities Questionnaire (FAQ, ref.^38^) were used to assess instrumental activities of daily living. Episodic memory was evaluated using the Logical Memory Delayed Recall subtest from the Wechsler Memory Scale-Revised (ref.^39^; version IIA in ADNI and NACC; the A4/LEARN protocol alternated among three versions across study visits) or the Wechsler Memory Scale-IV (ref.^40^; version IIB in DELCODE). As NACC discontinued the collection of MMSE and Logical Memory Delayed Recall scores in March 2015, we converted available Montreal Cognitive Assessment (ref.^41^) scores to MMSE scores and Craft Story 21 Delayed Recall scores to WMS-R Logical Memory Delayed Recall scores using published crosswalk tables.^42^ The difference in completion time between Trail-Making Test parts B and A (TMT B–A) was used as a measure of executive functioning.^43^ Negative TMT B–A scores were omitted. TMT B–A and FAQ scores were inverted so that higher scores corresponded to better performance across instruments. Only neuropsychological data acquired within one year of the included MRI scan were used. Details on neuropsychological data availability for each sample as well as score standardization and normalization are provided in the Supplementary Methods. Prognostic analyses were conducted using longitudinal CDR–SB scores in the ADNI-based mild symptomatic AD longitudinal sample and PACC–5 scores in the A4-based preclinical AD longitudinal sample. Participants were included if at least two observations fell within the pre-specified follow-up windows of 180 weeks post-MRI for CDR–SB and 240 weeks post-MRI for PACC–5.

### MRI selection and processing

All used T_1_-weighted MRI scans were acquired on MRI systems manufactured by Philips, Siemens, or General Electric (GE). To ensure robust normative modeling, we required at least 15 scans per scanner manufacturer within each sample, leading to the exclusion of scans acquired on Philips and Siemens systems in NACC-1.5T and on Philips and Siemens systems in ARWIBO-1.5T.

Raw images were processed using FastSurfer (v2.2.0), a faster, deep learning-based alternative to FreeSurfer.^44^ The exact difference in processing time between FreeSurfer, which was used in our original atrophy subtyping and staging model (ref.^19^), and FastSurfer depends on image characteristics and available computational resources. Henschel *et al*. report a mean runtime reduction of approximately 78% compared with FreeSurfer (54 *versus* 244 min.) when both methods are executed with parallelized segmentation and surface reconstruction (four threads) for both hemispheres.^44^ Desikan-Killiany labels were collapsed into bilateral meta-regions, including the medial temporal lobe (MTL) as well as the temporal, parietal, frontal, and occipital lobes (see Supplementary Table 1). Gray matter volume served as the atrophy marker in the MTL, whereas average cortical thickness was used for other regions.

**Table 1.** Core participant characteristics, stratified by sample.

|  | A4/LEARN-3T | ADNI-3T | DELCODE-3T | NACC-3T | ADNI-1.5T | ARWIBO-1.5T | NACC-1.5T | Whole sample |
| --- | --- | --- | --- | --- | --- | --- | --- | --- |
| *N* | 1,237 | 1,052 | 911 | 3,801 | 606 | 151 | 657 | 8,415 |
| **Treatment** |  |  |  |  |  |  |  |  |
| Placebo | 352 (28.5%) | 0 (0.0%) | 0 (0.0%) | 0 (0.0%) | 0 (0.0%) | 0 (0.0%) | 0 (0.0%) | 352 (4.2%) |
| Solanezumab | 353 (28.5%) | 0 (0.0%) | 0 (0.0%) | 0 (0.0%) | 0 (0.0%) | 0 (0.0%) | 0 (0.0%) | 353 (4.2%) |
| None | 532 (43.0%) | 1,052 (100.0%) | 911 (100.0%) | 3,801 (100.0%) | 606 (100.0%) | 151 (100.0%) | 657 (100.0%) | 7,710 (91.6%) |
| **Diagnosis<sup>a</sup>** |  |  |  |  |  |  |  |  |
| CU | 1,237 (100.0%) | 445 (42.3%) | 654 (71.8%) | 2,595 (68.3%) | 182 (30.0%) | 77 (51.0%) | 323 (49.2%) | 5,513 (65.5%) |
| MCI | 0 (0.0%) | 459 (43.6%) | 154 (16.9%) | 725 (19.1%) | 278 (45.9%) | 35 (23.2%) | 123 (18.7%) | 1,774 (21.1%) |
| DAT | 0 (0.0%) | 148 (14.1%) | 103 (11.3%) | 481 (12.7%) | 146 (24.1%) | 39 (25.8%) | 211 (32.1%) | 1,128 (13.4%) |
| **Age at MRI, years** | 71.5 (4.6) | 72.8 (6.9) | 71.3 (6.1) | 72.6 (7.1) | 76.0 (6.1) | 70.4 (6.5) | 76.9 (7.4) | 72.9 (6.8) |
| **Sex, female** | 481 (38.9%) | 510 (48.5%) | 465 (51.0%) | 2,362 (62.1%) | 241 (39.7%) | 81 (53.6%) | 381 (58.0%) | 4,521 (53.7%) |
| **Race** |  |  |  |  |  |  |  |  |
| Asian | 25 (2.0%) | 36 (3.4%) | 0 (0.0%) | 74 (1.9%) | 13 (2.1%) | 0 (0.0%) | 27 (4.1%) | 175 (2.1%) |
| Black or African American | 31 (2.5%) | 76 (7.2%) | 0 (0.0%) | 794 (20.9%) | 33 (5.4%) | 0 (0.0%) | 80 (12.2%) | 1,014 (12.0%) |
| White | 1,160 (93.8%) | 912 (86.7%) | 0 (0.0%) | 2,834 (74.6%) | 552 (91.1%) | 151 (100.0%) | 543 (82.6%) | 6,152 (73.1%) |
| Other | 6 (0.5%) | 19 (1.8%) | 0 (0.0%) | 74 (1.9%) | 3 (0.5%) | 0 (0.0%) | 3 (0.5%) | 105 (1.2%) |
| Unknown | 15 (1.2%) | 9 (0.9%) | 911 (100.0%) | 25 (0.7%) | 5 (0.8%) | 0 (0.0%) | 4 (0.6%) | 969 (11.5%) |
| **Ethnicity** |  |  |  |  |  |  |  |  |
| Hispanic or Latino | 41 (3.3%) | 61 (5.8%) | 0 (0.0%) | 268 (7.1%) | 16 (2.6%) | 0 (0.0%) | 178 (27.1%) | 564 (6.7%) |
| Not Hispanic or Latino | 1,185 (95.8%) | 985 (93.6%) | 0 (0.0%) | 3,521 (92.6%) | 585 (96.5%) | 151 (100.0%) | 476 (72.5%) | 6,903 (82.0%) |
| Unknown | 11 (0.9%) | 6 (0.6%) | 911 (100.0%) | 12 (0.3%) | 5 (0.8%) | 0 (0.0%) | 3 (0.5%) | 948 (11.3%) |
| **Education, years** | 16.6 (2.7) | 16.4 (2.6) | 14.4 (3.0) | 16.0 (2.6) | 15.5 (3.1) | 10.7 (4.7) | 14.9 (3.0) | 15.8 (2.9) |
| Missing | 0 (0.0%) | 50 (4.8%) | 0 (0.0%) | 465 (12.2%) | 5 (0.8%) | 70 (46.4%) | 135 (20.5%) | 725 (8.6%) |
| **Scanner manufacturer** |  |  |  |  |  |  |  |  |
| GE | 255 (20.6%) | 250 (23.8%) | 0 (0.0%) | 897 (23.6%) | 271 (44.7%) | 151 (100.0%) | 657 (100.0%) | 2,481 (29.5%) |
| Philips | 133 (10.8%) | 179 (17.0%) | 0 (0.0%) | 297 (7.8%) | 81 (13.4%) | 0 (0.0%) | 0 (0.0%) | 690 (8.2%) |
| Siemens | 849 (68.6%) | 623 (59.2%) | 911 (100.0%) | 2,607 (68.6%) | 254 (41.9%) | 0 (0.0%) | 0 (0.0%) | 5,244 (62.3%) |
| **APOE ε4 status, carrier** | 532 (43.0%) | 433 (44.1%) | 319 (35.0%) | 1,200 (39.8%) | 314 (51.8%) | 4 (14.3%) | 283 (44.8%) | 3,085 (41.6%) |
| Missing | 0 (0.0%) | 71 (6.7%) | 0 (0.0%) | 783 (20.6%) | 0 (0.0%) | 123 (81.5%) | 25 (3.8%) | 1,002 (11.9%) |
| **Aβ status, positive<sup>b</sup>** | 705 (57.0%) | 450 (47.0%) | 280 (38.3%) | 314 (46.8%) | 187 (64.7%) | NA | NA | 1,936 (49.8%) |
| Missing | 0 (0.0%) | 94 (8.9%) | 179 (19.6%) | 3130 (82.3%) | 317 (52.3%) | NA | NA | 4,528 (53.8%) |
| **Aβ status source<sup>c</sup>** |  |  |  |  |  |  |  |  |
| Probabilistic modeling | 0 (0.0%) | 0 (0.0%) | 298 (40.7%) | 0 (0.0%) | 0 (0.0%) | NA | NA | 298 (7.7%) |
| CSF Aβ<sub>42</sub>/Aβ<sub>40</sub> | 0 (0.0%) | 0 (0.0%) | 434 (59.3%) | 0 (0.0%) | 0 (0.0%) | NA | NA | 434 (11.2%) |
| CSF p-tau181/ Aβ<sub>42</sub> | 0 (0.0%) | 352 (36.7%) | 0 (0.0%) | 0 (0.0%) | 288 (99.7%) | NA | NA | 640 (16.5%) |
| <sup>18</sup>F-Florbetaben PET | 0 (0.0%) | 133 (13.9%) | 0 (0.0%) | 279 (41.6%) | 0 (0.0%) | NA | NA | 412 (10.6%) |
| <sup>18</sup>F-Florbetapir PET | 1237 (100.0%) | 473 (49.4%) | 0 (0.0%) | 78 (11.6%) | 1 (0.3%) | NA | NA | 1,789 (46.0%) |
| <sup>18</sup>F-NAV4694 PET | 0 (0.0%) | 0 (0.0%) | 0 (0.0%) | 39 (5.8%) | 0 (0.0%) | NA | NA | 39 (1.0%) |
| <sup>11</sup>C-Pittsburgh Compound B PET | 0 (0.0%) | 0 (0.0%) | 0 (0.0%) | 275 (41.0%) | 0 (0.0%) | NA | NA | 275 (7.1%) |

Reported are mean (*SD*) for continuous variables and counts (%) for categorical variables. <sup>a</sup>Diagnosis closest in time to the MRI scan was selected. Participants were included only if both an eligible MRI scan and a diagnosis recorded within one year of the scan were available. <sup>b</sup>Aβ status was considered only if recorded within one year of the MRI scan. When multiple records of diagnosis or Aβ status met these criteria, those closest in time to the MRI scan were used. <sup>c</sup>Sources of Aβ status are reported only for the sub-samples with available Aβ biomarker data. Abbreviations: NA, not applicable.

### Medial temporal atrophy scale

A subset of MRI scans acquired in ADNI were visually assessed by an accredited rater (S.E.M. or C.G.; inter-rater agreement κ = 0.68, 95% CI [0.40–0.97], ref.^45^) using the MTA rating scale. Scores were averaged across hemispheres and rounded up to the next highest MTA score.

### Atrophy subtyping and staging

The subtyping and staging workflow consisted of three main steps: (i) atrophy marker normalization, (ii) reconstruction of atrophy progression sequences, and (iii) cross-validation. Each step is described below. This workflow was first applied to individual samples to evaluate the reproducibility of atrophy progression patterns across diverse datasets (within-sample modeling). To ensure that results were not driven by participants with cognitive decline due to non-AD etiology, within-sample modeling was repeated in biological AD samples. Subsequently, all participants were pooled into a combined dataset to derive a global subtyping and staging model (PHASE–AD).

#### Marker normalization

MTL volumes were corrected for total intracranial volume (Supplementary Methods). Linear regression models were fitted as normative models of atrophy in all CU participants, with age, sex, scanner manufacturer, and field strength as predictors. For each atrophy marker, multiple model specifications differing in their treatment of non-linear age effects and predictor interactions were compared, selecting the best-fitting formula based on the Akaike Information Criterion (AIC; see Supplementary Methods). This model selection step was performed once, resulting in a fixed formula for each atrophy marker. However, to prevent data leakage, model parameters were re-estimated within the relevant reference group at each modeling iteration. From the fitted normative models, *w*-scores were derived for each atrophy marker, representing covariate-adjusted *SD* differences from the reference group mean. All normalized atrophy measures were screened for numerical outliers (Supplementary Methods).

#### Reconstruction of atrophy sequences

The *w*-scored atrophy markers were next passed to SuStaIn (ref.^23^, using python v3.9.7), modeling up to five atrophy progression sequences (i.e., atrophy subtypes). The exact model specifications have been described previously by our group (ref.^19^), though we now consider three instead of two atrophy thresholds (*w* ∈ {–1, –2, –3}; *w* = –3 was not included in the original model) for improved granularity in more advanced disease stages. Consequently, SuStaIn estimated the most likely sequence(s) of 15 atrophy events. We refer to the positions of these events within the reconstructed sequence as atrophy stages. Individuals at atrophy stage 0 in any subtype were collapsed to form an atrophy-negative group. However, like participants at stages > 0, they were assigned probabilities of belonging to each subtype, which we incorporated into our downstream analyses. Though continuous modeling occurred across atrophy stages, their scaling does not imply equidistant time intervals.

#### Cross-validation

Each SuStaIn model was evaluated using stratified ten-fold cross-validation to assess robustness and to determine the optimal number of atrophy subtypes using previously described cross-validation information criterion (CVIC) criteria.^23^ Participants were randomly assigned to folds while preserving the proportions of the reference group, scanner manufacturers, and MRI field strength (where applicable). Leave-one-sample-out (LOSO) cross-validation of the PHASE–AD model was used to estimate generalizability to independent samples and robustness to sample exclusion.

### Statistical analyses

All statistical analyses were conducted in R v4.2.2 using RStudio.^46,47^

#### Evaluation of atrophy classification models

Replicability of atrophy subtyping and staging models was assessed using two-way random-effects intraclass correlation coefficients (*ICC*_2_,_1_) for continuous measures (*w*-scores, subtype probabilities, atrophy stages) and Cohen’s κ for discrete subtype assignments. Similarity of atrophy progression sequences was quantified using the mean and *SD* of pairwise Wasserstein distances (*WD*) between posterior positional distributions of corresponding marker events. This measure reflects the stage shift required for sequence alignment, with lower values indicating greater sequence similarity. These metrics were applied across replicability analyses (full samples *versus* biological AD samples), within-sample *versus* PHASE–AD model comparisons, and cross-validation analyses. Subtype agreement metrics were reported separately for participants at atrophy stage 0 and atrophy stage ≥1, given that subtype assignment is expected to be less reliable in the absence of observed atrophy events.

#### Clinical associations of PHASE–AD classifications

Cross-sectional demographic and clinical associations of PHASE–AD classifications were examined within each individual sample and, where available, their Aβ-positive sub-samples. Subtype effects (including atrophy-negative individuals as a separate group) were assessed via Tukey-adjusted pairwise estimated marginal mean contrasts. Stage effects were modeled separately within each subtype. Associations with continuous outcomes (age, education, neuropsychological test scores) were estimated using linear regression (Cohen’s *d*), and stage effects were summarized as Fisher *z-*transformed partial correlation coefficients. Effects on binary outcomes (sex, APOE ε4, Aβ status, diagnosis) were modeled using Odds ratios (*OR*s) from bias-reduced logistic regression (*brglm2* R package, ref.^48^). All models adjusted for age, sex, and diagnostic group unless these were the outcome, years of education when predicting neuropsychological test scores, as well as treatment arm where applicable. Sample-level estimates were pooled via REML meta-analysis (*metafor* R package, ref.^49^). Between-sample heterogeneity was quantified using I_2_. Correction for false discovery rate (FDR) used the Benjamini-Hochberg procedure applied within prespecified families stratified by analysis type (subtype *versus* stage) and sample (full *versus* Aβ-positive).

#### Associations of PHASE–AD classifications with tau burden

To examine the relationship between PHASE–AD classifications and regional tau burden, linear regression models predicting regional CenTauR scores as a function of atrophy subtype, stage, and their interaction were fitted, adjusting for the PET–MRI time interval. For each ROI, a linear and a natural spline (internal knot at stage 3) specification were compared, selecting the best-fitting model by AIC. To quantify the unique variance in tau burden explained by atrophy subtype and stage beyond demographic and clinical covariates (age, sex, APOE ε4 carriership, diagnosis), a full model including both covariate and PHASE–AD classification terms was compared against a covariate-only reference model. Model improvement was assessed via differences in AIC (ΔAIC; threshold for model superiority: ≤ −2, ref.^50^) and *R*_2_ (Δ*R*_2_). In subsets with available plasma p-tau_217_ biomarkers, analyses were repeated cohort-wise with p-tau_217_ biomarkers included as an additional covariate. Ordinal logistic regression (cumulative link model with logit link) was used to model tau burden group as a function of atrophy subtype, stage, and their interaction. Covariate adjustment as well as model selection and comparisons were performed as described above.

#### Prognostic value of atrophy subtype and stage

The prognostic properties of PHASE–AD classifications were evaluated using linear mixed-effects models in the preclinical and mild symptomatic AD longitudinal samples using their respective outcomes of interest. All models covaried for age at MRI, sex, and APOE ε4 carrier status interacting with time. Models of PACC–5 additionally included test version and treatment arm. Models of CDR–SB included baseline diagnosis. Atrophy subtype, stage, and time were modeled as a three-way interaction. To identify the optimal functional form of time and atrophy stage effects, four variants per model specification were compared, crossing linear and natural spline parameterisations of time (internal knot at midpoint of the maximum follow-up window) and atrophy stage (internal knot at stage 3). The lowest-AIC variant was selected per specification. The incremental predictive value of models including PHASE–AD classifications as predictors over covariates-only reference models was assessed via 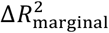 and ΔAIC, which were estimated under maximum likelihood (ML) to enable comparison across models with differing fixed effects. Models were fit under restricted maximum likelihood (REML) for parameter estimation and inference. In the mild symptomatic AD longitudinal sample, PHASE–AD classifications were additionally compared against MTA scores as a clinically established benchmark for visual atrophy rating, using the same model fit indices.

## Results

Sample characteristics are summarized in Table 1. An overview of the selected model formulae used for marker normalization is provided in Supplementary Figure 3.

### Limbic-predominant and hippocampal-sparing atrophy subtypes are identified across independent samples

Within-sample modeling using the original discovery cohort (i.e., DELCODE-3T; *n* = 793 overlapping between the current and original sample) replicated the previously published atrophy progression sequences using the updated methodology introduced in this study. The individual *w*-scored atrophy markers showed excellent overlap (mean ± *SD ICC*_2,1_ = 0.93 ± 0.01, all *p* < .001), enabling the recovery of the previously reported limbic-predominant and hippocampal-sparing atrophy subtypes (Supplementary Figure 4A–C). Regarding individual classifications, agreement for the probability of limbic-predominant atrophy (*P*_LP_; the inverted probability of hippocampal-sparing atrophy) was good (*ICC*_2,1_ = 0.83, *p* < .001 for stage > 0 classifications; *ICC*_2,1_ = 0.75, *p* < .001 otherwise). Subtype assignments showed fair to good agreement (κ = 0.70, *p* < .001 for stage > 0 classifications; otherwise κ = 0.54, *p* < .001; Supplementary Figure 4 D–F).

*De novo* within-sample modeling consistently identified limbic-predominant and hippocampal-sparing atrophy subtypes across nearly all samples. The only exception was ARWIBO-1.5T, where the two-subtype model yielded highly similar subtypes, but did not outperform the one-subtype solution (Supplementary Figures 5,6). For consistency with the other samples, the two-subtype model was nevertheless evaluated in ARWIBO-1.5T, although it was not clearly the best-fitting model. The atrophy sequences reconstructed within each sample are shown in Figure 1A. Subtype proportions across samples are presented in Figure 1C. Similarity among the identified atrophy sequences was high overall and comparable across subtypes, with a mean stage shift (*WD*) of 1.54 (*SD* = 1.25) for the hippocampal-sparing subtype and 1.81 (*SD* = 2.47) for the limbic-predominant subtype (Figure 1B). The higher mean and greater variability for the limbic-predominant subtype were driven by the sequence identified in A4/LEARN-3T (mean *WD* = 4.09; all other samples ≤ 1.56), likely reflecting the limited representation of advanced atrophy in this CU sample. As 95% of participants assigned to the limbic-predominant subtype were classified below atrophy stage 6, the later sequence events were estimated from very few cases. Within-sample modeling in the biological AD samples yielded limbic-predominant and hippocampal-sparing atrophy sequences that closely matched those identified in the full samples (mean ± *SD WD*_limbic-predominant_ = 0.33 ± 0.47; mean ± *SD WD*_hippocampal-sparing_ = 0.98 ± 0.87; Supplementary Figures 7,8).

**Figure 1.**
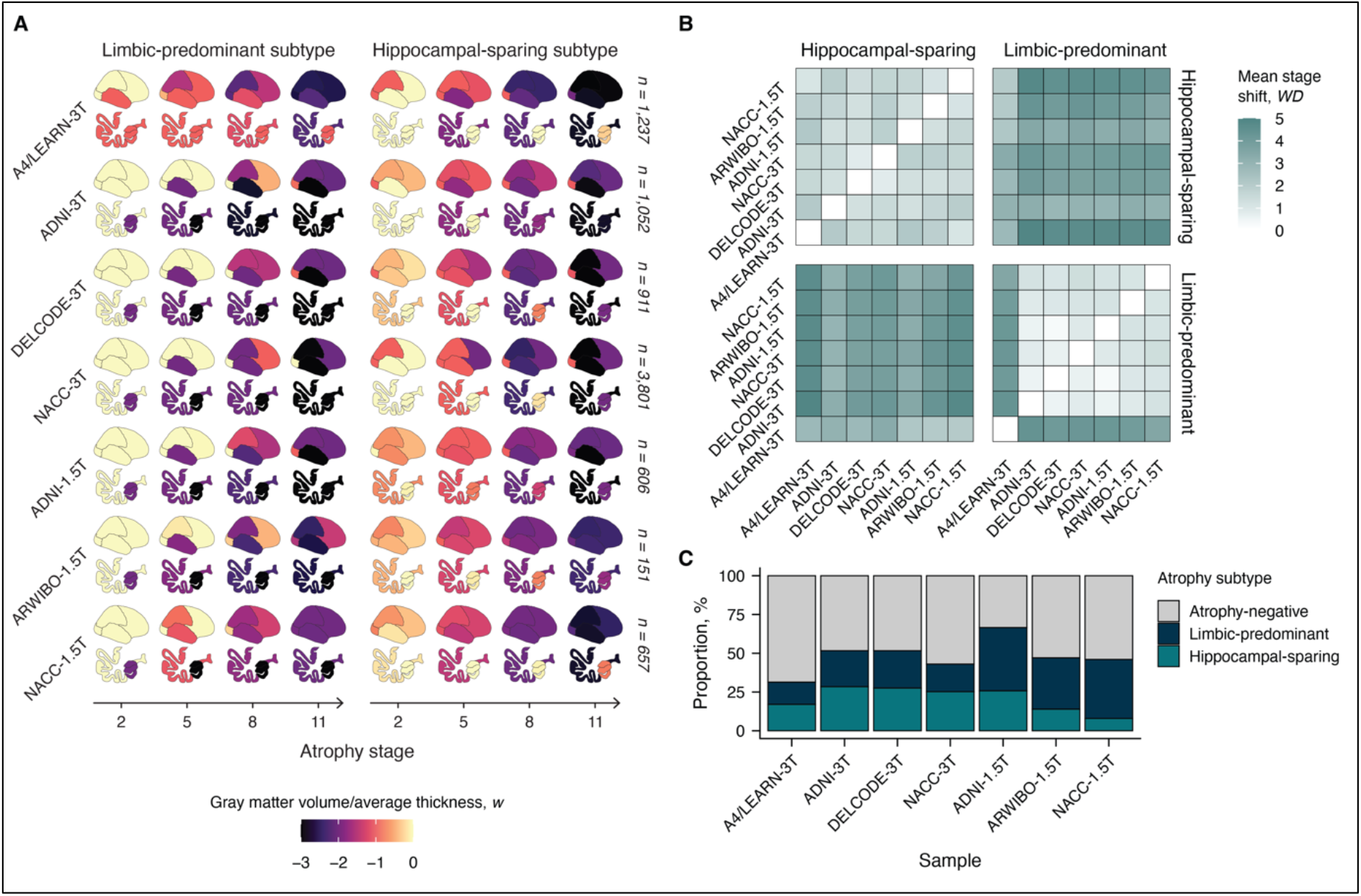
Independent identi4ication of limbic-predominant and hippocampal-sparing atrophy sequences within each sample. (**A**) Visualization of atrophy progression sequences within each atrophy subtype and sample. The top rows of the pictograms depict lateral views of the right cerebrum, whereas the bottom rows illustrate coronal sections of the left temporal lobe. (**B**) Average stage shift between atrophy sequences (*WD*). Values reelect the mean number of atrophy stages by which events would need to be displaced to align the two models. Lower values indicate higher sequence similarity. (**C**) Proportions of the two atrophy subtypes and of the atrophy-negative group (atrophy stage 0) across samples.

### PHASE–AD is cohort-independent and robustly classifies unseen MRI scans

We next trained PHASE–AD using the pooled data as a global model of atrophy progression. The resulting *w*-scored atrophy markers showed excellent agreement with those derived from within-sample modeling (mean ± *SD ICC*_2,1_ = .95 ± .03, all *p* < .001), resulting in the recovery of limbic-predominant and hippocampal-sparing atrophy sequences (Figure 2A,B; Supplementary Figure 7). Stage shifts between within-sample and global atrophy sequences were generally small (mean ± *SD WD*_limbic-predominant_ = 1.15 ± 1.93, *WD*_hippocampal-sparing_ = 1.71 ± 1.30; Figure 2B). An exception was the limbic-predominant subtype in the A4/LEARN-3T sample, which exhibited the highest dissimilarity relative to the corresponding subtype identified from the pooled data (mean *WD* = 4.13; otherwise mean *WD* ≤ 2.05). Modeling with up to five subtypes indicated the possible presence of a third subtype. However, this subtype did not closely correspond to any subtype identified in within-sample analyses (Supplementary Figure 9), and only a small proportion of participants with an atrophy stage > 0 were assigned to it (11.9%). Therefore, the two-subtype model was retained also for PHASE–AD. Distributions of atrophy subtype and stage across diagnostic groups are shown in Figure 2C,D.

**Figure 2.**
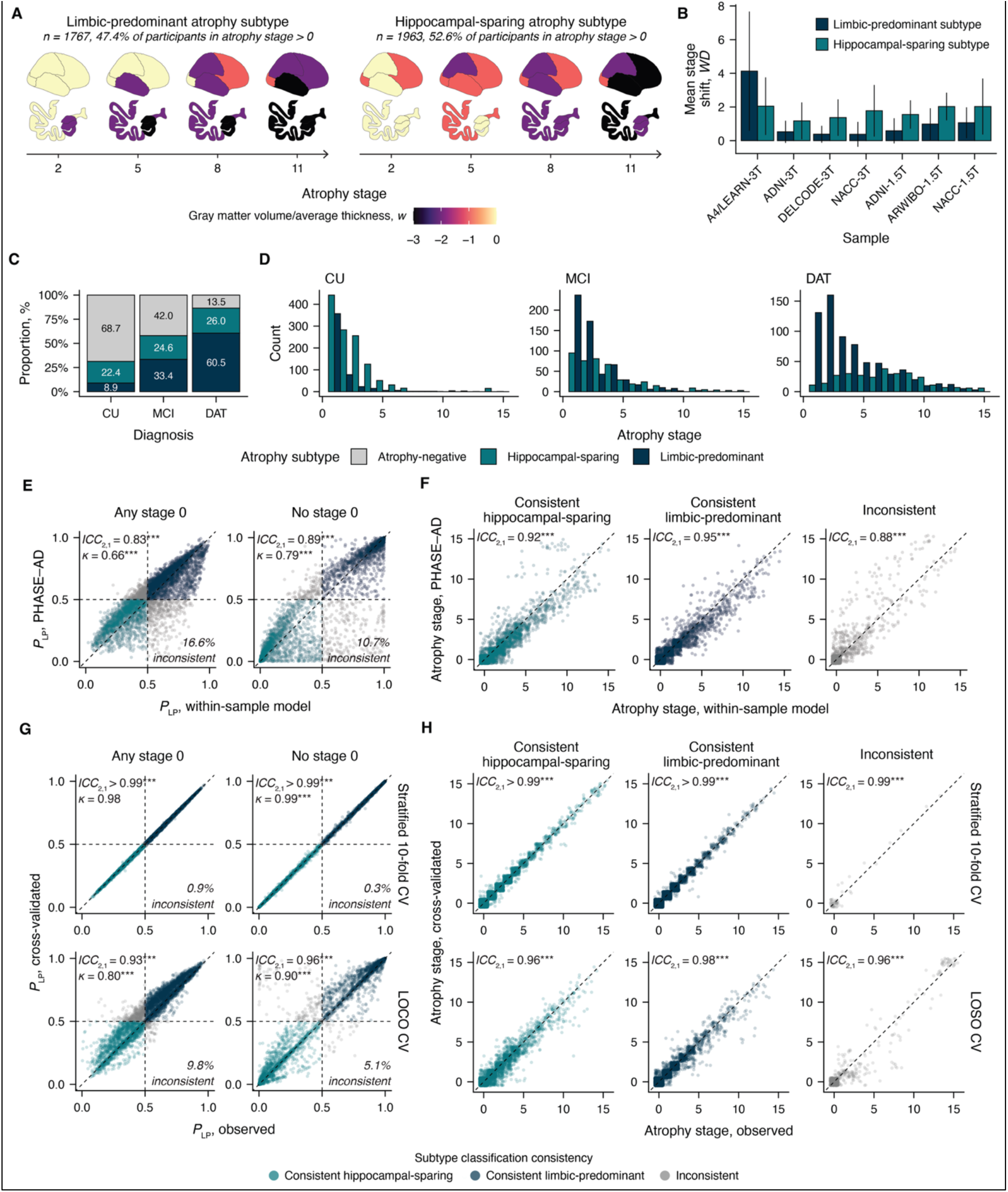
Description and cross-validation of PHASE–AD. (**A**) Limbic-predominant and hippocampal-sparing atrophy sequences identified from PHASE–AD using the pooled dataset (*N* = 8,415). The top rows of the pictograms depict lateral views of the right cerebrum, whereas the bottom rows illustrate coronal sections of the left temporal lobe. (**B**) Similarity of PHASE–AD sequences with their within-sample equivalents, quantified as average stage shifts (*WD*). Error bars indicate *SD*s. (**C**) Distribution of atrophy subtype classifications among diagnostic groups. (**D**) Distribution of atrophy stage classifications among diagnostic groups, only shown for individuals assigned an atrophy subtype. (**E**) Agreement of atrophy subtyping between within-sample models and PHASE–AD. Shown is the probability of limbic-predominant atrophy (*P*_LP_; the inverse of the probability of hippocampal-sparing atrophy). The bottom-left quadrant indicates participants assigned the hippocampal-sparing subtype by both models, and the top-right quadrant those assigned the limbic-predominant subtype. Data points outside these quadrants represent inconsistent subtype assignment and their proportion is indicated in the bottom-right corner of each plot. The left panel shows participants assigned stage 0 by either model, and the right panel shows all remaining participants. (**F**) Agreement of atrophy staging between within-sample models and PHASE– AD, stratified by subtype classification consistency. (**G**,**H**) Agreement of cross-validated and observed subtyping and staging, using stratified ten-fold cross-validation (top row) and leave-one-sample-out (LOSO) cross-validation (bottom row). Results are presented as in panels (**F**,**H**). All axes representing atrophy stage were jittered by up to ± 0.5 stages to improve visual clarity. \*\*\**p* < .001. Abbreviations: CV, cross-validation.

Agreement between within-sample models and PHASE–AD was also high for individual subtype and stage classifications (Figure 2E,F). Ten-fold stratified and LOSO cross-validation of PHASE–AD demonstrated overall robustness and supported generalizability to unseen data (Figure 2G,H). In the LOSO cross-validation scheme, atrophy stage classifications were robust to the exclusion of any individual sample, and subtype assignments were particularly consistent in participants classified at atrophy stage > 0 (Supplementary Figure 10). Supplementary Figure 11 summarizes robustness indices across cross-validation schemes, scanner manufacturers, and field strengths, with agreement being generally high across all groups.

### Atrophy subtype and stage relate to distinct clinical profiles

Meta-analyses of baseline demographic and clinical characteristics revealed several associations with PHASE–AD classifications that were independent of clinical diagnosis and other relevant covariates. The pooled effects, summarized in Figure 3, were largely consistent between the full samples and the Aβ-positive sub-samples. For readability, only pooled statistics from the full-sample analyses are reported in the following section. Supplementary Tables 2–5 present the individual results underlying the meta-analytical estimates presented in the main text.

**Figure 3.**
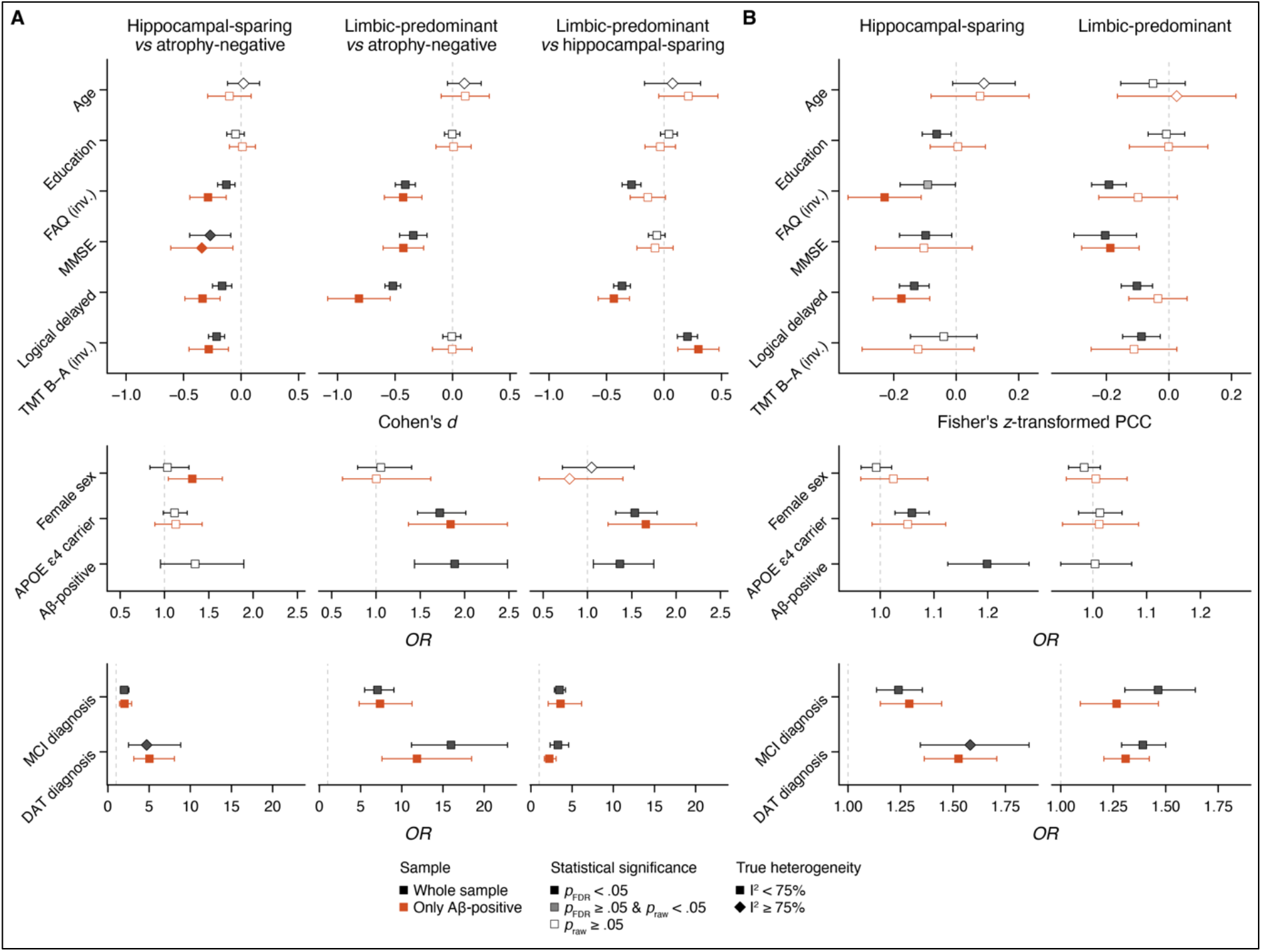
Cross-sectional associations of PHASE–AD classi4ications with demographic and clinical characteristics across samples. (**A**) Pairwise subtype comparisons are displayed using Cohen’s *d* for continuous variables and *OR* for binary outcomes. (**B**) Baseline relationships with atrophy stage are shown as Fisher’s *z*-transformed partial correlation coefficients (PCC) for continuous outcomes and *OR* for binary outcomes. All continuous outcome variables and predictors were *z*-standardized to the CU group within each sample. Exceptions are atrophy stages (*OR*s and longitudinal estimates can be interpreted as the effect of unscaled atrophy stage on the outcome) and FAQ scores (*z*-standardization occurred in reference to the entire samples due to low variance in the CU groups). All models included diagnostic group as a covariate. Models predicting Aβ-positivity were additionally adjusted for age, sex, and the interval between Aβ biomarker sampling and MRI. Models predicting neuropsychological test scores were further adjusted for age, sex, education, and the interval between testing and MRI. Models fitted in A4/LEARN-3T additionally controlled for study arm. Error bars represent 95% C.I.s. All samples with available outcomes were included in the meta-analysis. Results with I_2_ > 75% are marked with diamond symbols, indicating considerable heterogeneity among included studies. Abbreviations: inv., inverted. Logical delayed, Logical Memory Delayed Recall. PCC, partial correlation coefficient.

Regarding demographic associations, atrophy subtype and stage showed generally modest effects and high inter-sample variability for many estimates (I_2_ > 75%). Limbic-predominant atrophy was associated with higher odds of carrying at least one APOE ε4 allele than both the atrophy-negative group (*OR* = 1.72, *p*_FDR_ < .001) and the hippocampal-sparing subtype (*OR* = 1.53, *p*_FDR_ < .001), in both the whole and Aβ-positive samples. Aβ-positivity was also more prevalent in the limbic-predominant subtype compared with both the atrophy-negative group (*OR* = 1.89, *p*_FDR_ < .001) and the hippocampal-sparing subtype (*OR* = 1.37, *p*_FDR_ = .022). Although the rate of Aβ-positivity was numerically higher in the hippocampal-sparing subtype than in the atrophy-negative group (*OR* = 1.35, *p*_FDR_ = .129), this effect did not reach statistical significance and exhibited substantial heterogeneity across samples (I_2_ = 70.4%). Higher atrophy stage increased the odds of Aβ-positivity in the hippocampal-sparing (*OR* = 1.20, *p* < .001), but not the limbic-predominant subtype (*OR* = 1.00, *p* = .908). The odds of MCI and DAT diagnoses were highest in the limbic-predominant subtype, followed by the hippocampal-sparing subtype and the atrophy-negative group, across both the whole and Aβ-positive samples (all *p*_FDR_ < .001, pooled *OR*s reported in Supplementary Table 4). Within both subtypes, higher atrophy stage significantly increased the odds of either diagnosis (all *p*_FDR_ ≤ .005, pooled *OR*s reported in Supplementary Table 5).

Both atrophy subtypes showed worse scores than the atrophy-negative group on the MMSE (limbic-predominant: *d* = –0.34, *p*_FDR_ < .001; hippocampal-sparing: *d* = –0.27, *p*_FDR_ = .006) and the FAQ (limbic-predominant: *d* = 0.41, *p*_FDR_ < .001; hippocampal-sparing: *d* = 0.13, *p*_FDR_ = .002). Between-subtype differences were observed for the FAQ (*d* = 0.28, *p*_FDR_ < .001), but not the MMSE (*d* = –0.06, *p*_FDR_ = .129), and only in the whole samples. Meanwhile, the domain-specific tests showed distinct patterns; limbic-predominant atrophy was associated with worse Logical Memory Delayed Recall scores (*d* = –0.37, *p*_FDR_ < .001) while hippocampal-sparing atrophy was linked with worse TMT B–A scores (*d* = –0.20, *p*_FDR_ < .001) across samples. The only consistent effect of atrophy stage across both samples was a negative association between atrophy stage and Logical Memory Delayed Recall performance in the hippocampal-sparing subtype (*z*-transformed partial correlation coefficient = –0.13, *p*_FDR_ < .001).

### Atrophy subtype and stage predict regional tau burden

Two participants with available tau PET data were outliers due to extreme atrophy stage assignments (stage ≥ 14, both MCI) and were excluded from the analyses reported here (Supplementary Figure 12). Among participants whose tau PET signal was analyzed in relation to an MRI scan acquired at a different visit than the scan used for model training (*n* = 61), subtype agreement across acquisitions was excellent in those originally assigned an atrophy stage > 0 (*n* = 29), with only one inconsistent classification (96.55% agreement). Stage monotonicity was observed in 89.66% of cases (*n* = 26), with the remaining cases representing minimal violations (all single-stage decrements). Among those originally at atrophy stage 0, subtype classifications were consistent in 75.68% of cases (*n* = 28, Supplementary Figure 13).

Relationships between atrophy stages and regional tau PET signal were non-linear in both subtypes and across all CenTauR ROIs. In the limbic-predominant subtype, mesial temporal tau PET signal increased sharply in early atrophy stages relative to other ROIs, whereas the hippocampal-sparing subtype exhibited a more uniform increase across ROIs that emerged at later atrophy stages (Figure 4A,B). Including PHASE– AD classifications, rather than relying only on clinical and demographic variables, significantly improved model fit for tau PET signal across all ROIs (ΔAIC range: –76.86 to –43.89, Δ*R*_2_ range: 0.06–0.12; Figure 4C). Atrophy stage main effects and subtype-by-stage interactions were significant across all ROIs and atrophy stage spline terms. Subtype main effects were consistently non-significant, suggesting that the two subtypes diverge in tau burden across atrophy stages rather than at baseline (Supplementary Table 6). Within cohorts, this effect could be replicated after adjustments for plasma p-tau_217_ biomarkers for mesial temporal tau PET signal in A4 (ΔAIC = –6.56, Δ*R*_2_ = 0.05) and across all CenTauR ROIs in the combined ADNI sample (ΔAIC range: –2.41 to –14.12, Δ*R*_2_ range: 0.03–0.09; Supplementary Figure 14), though coefficient-level significance of stage and interaction terms was more variable across ROIs and cohorts compared to the primary analysis (Supplementary Table 7). Atrophy subtype and stage improved the prediction of tau burden groups beyond clinical and demographic variables (ΔAIC = –12.02, 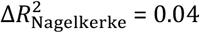 Figure 4D). This improvement was driven primarily by atrophy stage (*b* = 0.39, *z* = 2.68, *p* = .007), whereas the subtype main effect (*b* = 0.32, *z* = 0.73, *p* = .662) and the subtype-by-stage interaction (*b* = 0.07, *z* = 0.32, *p* = .082) were non-significant predictors. This effect was robust in a sub-sample approximating ATT eligibility (i.e., MCI and DAT diagnosis only, MMSE ≥ 19, no APOE ε4 homozygotes; *n* = 80; Supplementary Table 8).

**Figure 4.**
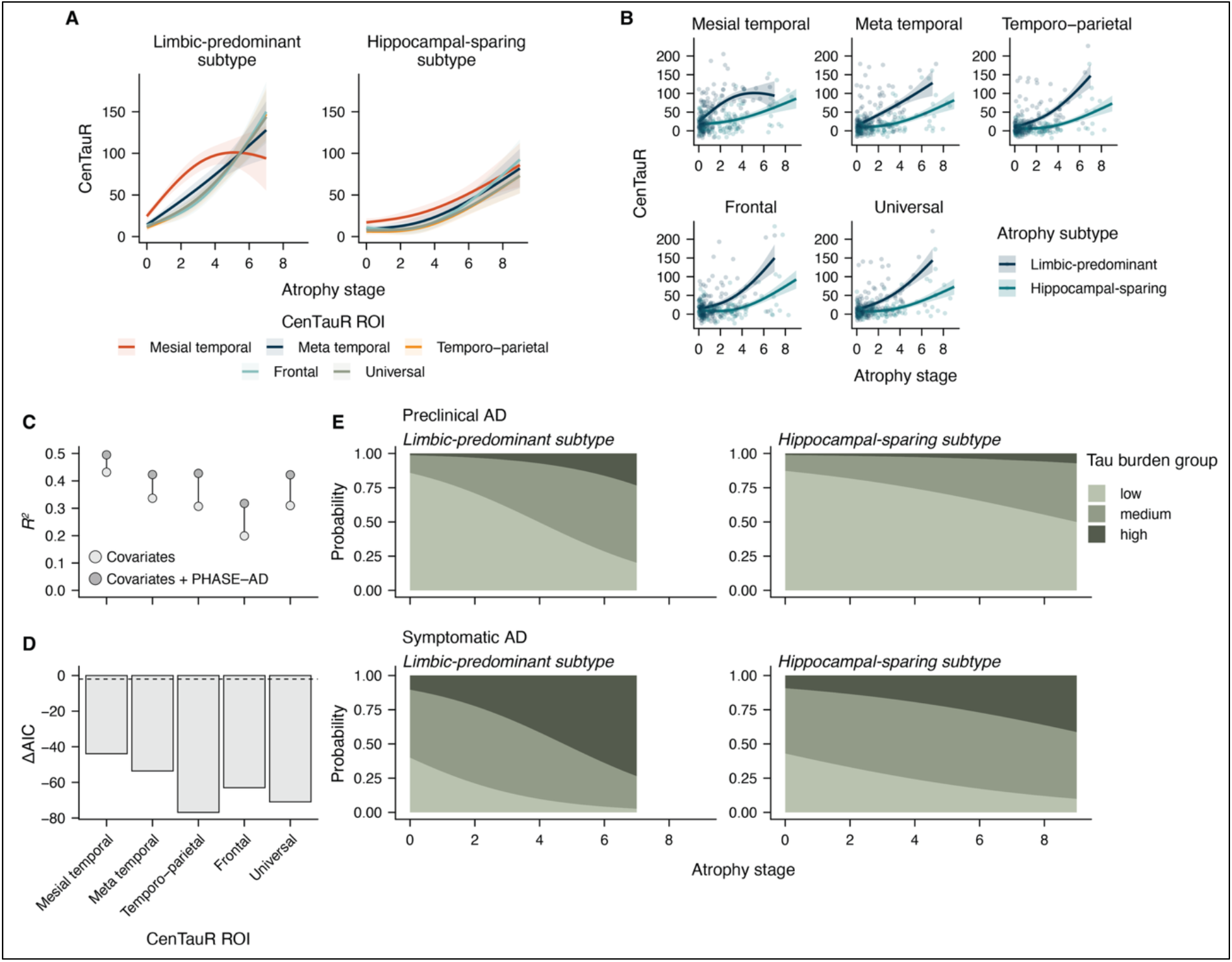
Associations between PHASE–AD classi4ications and regional tau burden in Aβ-positive participants. (**A**) Predicted trajectories of regional tau burden (expressed as CenTauR scores) across atrophy stages, stratified by CenTauR ROI and atrophy subtype. Lines represent model-based estimates from the best-fitting model, ribbons indicate 95% C.I.s. Predictions were generated across the observed stage range for each subtype. The PET–MRI time interval was fixed at the sample median. (**B**) Same trajectories with observed data points overlaid. (**C**,**D**) Model comparisons between covariates-only and full linear regression models including PHASE–AD classifications (atrophy subtype and stage) across CenTauR ROIs. (**C**) Total variance explained (*R*_2_) for each model. Lower points indicate the null model *R*_2_ (covariates only), upper points indicate the full model *R*_2_. (**D**) ΔAIC for full models relative to covariates-only models. The dashed line indicates ΔAIC = −2, prespecified as evidence of superior model fit. (**E**) Predicted probabilities of tau burden group across atrophy subtypes and stages, for individuals with preclinical AD (i.e., Aβ-positive CU participants; top panel) and symptomatic AD (i.e., Aβ-positive MCI and DAT participants; bottom panel) individuals. Predictions were evaluated over the observed stage range per subtype, with continuous covariates fixed at their sample medians, treatment set to placebo, and other categorical covariates averaged across included levels.

### PHASE–AD classifications predict longitudinal cognitive decline in preclinical and mild symptomatic Alzheimer’s disease

The addition of atrophy subtype and stage to covariate-only reference models improved model fit and increased explained variance both when predicting PACC–5 trajectories over 240 weeks in individuals with preclinical AD (A4; ΔAIC = –92.71, 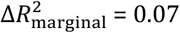) and when predicting CDR–SB trajectories over 180 weeks in individuals with mild symptomatic AD (ADNI; ΔAIC = –245.98, 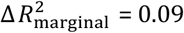). Significant time–by–atrophy classification interactions were observed in both samples and are reported in Supplementary Tables 9,10. Predicted outcome trajectories by atrophy subtype and stage are shown in Figure 5A,B for preclinical AD and Figure 5D,E for mild symptomatic AD. Pairwise contrasts revealed various significant differences in outcome change from baseline across atrophy subtype × stage strata (Figure 5C,F). These findings were independent of plasma p-tau_217_ biomarkers in both scenarios, as shown in additional analyses including plasma p-tau_217_ levels as a covariate (Supplementary Table 11,12). Atrophy subtype and stage outperformed MTA scores in explained variance and model fit in a mild symptomatic ADNI sub-sample (all MCI, *n* = 31; Figure 5G,H; Supplementary Table 13,14).

**Figure 5.**
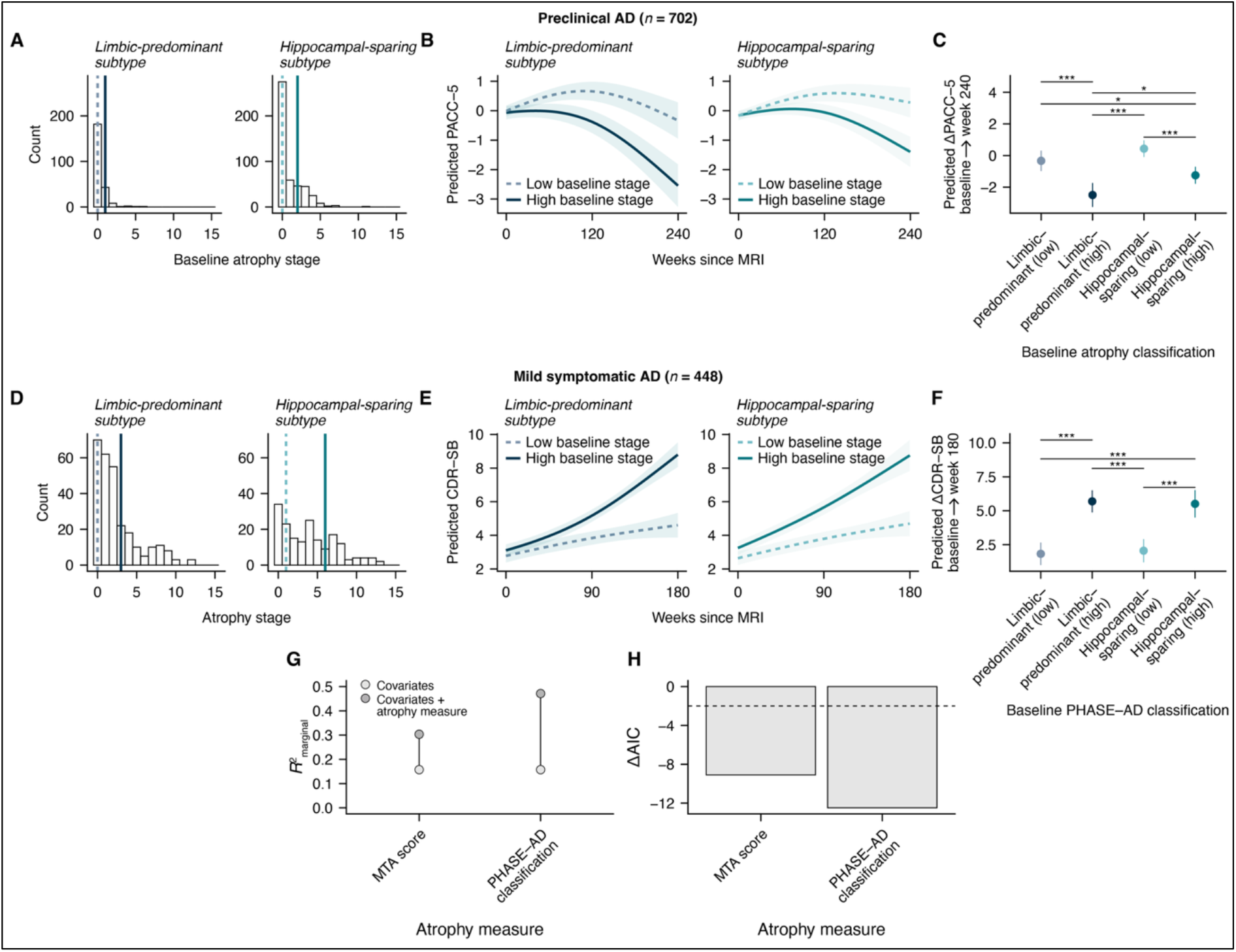
Prognostic value of PHASE–AD classi4ications for longitudinal cognitive decline. (**A**–**C**) Preclinical AD sample (A4, Aβ-positive; predicting PACC–5 over 240 weeks). (**D**–**F**) Mild symptomatic AD sample (ADNI, Aβ-positive, MCI and DAT, MMSE ≥ 19; predicting CDR–SB over 180 weeks). (**A**,**D**) Atrophy stage distributions within the limbic-predominant and hippocampal-sparing subtypes. Vertical dashed and solid lines indicate the 25^th^ and 75^th^ percentile stages, respectively, used to define low and high baseline stage levels for model predictions. (**B**,**E**) Outcome trajectories predicted from linear mixed effects models, stratified by subtype and baseline stage (low: dashed; high: solid). Predictions were evaluated with continuous covariates fixed at their sample medians, treatment set to placebo, and other categorical covariates averaged across included levels. Ribbons indicate 95% C.I.s. (**C**,**F**) Estimated outcome change per subtype × stage stratum from baseline to the maximum follow-up duration. (**G**) 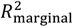 of models predicting CDR–SB in the mild symptomatic AD sample, comparing PHASE–AD classifications against MTA score as the atrophy measure. Lower points indicate the null model 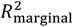 (covariates only), upper points indicate the full model 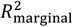. (**H**) Corresponding ΔAIC relative to the null model. The dashed line indicates ΔAIC = −2, prespecified as evidence of superior model fit. Error bars and ribbons indicate 95% C.I.s. \**p*_FDR_ < 0.05. ** *p*_FDR_ < 0.01. *** *p*_FDR_ < 0.001.

## Discussion

Structural MRI is routinely acquired in the clinical assessment of AD, yet quantitative morphometric indices derived from these scans rarely inform standard clinical workflows.^7^ The present study addresses this gap by introducing PHASE–AD as a universal atrophy classification framework that translates such indices into intuitive classifications of atrophy subtype and stage, evaluates its robustness and generalizability, and tests its translational utility in two clinically relevant application contexts. Specifically, limbic-predominant and hippocampal-sparing atrophy subtypes were replicated across seven independent samples that varied by recruitment settings, MRI acquisition parameters, and clinical composition. Next, PHASE–AD was trained on a pooled dataset of 8,415 individuals, yielded atrophy subtype and stage classifications that mirrored these sample-specific results, and generalized well to held-out samples in two cross-validation schemes. Individual atrophy subtype and stage classifications were associated with distinct clinical profiles across samples, including an amnestic-dysexecutive neuropsychological dissociation between the two subtypes. In addition, PHASE–AD classifications related to diverging tau PET signatures in both spatial distribution and overall burden, and jointly stratified longitudinal cognitive trajectories in preclinical and mild symptomatic AD.

The architecture of PHASE–AD was directly informed by factors that we identified as likely barriers to adopting quantitative morphometrics in routine clinical practice. Processing time was substantially reduced by replacing traditional segmentation and surface reconstruction with a deep learning-based alternative.^44^ The interpretive burden associated with multi-regional morphometric output was addressed by distilling this information into a disease-specific classification scheme.^7^ Generalizability and clinical applicability were pursued through normative modeling using a large and diverse reference sample (*n* = 5,513). The substantial overlap between within-sample modeling results, combined with robust performance across two cross-validation schemes, supports that PHASE–AD generalizes to unseen data without requiring a locally acquired reference sample of healthy controls. Complementing this, PHASE–AD accommodates the three scanner manufacturers and two field strengths most commonly encountered in clinical and research MRI settings. Robustness indices were high overall but marginally lower for 1.5T than 3T acquisitions, likely reflecting the relative underrepresentation of 1.5T data in modern publicly available cohorts, and may improve with the addition of targeted training data.

The replication of limbic-predominant and hippocampal-sparing subtypes across the broad majority of analyzed samples supports the view that atrophy heterogeneity in AD largely reduces to a single dimension anchored by these two extremes.^18^ Although the primary training sample lacked AD biomarker characterization, successful replication of the identified atrophy cascades in biological AD samples provides strong evidence that the model captures structural heterogeneity within the contemporary biological definition of AD.^4^ This interpretation is further supported by the broader literature, in which data-driven subtyping studies have consistently identified similar atrophy patterns, albeit under varying nomenclature and sometimes alongside additional subtypes.^18–20,23,51,52^ Meanwhile, two samples deviated from the overall replication pattern and warrant further discussion. In ARWIBO-1.5T, the pre-specified model selection criterion favored a one-subtype solution, likely due to insufficient power to detect multiple subtypes (10.07 observations per modeled event; recommended minimum 10–20, ref.^53^). Notably, the two-subtype model was nevertheless highly concordant with those identified in other samples, suggesting the underlying subtype structure was present but not reliably detected. In A4/LEARN-3T, the two-subtype model was selected as the best-fitting solution, but the limbic-predominant subtype was less distinctly expressed relative to other samples. This sample consisted exclusively of CU individuals, which probably constrained the reconstruction of full atrophy cascades due to limited variance in brain structure. Additionally, our finding of elevated odds of MCI and DAT diagnoses associated with the limbic-predominant subtype is consistent with the underrepresentation of this subtype in the A4/LEARN-3T sample. Nonetheless, PHASE– AD classifications in A4/LEARN-3T showed various significant cross-sectional and longitudinal clinical associations, supporting the framework’s relevance even in this early-stage population.

The meta-analytically derived clinical characteristics associated with atrophy subtype and stage were mostly robust across samples, as indicated by low-to-moderate indices of between-sample heterogeneity. The finding of higher odds of APOE ε4 carriership in the limbic-predominant subtype aligns with previous work and may reflect an increased vulnerability of the MTL to tau pathology in APOE ε4 carriers.^54,55^ While the biological mechanisms underlying this vulnerability are still unclear, proposed mechanisms include increased neuronal excitability and local blood-brain barrier dysfunction within the MTLs of APOE ε4 carriers.^56,57^ Consistent with the role of the APOE ε4 allele as a genetic risk factor for Aβ pathology, the limbic-predominant atrophy pattern was associated with increased odds of Aβ-positivity in our study.^58^ Limbic-predominant atrophy was related to greater odds of MCI and DAT than hippocampal-sparing atrophy, and within both subtypes, a higher atrophy stage further increased these odds. When these effects of diagnosis were controlled for, the two subtypes showed only subtle differences in global cognition and instrumental activities of daily living. Yet, subtype differences became evident at the level of individual cognitive domains, where limbic-predominant atrophy was associated with greater episodic memory impairment, and hippocampal-sparing atrophy was characterized by worse executive functioning. These results mirror established atrophy signatures of amnestic and non-amnestic AD phenotypes (ref.^22^) as well as data-driven biological subtyping studies in AD (refs.^19,20,51^).

The higher mesial temporal tau burden in limbic-predominant relative to hippocampal-sparing atrophy suggests that atrophy subtypes in AD — at least partly (ref.^59^)— reflect the downstream structural consequences of diverging tau pathology.^25,60^ The absence of a clear tau hotspot in the hippocampal-sparing subtype may reflect the fact that many regions characteristically affected by tau pathology in atypical AD variants are not represented as isolated ROIs within the CenTauR framework.^53,61^ Moreover, this subtype potentially aggregates multiple atypical tau accumulation patterns rather than representing a single coherent subtype of tau accumulation.^53,61^

The association between PHASE–AD classification and tau burden group carries direct practical implications. Phase 3 trial data for donanemab have demonstrated highest treatment efficacy in low-to-medium tau burden groups.^27^ In this study, we demonstrate that analogous tau burden groups can be predicted via atrophy subtype and stage, which raises the question of whether they could serve as predictive markers of treatment response for a drug already in clinical use. Beyond this predictive use case, PHASE–AD classifications could serve as a rule-out marker in trials implementing recruitment strategies based on tau burden. Individuals unlikely to fall within a targeted tau burden range could be identified from their atrophy profile alone, thereby reducing the number of required tau PET acquisitions. While we show that PHASE–AD subtypes track with tau PET signal, atrophy should not be read as a one-to-one signature of any single pathology. Co-existing pathologies may push the atrophy pattern in different directions, and distinct pathological profiles may converge on the very same atrophy subtype.

Our prognostic analyses indicate that PHASE–AD classifications are related not only to current disease severity but also to an individual’s longitudinal cognitive trajectory. Notably, these effects were observed not only in mild symptomatic AD but also in preclinical AD, complementing prior studies showing that structural brain heterogeneity relates to diverging cognitive trajectories before symptom onset.^19,51^ In fact, the added value of automated structural assessments may be greatest at this early stage, where the subtle atrophy patterns associated with accelerated cognitive decline are difficult to detect through visual inspection. The more pronounced subtype effects on PACC–5 relative to CDR–SB trajectories are consistent with the amnestic profile cross-sectionally associated with limbic-predominant atrophy and the episodic memory weighting of the PACC–5, whereas the CDR–SB reflects a broader composite of cognition and functional ability. The significant and complementary prognostic effects of atrophy subtype and stage across both scenarios have direct implications for trial design, as they suggest that PHASE–AD can support prognostic stratification, enrichment, or sample homogenization using an imaging modality already routinely acquired during trial recruitment. Notably, our exploratory comparison with MTA scores suggested greater prognostic utility of PHASE–AD over this clinically established measure.

While our study supports the applied value of AD subtyping and staging based on MRI, atrophy assessment captures only one portion of the AD disease cascade. A complete picture of an individual case with its own set of risk and protective factors requires a multimodal integration of atrophy subtype and stage with upstream factors (e.g., genetic risk, proteinopathy, neuroinflammation) and downstream outcomes (e.g., cognitive and functional impairment). Parallel efforts to make assessments of these portions increasingly accessible and precise include blood-based biomarkers of AD pathology and remote cognitive testing.^62,63^ In our study, the added value of MRI-based atrophy subtyping and staging beyond plasma p-tau_217_ highlights the importance of characterizing the full AD disease cascade rather than relying on any single marker.

The present study should be interpreted considering its limitations. First, PHASE–AD does not account for hemispheric lateralization, as all morphometric markers were averaged across hemispheres. This is a meaningful omission given that lateralization patterns have been described for both typical and atypical clinical phenotypes, as well as data-driven biological subtypes of AD.^53,64^ Second, although the included cohorts vary across many relevant characteristics, participants recruited into observational and interventional research studies remain imperfect representatives of the broader clinical population this study ultimately targets, particularly with respect to the representation of marginalized groups.^65,66^ Validation studies evaluating the framework in real-world clinical workflows are therefore needed. Finally, while the comparison with MTA scores provides an encouraging proof of concept, replication in larger samples is necessary to draw conclusions about prognostic utility of automated atrophy subtyping and staging compared with current clinical standards.

In conclusion, PHASE–AD represents a universal framework for characterizing atrophy heterogeneity and progression in AD, designed to address the barriers that have limited the broad implementation of quantitative morphometrics in routine clinical practice. The results provide strong evidence that the biological heterogeneity of AD is reflected in limbic-predominant and hippocampal-sparing atrophy subtypes with diverging downstream clinical profiles. The per-scan classifications generated by PHASE–AD offer substantial utility for inferring regional tau accumulation patterns without reliance on molecular imaging, and provide prognostic information in prospective settings directly relevant to contemporary trial design in AD.

## Data Availability

DELCODE data, study protocol and biomaterials can be shared with partners based on individual data and biomaterial transfer agreements. Data from the other included cohorts can be requested at the respective managing institutions.

## Competing interests

H.B. has received honoraria from Elsevier. D.B. has received speaker fees from Eli Lilly and Company. The other authors declare no competing interests.

## Acknowledgements

The authors would like to thank and acknowledge all study participants as well as their study partners and caregivers.

The A4 Study was a secondary prevention trial in preclinical Alzheimer’s disease, aiming to slow cognitive decline associated with brain amyloid accumulation in clinically normal older individuals. The A4 Study was funded by a public-private-philanthropic partnership, including funding from the National Institutes of Health-National Institute on Aging, Eli Lilly and Company, Alzheimer’s Association, Accelerating Medicines Partnership, GHR Foundation, an anonymous foundation, and additional private donors, with in-kind support from Avid Radiopharmaceuticals, Cogstate, Albert Einstein College of Medicine and the Foundation for Neurologic Diseases. The companion observational Longitudinal Evaluation of Amyloid Risk and Neurodegeneration (LEARN) Study was funded by the Alzheimer’s Association and GHR Foundation. The A4 and LEARN Studies were led by Dr. Reisa Sperling at Brigham and Women’s Hospital, Harvard Medical School, and Dr. Paul Aisen at the Alzheimer’s Therapeutic Research Institute (ATRI) at the University of Southern California. The A4 and LEARN Studies were coordinated by ATRI at the University of Southern California, and the data are made available under the auspices of Alzheimer’s Clinical Trial Consortium through the Global Research & Imaging Platform (GRIP). The complete A4 Study Team list is available on: https://www.actcinfo.org/a4-study-team-lists/. We would like to acknowledge the dedication of the study participants and their study partners who made the A4 and LEARN Studies possible.

Data collection and sharing for this project was funded by the Alzheimer’s Disease Neuroimaging Initiative (ADNI) (National Institutes of Health Grant U01 AG024904) and DOD ADNI (Department of Defense award number W81XWH-12-2-0012). ADNI is funded by the National Institute on Aging, the National Institute of Biomedical Imaging and Bioengineering, and through generous contributions from the following: AbbVie, Alzheimer’s Association; Alzheimer’s Drug Discovery Foundation; Araclon Biotech; BioClinica, Inc.; Biogen; Bristol-Myers Squibb Company; CereSpir, Inc.; Cogstate; Eisai Inc.; Elan Pharmaceuticals, Inc.; Eli Lilly and Company; EuroImmun; F. Hoffmann-La Roche Ltd and its afeiliated company Genentech, Inc.; Fujirebio; GE Healthcare; IXICO Ltd.; Janssen Alzheimer Immunotherapy Research & Development, LLC.; Johnson & Johnson Pharmaceutical Research & Development LLC.; Lumosity; Lundbeck; Merck & Co., Inc.; Meso Scale Diagnostics, LLC.; NeuroRx Research; Neurotrack Technologies; Novartis Pharmaceuticals Corporation; Peizer Inc.; Piramal Imaging; Servier; Takeda Pharmaceutical Company; and Transition Therapeutics. The Canadian Institutes of Health Research is providing funds to support ADNI clinical sites in Canada. Private sector contributions are facilitated by the Foundation for the National Institutes of Health (www.fnih.org). The grantee organization is the Northern California Institute for Research and Education, and the study is coordinated by the Alzheimer’s Therapeutic Research Institute at the University of Southern California. ADNI data are disseminated by the Laboratory for Neuro Imaging at the University of Southern California.

Data collection and sharing for this project was supported by the Italian Ministry of Health, under the following grant agreements: Ricerca Corrente IRCCS Fatebenefratelli, Linea di Ricerca 2; Progetto Finalizzato Strategico 2000-2001 “Archivio normativo italiano di morfometria cerebrale con risonanza magnetica (età 40+)”; Progetto Finalizzato Strategico 2000-2001 “Decadimento cognitivo lieve non dementigeno: stadio preclinico di malattia di Alzheimer e demenza vascolare. Caratterizzazione clinica, strumentale, genetica e neurobiologica e sviluppo di criteri diagnostici utilizzabili nella realtà nazionale,”; Progetto Finalizzata 2002 “Sviluppo di indicatori di danno cerebrovascolare clinicamente signieicativo alla risonanza magnetica strutturale”; Progetto Fondazione CARIPLO 2005-2007 “Geni di suscettibilità per gli endofenotipi associati a malattie psichiatriche e dementigene”; “Fitness and Solidarietà”; and anonymous donors.

DELCODE is funded by Clinical Research, German Center for Neurodegenerative Disorders. This work was supported by the German Research Foundation (Deutsche Forschungsgemeinschaft, DFG; Project ID 374011584/3T Ganzkörper MR-Tomograf, to P.D.; Project ID 425899996 – CRC 1436).

The NACC database is funded by NIA/NIH Grant U24 AG072122. NACC data are contributed by the NIA-funded ADRCs: P30 AG062429 (PI James Brewer, MD, PhD), P30 AG066468 (PI Oscar Lopez, MD), P30 AG062421 (PI Teresa Gomez-Isla, MD), P30 AG066509 (PI Thomas Grabowski, MD), P30 AG066514 (PI Mary Sano, PhD), P30 AG066530 (PI Helena Chui, MD, Arthur Toga, PhD), P30 AG066507 (PI Marilyn Albert, PhD), P30 AG066444 (PI David Holtzman, MD), P30 AG066518 (PIs Lisa Silbert, MD, Kevin Duff, PhD), P30 AG066512 (PI Thomas Wisniewski, MD), P30 AG066462 (PI Scott Small, MD), P30 AG072979 (PI David Wolk, MD), P30 AG072972 (PIs Charles DeCarli, MD, Rachel Whitmer, PhD), P30 AG072976 (PI Andrew Saykin, PsyD), P30 AG072975 (PI Julie Schneider, MD, MS), P30 AG072978 (PI Ann McKee, MD), P30 AG072977 (PI Robert Vassar, PhD), P30 AG066519 (PI Joshua Grill, PhD), P30 AG062677 (PIs Brad Boeve, MD, Ronald Petersen, MD, PhD), P30 AG079280 (PI Jessica Langbaum, PhD), P30 AG062422 (PI Gil Rabinovici, MD), P30 AG066511 (PI Allan Levey, MD, PhD), P30 AG072946 (PI Linda Van Eldik, PhD), P30 AG062715 (PI Sanjay Asthana, MD, FRCP), P30 AG072973 (PI Russell Swerdlow, MD), P30 AG066506 (PIs Glenn Smith, PhD, ABPP, David Lowenstein, PhD, Ranjan Duara, MD), P30 AG066508 (PIs Stephen Strittmatter, MD, PhD, Christopher Van Dyck, MD), P30 AG066515 (PI Victor Henderson, MD, MS), P30 AG072947 (PI Suzanne Craft, PhD), P30 AG072931 (PI Henry Paulson, MD, PhD), P30 AG066546 (PIs Sudha Seshadri, MD, Gladys Maestre, MD, PhD), P30 AG086401 (PI Erik Roberson, MD, PhD), P30 AG086404 (PI Gary Rosenberg, MD), P30 AG086403 (PI Angela Jefferson, PhD), P30 AG072958 (PIs Heather Whitson, MD, Gwenn Garden, MD, PhD), P30 AG072959 (PI Jagan Pillai, MD, PhD), P30 AG092752 (Ihab Hajjar, MD, MS).

SCAN is a multi-institutional project that was funded as a U24 grant (AG067418) by the National Institute on Aging in May 2020. Data collected by SCAN and shared by NACC are contributed by the NIA-funded ADRCs as follows: Arizona Alzheimer’s Center – P30 AG072980 (PI: Eric Reiman, MD); R01 AG069453 (PI: Eric Reiman (contact), MD); P30 AG019610 (PI: Eric Reiman, MD); and the State of Arizona which provided additional funding supporting our center; Boston University – P30 AG013846 (PI Neil Kowall MD); Cleveland ADRC – P30 AG062428 (James Leverenz, MD); Cleveland Clinic, Las Vegas – P20AG068053; Columbia – P50 AG008702 (PI Scott Small MD); Duke/UNC ADRC – P30 AG072958; Emory University – P30AG066511 (PI Levey Allan, MD, PhD); Indiana University – R01 AG19771 (PI Andrew Saykin, PsyD); P30 AG10133 (PI Andrew Saykin, PsyD); P30 AG072976 (PI Andrew Saykin, PsyD); R01 AG061788 (PI Shannon Risacher, PhD); R01 AG053993 (PI Yu-Chien Wu, MD, PhD); U01 AG057195 (PI Liana Apostolova, MD); U19 AG063911 (PI Bradley Boeve, MD); and the Indiana University Department of Radiology and Imaging Sciences; Johns Hopkins – P30 AG066507 (PI Marilyn Albert, Phd.); Mayo Clinic – P50 AG016574 (PI Ronald Petersen MD PhD); Mount Sinai – P30 AG066514 (PI Mary Sano, PhD); R01 AG054110 (PI Trey Hedden, PhD); R01 AG053509 (PI Trey Hedden, PhD); New York University – P30AG066512-01S2 (PI Thomas Wisniewski, MD); R01AG056031 (PI Ricardo Osorio, MD); R01AG056531 (PIs Ricardo Osorio, MD; Girardin Jean-Louis, PhD); Northwestern University – P30 AG013854 (PI Robert Vassar PhD); R01 AG045571 (PI Emily Rogalski, PhD); R56 AG045571, (PI Emily Rogalski, PhD); R01 AG067781, (PI Emily Rogalski, PhD); U19 AG073153, (PI Emily Rogalski, PhD); R01 DC008552, (M.-Marsel Mesulam, MD); R01 AG077444, (PIs M.-Marsel Mesulam, MD, Emily Rogalski, PhD); R01 NS075075 (PI Emily Rogalski, PhD); R01 AG056258 (PI Emily Rogalski, PhD); Oregon Health and Science University – P30 AG008017 (PI Jeffrey Kaye MD); R56 AG074321 (PI Jeffrey Kaye, MD); Rush University – P30 AG010161 (PI David Bennett MD); Stanford – P30AG066515; P50 AG047366 (PI Victor Henderson MD MS); University of Alabama, Birmingham – P20; University of California, Davis – P30 AG10129 (PI Charles DeCarli, MD); P30 AG072972 (PI Charles DeCarli, MD); University of California, Irvine – P50 AG016573 (PI Frank LaFerla PhD); University of California, San Diego – P30AG062429 (PI James Brewer, MD, PhD); University of California, San Francisco – P30 AG062422 (Rabinovici, Gil D., MD); University of Kansas – P30 AG035982 (Russell Swerdlow, MD); University of Kentucky – P30 AG028283-15S1 (PIs Linda Van Eldik, PhD and Brian Gold, PhD); University of Michigan ADRC – P30AG053760 (PI Henry Paulson, MD, PhD) P30AG072931 (PI Henry Paulson, MD, PhD) Cure Alzheimer’s Fund 200775 – (PI Henry Paulson, MD, PhD) U19 NS120384 (PI Charles DeCarli, MD, University of Michigan Site PI Henry Paulson, MD, PhD) R01 AG068338 (MPI Bruno Giordani, PhD, Carol Persad, PhD, Yi Murphey, PhD) S10OD026738-01 (PI Douglas Noll, PhD) R01 AG058724 (PI Benjamin Hampstead, PhD) R35 AG072262 (PI Benjamin Hampstead, PhD) W81XWH2110743 (PI Benjamin Hampstead, PhD) R01 AG073235 (PI Nancy Chiaravalloti, University of Michigan Site PI Benjamin Hampstead, PhD) 1I01RX001534 (PI Benjamin Hampstead, PhD) IRX001381 (PI Benjamin Hampstead, PhD); University of New Mexico – P20 AG068077 (Gary Rosenberg, MD); University of Pennsylvania – State of PA project 2019NF4100087335 (PI David Wolk, MD); Rooney Family Research Fund (PI David Wolk, MD); R01 AG055005 (PI David Wolk, MD); University of Pittsburgh – P50 AG005133 (PI Oscar Lopez MD); University of Southern California – P50 AG005142 (PI Helena Chui MD); University of Washington – P50 AG005136 (PI Thomas Grabowski MD); University of Wisconsin – P50 AG033514 (PI Sanjay Asthana MD FRCP); Vanderbilt University – P20 AG068082; Wake Forest – P30AG072947 (PI Suzanne Craft, PhD); Washington University, St. Louis – P01 AG03991 (PI John Morris MD); P01 AG026276 (PI John Morris MD); P20 MH071616 (PI Dan Marcus); P30 AG066444 (PI John Morris MD); P30 NS098577 (PI Dan Marcus); R01 AG021910 (PI Randy Buckner); R01 AG043434 (PI Catherine Roe); R01 EB009352 (PI Dan Marcus); UL1 TR000448 (PI Brad Evanoff); U24 RR021382 (PI Bruce Rosen); Avid Radiopharmaceuticals / Eli Lilly; Yale – P50 AG047270 (PI Stephen Strittmatter MD PhD); R01AG052560 (MPI: Christopher van Dyck, MD; Richard Carson, PhD); R01AG062276 (PI: Christopher van Dyck, MD); 1Florida – P30AG066506-03 (PI Glenn Smith, PhD); P50 AG047266 (PI Todd Golde MD PhD).

## Notes

### Author Declarations

All data was de-identified prior to study initiation.

