## Supplementary Material for "Subtyping and staging of Alzheimer’s disease from routine structural MRI with PHASE–AD"

Baumeister et al.

#### Supplementary methods

##### Inclusion criteria

The following descriptions provide an overview of the main inclusion criteria for each cohort study. They are not intended to be exhaustive. Readers are referred to the respective online resources and clinical trial registry entries for complete and detailed information.

###### *A4/LEARN*

The A4 study (NCT02008357, see https://clinicaltrials.gov) was conducted at 68 locations in the US, Canada, Australia, and Japan. It included clinically normal older adults with evidence of brain amyloid pathology. Inclusion criteria required a MMSE score between 25 and 30, a global CDR score of 0, and a Logical Memory II score between 6 and 18 at screening. Participants also had to show evidence of brain amyloid pathology on $^{18}$F-florbetapir PET and have a study partner with at least weekly contact who could provide collateral information (in person, by phone, or electronically). Key exclusion criteria included treatment with acetylcholinesterase inhibitors or memantine at screening or baseline, inadequate venous access, unstable or serious medical conditions, recent serious infectious disease affecting the brain, history of malignancy (except certain treated *in situ* cancers), history of human immunodeficiency virus (HIV), severe drug allergies or hypersensitivity reactions, major psychiatric illness (e.g., major depression or bipolar disorder in the past 2 years), chronic alcohol or drug abuse/dependence in the past 5 years, or serious suicide risk.

The A4 trial was a randomized, double-blind, placebo-controlled trial of with an open-label extension. During the double-blind period (240 weeks), participants were assigned to either intravenous (IV) solanezumab, a monoclonal antibody targeting monomeric amyloid, or placebo every 4 weeks. In the active treatment arm, solanezumab was administered in escalating doses of 400 mg, 800 mg, and then 1600 mg IV every 4 weeks. In the placebo arm, participants received matching placebo infusions on the same schedule. Following completion of the double-blind phase, all participants entered an open-label extension in which they received 1600 mg solanezumab IV every 4 weeks for an additional 204 weeks, resulting in a total treatment duration of approximately 444 weeks. After 240 weeks, the A4 trial did not show a significant slowing of cognitive decline on its primary endpoint in participants receiving solanezumab compared to those receiving placebo.[1]

The LEARN study (NCT02488720, see https://clinicaltrials.gov) was conducted at 40 U.S. locations and recruited individuals who had consented to the A4 trial and met its demographic, cognitive, and clinical criteria but did not meet the amyloid PET threshold required for randomization into the treatment arms of A4. Additional inclusion criteria required stable use of permitted medications for at least 8 weeks prior to baseline, the availability of a reliable study partner with sufficient contact to provide meaningful functional information, and willingness and ability to comply with study procedures for up to 240 weeks. Participants also needed adequate English or Spanish literacy and sufficient vision and hearing to complete psychometric testing. Exclusion criteria included treatment with acetylcholinesterase inhibitors or memantine at baseline, unstable or serious medical illness, contraindications for MRI, MRI findings of more than four hemosiderin deposits, or amyloid-related imaging abnormalities (ARIA-E), and use of exclusionary medications with significant central nervous system anticholinergic effects. Individuals currently enrolled in other interventional clinical trials or incompatible medical research were also excluded. For participants opting into the lumbar puncture sub-study, current use of anticoagulants such as warfarin or dabigatran was an additional exclusion criterion.

###### *ADNI*

Data used in the preparation of this article were obtained from the Alzheimer's Disease Neuroimaging Initiative (ADNI) database (adni.loni.usc.edu). The ADNI was launched in 2003 as a public-private partnership, led by Principal Investigator Michael W. Weiner, MD. The primary goal of ADNI has been to test whether serial magnetic resonance imaging (MRI), positron emission tomography (PET), other biological markers, and clinical and neuropsychological assessment can be combined to measure the progression of mild cognitive impairment and early Alzheimer's disease (AD).

In addition to ADNI's and our own general inclusion criteria, we included only participants whose MRI scans had been successfully processed with FreeSurfer, as documented by the University of California, San Francisco. This ensured the exclusion of scans more likely to yield low-quality FastSurfer segmentations.

*ADNI-1*

To be eligible for enrollment in ADNI-1 (NCT00106899, see https://clinicaltrials.gov), participants were required to have a Hachinski score ≤ 4, be between 55 and 90 years of age, have a Geriatric Depression Scale score < 6, have an available study partner with at least 10 hours contact per week and who is able to accompany study visits, have visual and auditory acuity adequate for neuropsychological testing, be in good health with no diseases precluding enrollment, be willing to complete a three-year imaging study (two years for DAT participants), have six grades education or work history, be fluent in English or Spanish, express commitment to neuroimaging and have no contraindications to MRI, agree to blood collection for APOE genotyping and DNA banking, agree to blood and urine collection for biomarker testing, and not be enrolled in other trials or studies. In addition, Women had to be post-menopausal for at least two years or surgically sterile.

The group-specific inclusion criteria for CU participants included reporting no memory complaints beyond what is common for their age, normal memory performance based on education-adjusted cut-offs on the delayed paragraph recall scores from the Wechsler Memory Scale – Revised (WMS-R), an MMSE score ≥ 24, a global CDR score of 0 (with a Memory Box score of 0), and no significant impairment in cognition or daily functioning. The MCI group included individuals who reported a memory complaint (or whose study partner reported one, with confirmation by the partner), abnormal memory performance based on education-adjusted cut-offs on the WMS-R delayed recall, an MMSE score ≥ 24, a CDR score of 0.5 (with a Memory Box score of 0.5), and general cognitive and functional performance insufficient for a diagnosis of DAT by the site physician. The DAT group included individuals who reported a memory complaint (or whose study partner did, with confirmation by the partner), had abnormal memory performance on the WMS-R delayed recall, an MMSE score between 20 and 26, a CDR score of 0.5 or 1.0, and met the NINCDS-ADRDA criteria for probable AD.

*ADNI-GO*

To be eligible for enrollment in ADNI-GO (NCT01078636, see https://clinicaltrials.gov), participants were required to have a Hachinski score ≤ 4, be between 55 and 90 years of age, have a Geriatric Depression Scale score < 6, have an available study partner with at least 10 hours contact per week and who is able to accompany study visits, have visual and auditory acuity adequate for neuropsychological testing, be in good health with no diseases precluding enrollment, be willing to participate in a longitudinal imaging study, have six grades education or work history, be fluent in English or Spanish, express commitment to repeated 3T MRI and at least two PET scans, have no contraindications to MRI, agree to blood collection for GWAS, APOE genotyping and DNA banking, agree to blood collection for biomarker testing, agree to at least one lumbar puncture for CSF collection, and not be enrolled in other trials or studies. In addition, women were required to be either sterile or at least two years past childbearing potential. The administration of permitted medications was required to be stable for four weeks.

The aim of ADNI-GO was to newly enroll participants with early MCI, which included individuals who reported a memory complaint (or whose study partner reported one), abnormal memory performance based on education-adjusted cut-offs on the WMS-R delayed recall, an MMSE score ≥ 24, a global CDR score of 0.5 (with a Memory Box score of 0.5), and general cognitive and functional performance insufficient for a diagnosis of DAT by the site physician.

*ADNI-2*

In ADNI-2 (NCT01231971, see https://clinicaltrials.gov), participants were required to be between 55 and 90 years of age, in good general health, and free of conditions expected to interfere with study participation. They had to have a Geriatric Depression Scale score <6, a Hachinski Ischemic Score ≤4, at least six years of education or equivalent work history, and sufficient visual and auditory acuity for neuropsychological testing. Eligible participants needed to be fluent in English or Spanish, not pregnant or of childbearing potential, and willing to participate in a longitudinal imaging study with repeated 3T MRI and PET scans. A reliable study partner with frequent contact (≥10 hours per week) and availability for clinic visits was required. Participants also agreed to provide blood samples for genetic and biomarker testing, as well as at least one lumbar puncture for CSF collection.

Participants in ADNI-2 were stratified into five diagnostic groups: cognitively unimpaired (CU), significant memory concern (SMC), early MCI, late MCI, and mild DAT. CU participants were required to be free of memory complaints, as verified by a study partner. They had to demonstrate normal memory function by scoring above education-adjusted cutoffs on the Logical Memory II subscale of the WMS-R, and achieve an MMSE score ≥ 24. Global CDR scores had to be 0 with a Memory Box score of 0, and participants could not exhibit significant impairment in cognition or activities of daily living. The SMC group met the same criteria as the CU group, but also required a significant subjective memory concern reported by the participant, study partner, or clinician, confirmed by a Cognitive Change Index score ≥16. Early MCI participants were required to report a subjective memory concern and demonstrate abnormal memory performance within a prespecified intermediate range on the Logical Memory II subscale. They also needed an MMSE score ≥ 24 and a global CDR of 0.5 with a Memory Box score of at least 0.5. General cognition and daily functioning had to be sufficiently preserved to preclude a DAT diagnosis by the site physician. Late MCI participants met the same general requirements as early MCI participants but demonstrated greater memory impairment based on Logical Memory II scores. DAT participants had to meet the NINCDS-ADRDA criteria for probable AD. They were required to have a memory complaint, impaired memory function on the Logical Memory II subscale (same as late MCI), an MMSE score between 20 and 26, and a global CDR of 0.5 or 1.0.

Participants were excluded if they had a significant neurologic disease or structural brain abnormality other than AD that could explain cognitive impairment. Excluded conditions included Parkinson's disease, multi-infarct dementia, Huntington's disease, normal pressure hydrocephalus, brain tumor, progressive supranuclear palsy, seizure disorders, subdural hematoma, multiple sclerosis, or significant head trauma with persistent deficits. For the CU and SMC groups, any such condition was exclusionary; for both MCI groups, suspected incipient Alzheimer's disease was permissible; and for the DAT group, neurologic diseases other than AD were exclusionary. In addition, participants were excluded if screening MRI revealed evidence of infection, infarction, or other focal lesions, including multiple lacunes or lacunes in critical memory structures, or if they had contraindications to MRI (e.g., pacemakers, aneurysm clips, metal implants). Major psychiatric illness was exclusionary, including major depression or bipolar disorder within the past year, schizophrenia, or recent psychotic features, agitation, or behavioral disturbances that could impair compliance. Current treatment for obsessive-compulsive disorder or attention deficit disorder, as well as a history of alcohol or substance abuse within the past two years, were also exclusionary. Additional exclusion criteria included unstable systemic illness, clinically significant abnormalities in vitamin B12 or thyroid function tests, residence in a skilled nursing facility, and current use of certain psychoactive medications, anticoagulants (warfarin or dabigatran, relevant to lumbar puncture), or other exclusionary drugs. Use of investigational agents within one month of enrollment or concurrent participation in other clinical studies involving repeated neuropsychological testing was not permitted. Finally, participants were excluded if prior or ongoing exposure to radioactive agents would exceed regulatory safety limits for PET imaging procedures.

*ADNI-3*

For inclusion in ADNI-3 (NCT02854033, see https://clinicaltrials.gov), participants had to be 55–90 years old, in good general health, and free of conditions likely to interfere with the study. Additional requirements included a Geriatric Depression Scale score <6, adequate vision and hearing for neuropsychological testing, and the availability of a study partner with frequent contact (≥10 hours per week) who could accompany the participant to all study visits. Women had to be post-menopausal for at least two years or surgically sterile. Participants had to be willing and able to take part in a longitudinal imaging study, including repeated 3T MRI and at least two PET scans, and to undergo at least one lumbar puncture. Finally, they were required to provide blood samples for genomic testing (including APOE genotyping and GWAS), biomarker analyses, and biospecimen banking, with consent to share resulting data and samples.

Participants in ADNI-3 were stratified into three diagnostic groups: cognitively unimpaired (CU), MCI, and mild DAT. CU participants could be participants with or without subjective memory complaints, provided memory performance was within the prespecified normal range on the Logical Memory II subscale. They were required to score ≥ 24 on the MMSE and have a global CDR score of 0 with a Memory Box score of 0. MCI participants were required to report a subjective memory concern, either self-reported or recalled by a study partner or clinician, and demonstrate abnormal memory performance below prespecified cutoffs on the Logical Memory II. They also needed an MMSE score between 24 and 30 and a global CDR of 0.5 with a Memory Box score ≥0.5. General cognition and functional performance had to be sufficiently preserved so that a diagnosis of dementia could not be made by a site physician. Stable treatment with cholinesterase inhibitors or memantine was permitted if ongoing for ≥12 weeks prior to screening. DAT participants were required to report a memory concern (either self-reported or recalled by a partner or clinician), demonstrate abnormal memory performance in the impaired range on the Logical Memory II subscale, and score between 20 and 26 on the MMSE. A global CDR score of 0.5 or 1.0 was required, and participants had to meet NINCDS-ADRDA criteria for probable AD.

Participants were excluded if baseline MRI showed focal lesions, multiple lacunes, or lacunes in critical memory structures, or if they had contraindications to MRI (e.g., pacemakers, metal implants, foreign objects in the body). Major psychiatric illness was exclusionary, including major depression or bipolar disorder within the past year, psychosis, agitation or behavioral problems within the past three months, current treatment for obsessive-compulsive disorder or attention-deficit/hyperactivity disorder, or a history of schizophrenia. Substance abuse or dependence within the past two years also led to exclusion. Additional criteria included unstable systemic illness, clinically significant abnormalities in vitamin B12 or thyroid function tests, or residence in a skilled nursing facility. Participants were excluded if they were taking psychoactive medications (e.g., certain antidepressants, neuroleptics, chronic anxiolytics, sedative hypnotics), anticoagulants (warfarin, dabigatran, rivaroxaban, apixaban), or other exclusionary drugs. Use of investigational agents within one month of entry, or concurrent participation in other studies involving repeated neuropsychological testing, was not permitted. For the CU group, the presence of any significant neurologic disease was exclusionary, including Parkinson's disease, multi-infarct dementia, Huntington's disease, normal pressure hydrocephalus, brain tumor, progressive supranuclear palsy, seizure disorder, subdural hematoma, multiple sclerosis, or a history of significant head trauma with persistent deficits or structural brain abnormalities. For MCI group, the same neurologic conditions were exclusionary, with the exception that suspected incipient AD was permissible. For the DAT group, all significant neurologic conditions other than AD itself were exclusionary.

***ARWIBO***

Data used in the preparation of this article were obtained from the Alzheimer's Disease Repository Without Borders (ARWIBO) (https://www.arwibo.it). The primary aim of ARWIBO is to publish all clinical, neuropsychological, EEG, neuroimaging, and biological data of patients with neurodegenerative diseases and healthy controls collected in over 10 years by a number of researchers of IRCCS Fatebenefratelli, Brescia, Italy.[2,3] The overall goal of ARWIBO is to contribute, thorough synergy with neuGRID (https://neugrid2.eu), to global data sharing and analysis in order to develop effective therapies, prevention methods and a cure for Alzheimer' and other neurodegenerative diseases.

Based on the predefined threshold requiring at least 15 scans per scanner manufacturer within each sample, we excluded scans acquired on 1.5T Philips ($n$ = 12) and Siemens ($n$ = 9) systems.

***DELCODE***

The DELCODE study (DRKS00007966, see German Clinical Trial Registry [DRKS]) included two control groups and three patient groups. Clinical screening of the patient groups (subjective cognitive decline [SCD], MCI, and DAT) was performed at the respective memory clinics.

Participants in the SCD group met research criteria for SCD, reported concerns about a subjectively perceived cognitive decline to a physician, and were cognitively unimpaired, defined as scoring better than −1.5 *SD* below German age-, sex-, and education-adjusted norms on the Consortium to Establish a Registry for Alzheimer's Disease (CERAD) test battery. The MCI group met research criteria for amnestic MCI, defined by performance below −1.5 *SD* on the delayed recall trial of the CERAD word-list episodic memory test. The DAT group met research criteria for DAT (ref.[4]) and had Mini-Mental State Examination (MMSE) scores ≥ 18. One control group included first-degree relatives of AD patients, and the other comprised unrelated controls. Control participants performed better than −1.5 *SD* below German age-, sex-, and education-adjusted CERAD norms and reported no subjective concerns about cognitive decline. For the present study, these two control groups and the SCD group were combined into a single CU group.

Inclusion criteria for all diagnostic groups were age of 60 years or older, fluency in German, the ability to provide informed consent, and the availability of a study partner. Exclusion criteria included conditions that would prevent participation in the study protocol (e.g., significant sensorimotor impairments), current or past psychiatric disorders (such as major depressive disorder or disorders due to psychoactive substance use), neurodegenerative disorders other than AD, vascular dementia, and a history of stroke with persistent clinical symptoms. The use of sedative, anticholinergic, or other anti-dementia medication, as well as investigational agents for the treatment of cognitive impairment or dementia, up to one month prior to screening and during the study period, led to exclusion in all groups except for the DAT group.

***NACC***

The NACC Uniform Data Set has captured the enrollment of the Alzheimer's Disease Research Center (ADRC) program since 2005 and includes participants across the full cognitive spectrum found in AD. Enrollment procedures differ by ADRC and may involve clinician referral, self-referral by participants or family members, or active community recruitment. Many ADRCs also enroll CU volunteers. In some ADRCs, agreement to brain autopsy is required for participation, which may introduce additional selection biases. Written informed consent is obtained from all participants and co-participants prior to enrollment.

Three NACC-3T participants were excluded due to impossible (e.g., negative) intracranial volume estimates generated by FastSurfer. Based on the predefined threshold requiring at least 15 scans per scanner manufacturer within each sample, we excluded scans acquired on 1.5T Philips ($n$ = 8) and Siemens ($n$ = 12) systems.

**Biomarkers of Aβ burden in each cohort**

In A4/LEARN, Aβ burden was assessed using $^{18}$F-Florbetapir PET. Global neocortical standardized uptake value ratios (SUVRs) were calculated with a cerebellar reference. SUVRs ≥ 1.10 indicated Aβ positivity. In ADNI, Aβ status was determined using the biomarker measure closest to the MRI scan. CSF-based classification relied on the p-tau$_{181}$/Aβ$_{42}$ ratio (cut-off at ≥ 0.025).[5] PET data were obtained using the $^{18}$F-Florbetapir or $^{18}$F-Florbetaben radiotracers, applying established cohort-specific cut-offs for global cortical SUVRs with a cerebellar reference (≥ 1.11 and ≥ 1.08, respectively). In DELCODE-3T, Aβ status was primarily derived from CSF Aβ$_{42}$/Aβ$_{40}$ ratios (≤ 0.08).[6] For participants lacking CSF data, a previously described probabilistic model of CSF-based Aβ positivity that relies on demographic, genetic, and plasma biomarker information was applied.[7] In NACC, Aβ status was determined from Aβ PET, using four different radiotracers and cohort-specific cut-offs for global cortical SUVRs with a cerebellar reference (≥ 1.12 for $^{18}$F-Florbetaben, ≥ 1.17 for $^{18}$F-Florbetapir, ≥ 1.14 for $^{18}$F-NAV4694, ≥ 1.14 for $^{11}$C-Pittsburgh Compound B).

**Neuropsychological data availability and processing**

Only neuropsychological measures with at least 75% baseline availability within a given sample were considered, leading to the exclusion of TMT B–A scores from ARWIBO-1.5T (available in 16.6% of participants). In addition, A4/LEARN-3T lacked TMT B–A and FAQ data, and ARWIBO-1.5T lacked Logical Memory Delayed Recall and FAQ data. In A4/LEARN-3T, 18 Logical Memory Delayed Recall scores closest to MRI acquisition (1.46%) were obtained using a version other than "A." These were excluded from cross-sectional analyses to ensure comparability, while longitudinal analyses controlled for version effects. The availability of cross-sectional neuropsychological test scores is summarized in Supplementary Figure 1.

Visual inspection of the neuropsychological test score distributions revealed pronounced deviations from normality for MMSE, FAQ, and TMT B–A scores across samples. Consequently, these scores were pre-normalized within each sample using latent process modeling implemented in the *lcmm* R package (ref.[8]), following published recommendations.[9] Supplementary Figure 2 shows the distributions of raw and pre-normalized scores. MMSE, Logical Memory Delayed Recall, and TMT B–A scores were $z$-standardized within each sample using the mean and standard deviations ($SD$s) from baseline CU group distributions. Expectedly, FAQ scores showed insufficient variability within CU groups to permit meaningful scaling and were therefore $z$-standardized to the whole samples.

**Markers standardization**

MTL volumes were corrected for total intracranial volume following the regression-based approach as described by Voevodskaya *et al.*[10] To compute $w$-scores, we first evaluated four OLS regression specifications for each marker:

1. age + sex + vendor + field strength (fs)
2. natural spline of age ($df$ = 2) + sex + vendor + fs
3. age + sex + vendor × fs
4. natural spline of age ($df$ = 2) + sex + vendor × fs

For each marker, we fitted the models using all available CU participants and selected the lowest AIC model to derive a fixed formula structure that was applied in all modelling iterations.

#### Outlier removal

All raw MRI measures were screened for numerical outliers. Prior to standardization, observations with total intracranial volume outside ±5 *SD* from the mean, stratified by sample, scanner manufacturer, and participant sex, were excluded, resulting in the following thresholds:

| Sample | Manufacturer | Sex | Mean | *SD* | Lower cut-off | Upper cut-off |
|---|---|---|---|---|---|---|
| A4/LEARN-3T | GE | female | 1486357.75 | 111929.91 | 926708.20 | 2046007.30 |
| | | male | 1330770.88 | 124571.12 | 707915.28 | 1953626.48 |
| | Philips | female | 1609235.98 | 138052.11 | 918975.43 | 2299496.54 |
| | | male | 1418524.11 | 125447.58 | 791286.22 | 2045761.99 |
| | Siemens | female | 1554149.94 | 131489.63 | 896701.80 | 2211598.08 |
| | | male | 1371582.49 | 128987.94 | 726642.77 | 2016522.20 |
| ADNI-1.5T | GE | female | 1449425.05 | 113637.06 | 881239.73 | 2017610.37 |
| | | male | 1653646.89 | 140077.62 | 953258.78 | 2354035.00 |
| | Philips | female | 1471159.39 | 100506.11 | 968628.85 | 1973689.92 |
| | | male | 1629031.74 | 137395.51 | 942054.18 | 2316009.30 |
| | Siemens | female | 1421330.09 | 113044.38 | 856108.18 | 1986552.00 |
| | | male | 1650845.04 | 161012.43 | 845782.88 | 2455907.19 |
| ADNI-3T | GE | female | 1361090.23 | 167716.63 | 522507.07 | 2199673.39 |
| | | male | 1480231.80 | 134187.87 | 809292.45 | 2151171.14 |
| | Philips | female | 1406654.51 | 130884.42 | 752232.41 | 2061076.60 |
| | | male | 1629318.76 | 129194.25 | 983347.50 | 2275290.02 |
| | Siemens | female | 1386173.75 | 130765.05 | 732348.49 | 2039999.00 |
| | | male | 1547612.64 | 140756.52 | 843830.06 | 2251395.23 |
| ARWIBO-1.5T | GE | female | 1303548.35 | 111819.44 | 744451.17 | 1862645.54 |
| | | male | 1448996.50 | 165751.44 | 620239.29 | 2277753.71 |
| DELCODE-3T | Siemens | female | 1512815.57 | 113543.84 | 945096.37 | 2080534.77 |
| | | male | 1735791.64 | 141958.26 | 1026000.32 | 2445582.96 |
| NACC-1.5T | GE | female | 1391967.43 | 128848.69 | 747723.97 | 2036210.88 |
| | | male | 1577128.73 | 160958.77 | 772334.89 | 2381922.57 |
| NACC-3T | GE | female | 1304766.89 | 247761.04 | 65961.69 | 2543572.09 |
| | | male | 1474749.02 | 189224.97 | 528624.16 | 2420873.89 |
| | Philips | female | 1383175.17 | 127510.65 | 745621.93 | 2020728.42 |
| | | male | 1596260.62 | 157548.90 | 808516.13 | 2384005.10 |
| | Siemens | female | 1384425.76 | 132880.84 | 720021.56 | 2048829.96 |
| | | male | 1574795.63 | 150677.88 | 821406.22 | 2328185.03 |

This led to the exclusion of $n = 1$ in ADNI-3T, $n = 1$ in ARWIBO-1.5T, $n = 1$ in DELCODE-3T, and $n = 6$ in NACC-3T.

After standardization, observations were further excluded if any *w*-scored atrophy marker lay outside $-4 < w < 4$ for CU participants or $-7 < w < 7$ for MCI and DAT participants. This resulted in the exclusion of participants as follows: ADNI-1.5T ($n = 1$), ARWIBO-1.5T ($n = 1$), A4/LEARN-3T ($n = 3$), ADNI-3T ($n = 3$), NACC-1.5T ($n = 3$), and NACC-3T ($n = 19$).

### Supplementary figures

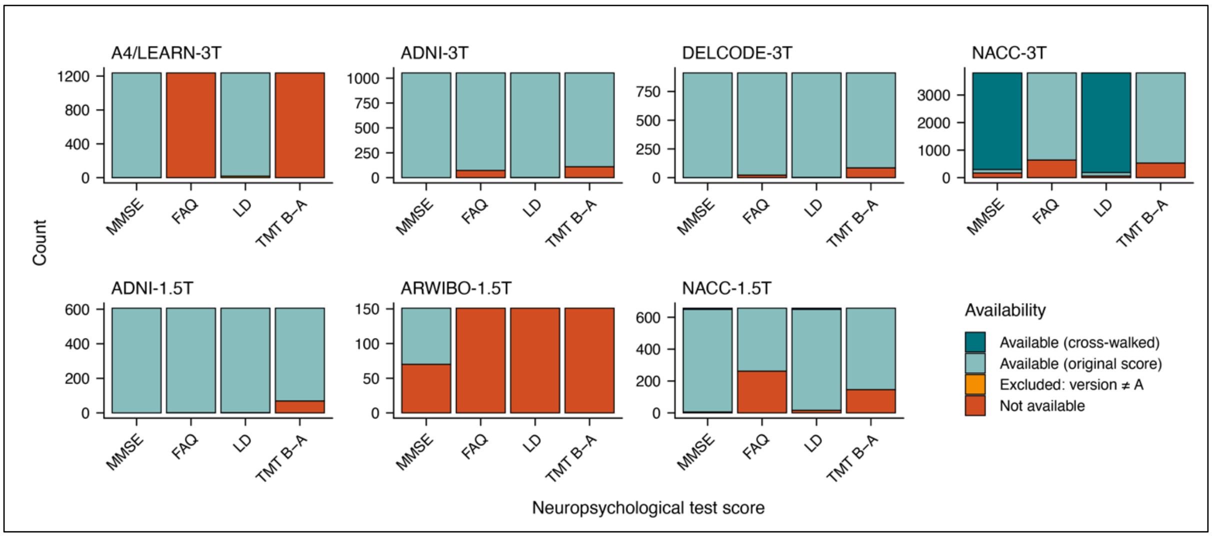


**Supplementary Figure 1 Data availability in the study sample.**

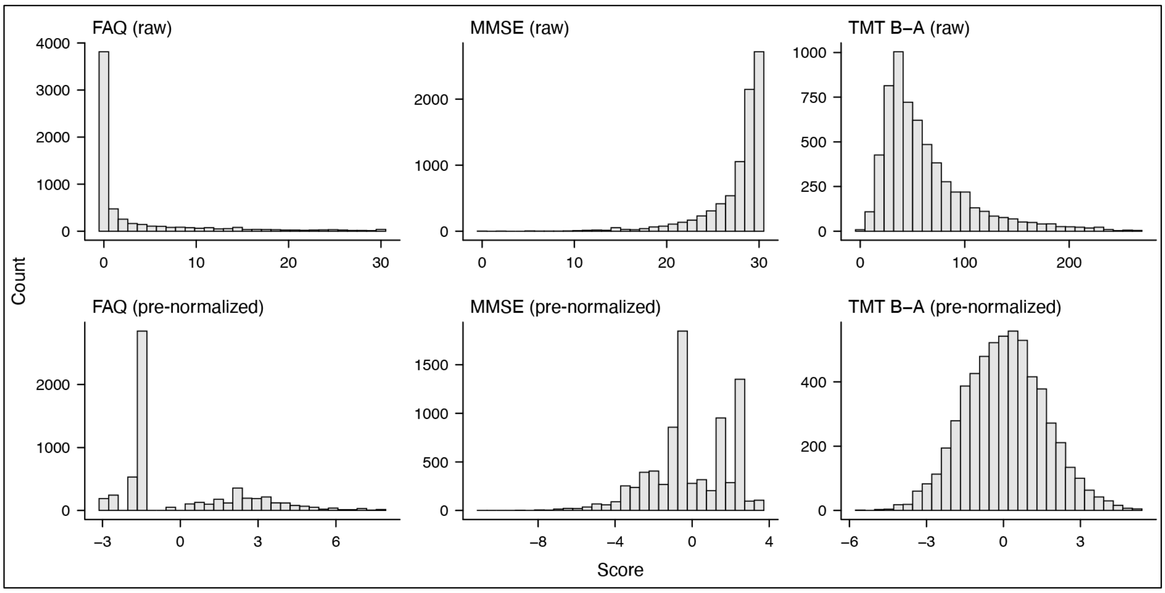


**Supplementary Figure 2 Distributions of FAQ, MMSE, and TMT B–A scores before (top row) and after (bottom row) pre-normalization.**

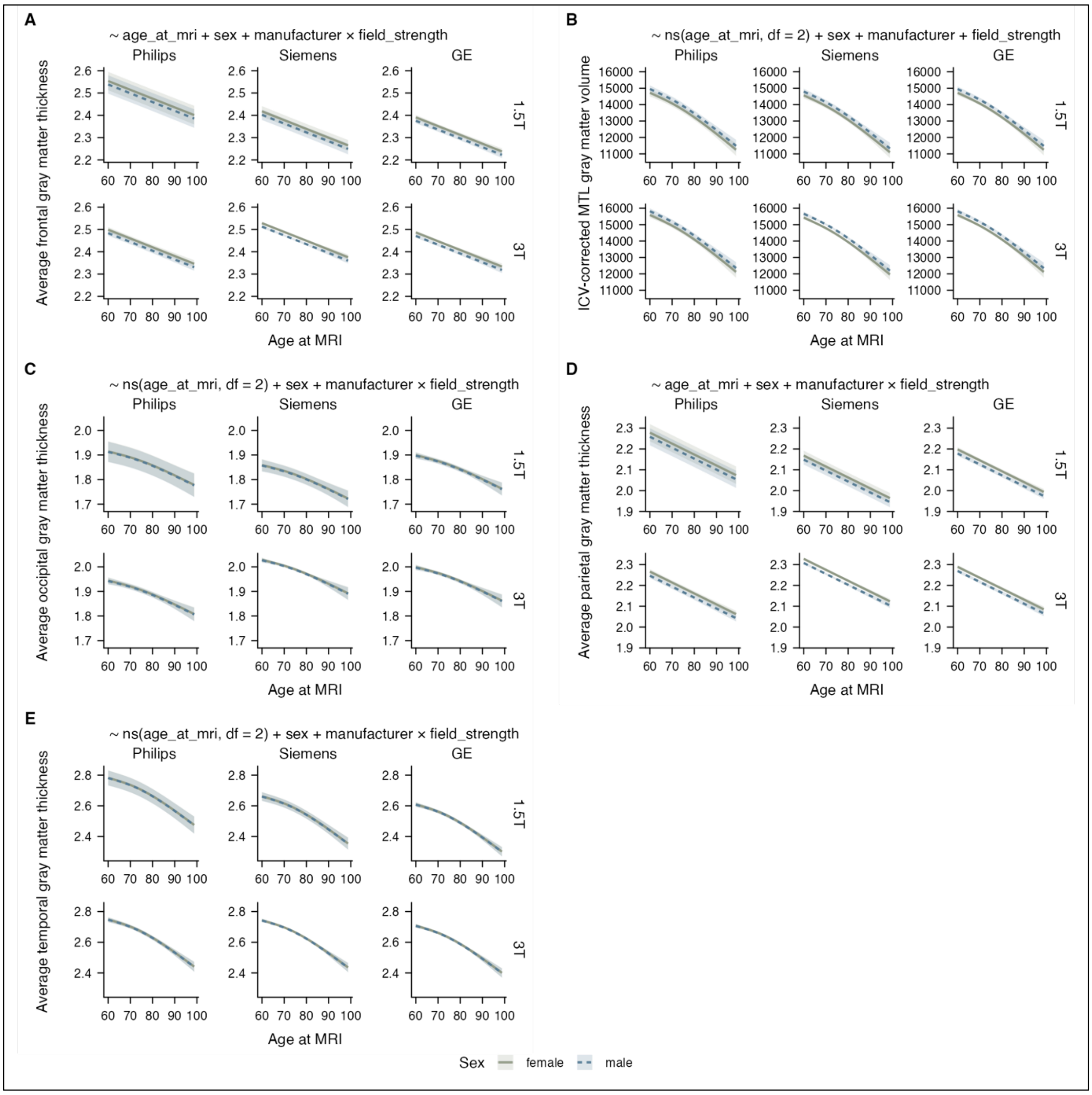


**Supplementary Figure 3 Modeled relationships between age and MRI markers in CU participants, stratified by scanner manufacturer, magnetic field strength, and participant sex.** The optimal model used for visualization and marker standardization is indicated above each respective plot. Ribbons depict 95% C.I.s. Abbreviations: ICV, intracranial volume.

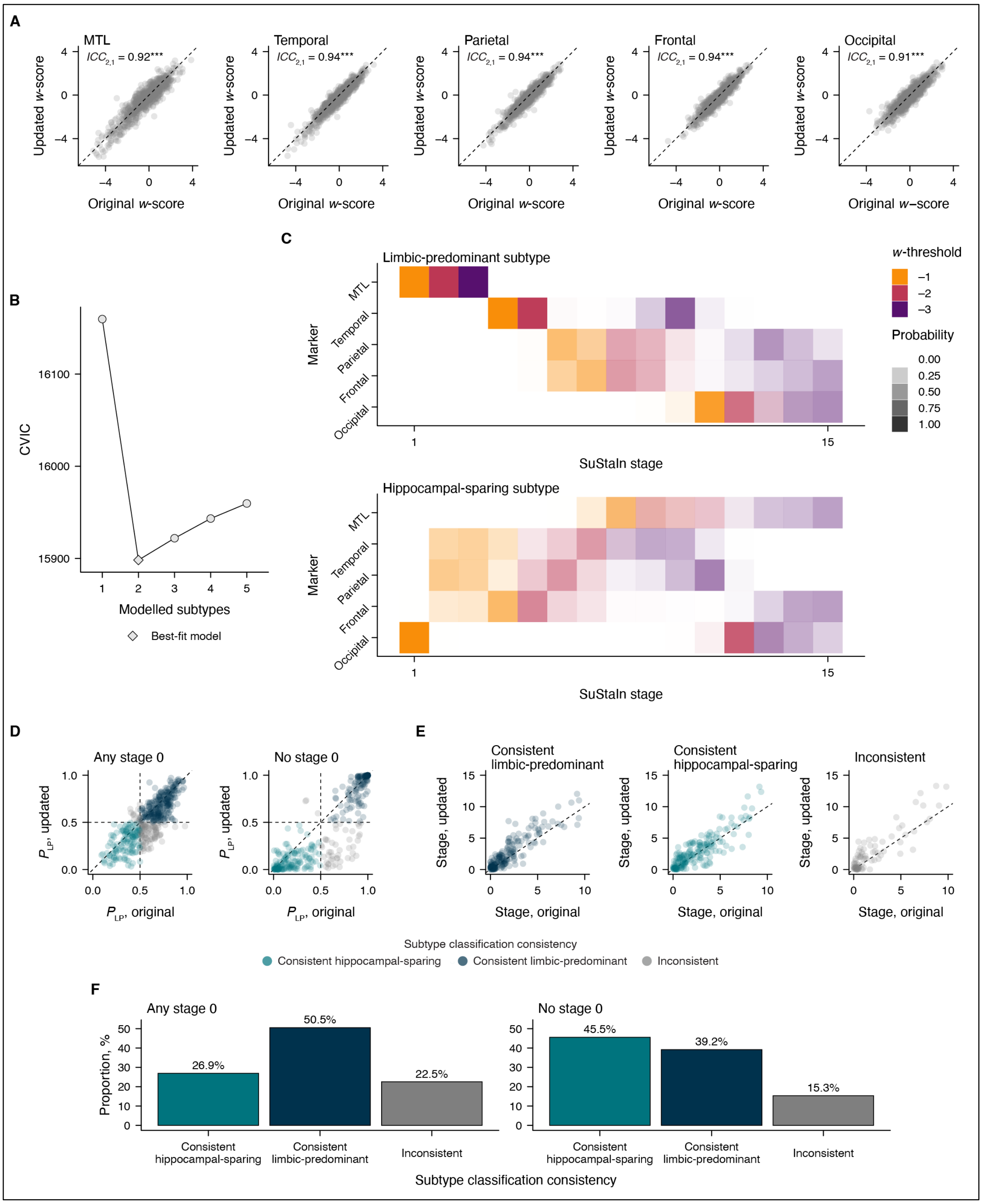


**Supplementary Figure 4. Replication of the original subtyping and staging results in DELCODE-3T using updated methods for faster and more accessible atrophy subtyping and staging.** (**A**) Scatter plots comparing the original and updated *w*-scored atrophy markers used for SuStaIn modeling. The original model based on these markers was published in Baumeister *et al.*[11] (**B**) Cross-validation information criterion (CVIC) from ten-fold cross-validation used to identify the most likely number of atrophy subtypes within the DELCODE-3T sample. (**C**) Positional variance diagram visualizing the optimal two-subtype model. (**D**, **E**) Scatter plots showing individual probabilities of limbic-predominant atrophy ($P_{LP}$) and atrophy stage as estimated by the original and updated models. The "Any stage 0"panel includes observations classified as stage 0 by at least one of the two models, whereas the "No stage 0" panel includes only those not classified as stage 0 by either model. (**F**) Proportions of consistent and inconsistent subtype classifications between the two models. All axes representing atrophy stage were jittered by up to ± 0.5 stages to improve visual clarity.

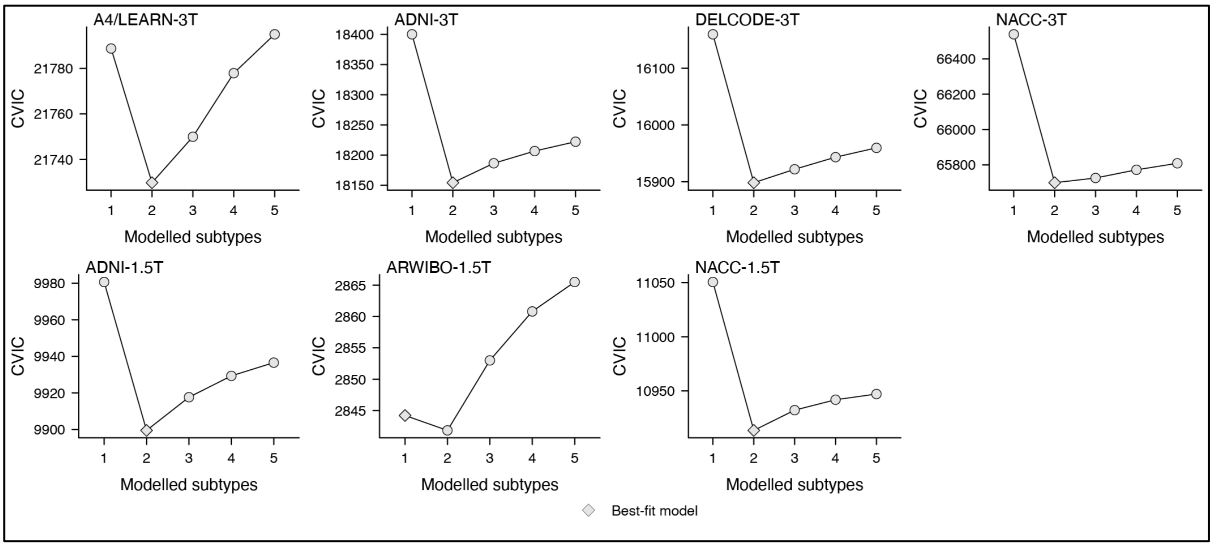


**Supplementary Figure 5 Model selection criteria derived from within-sample SuStaIn modeling, including all available data.**

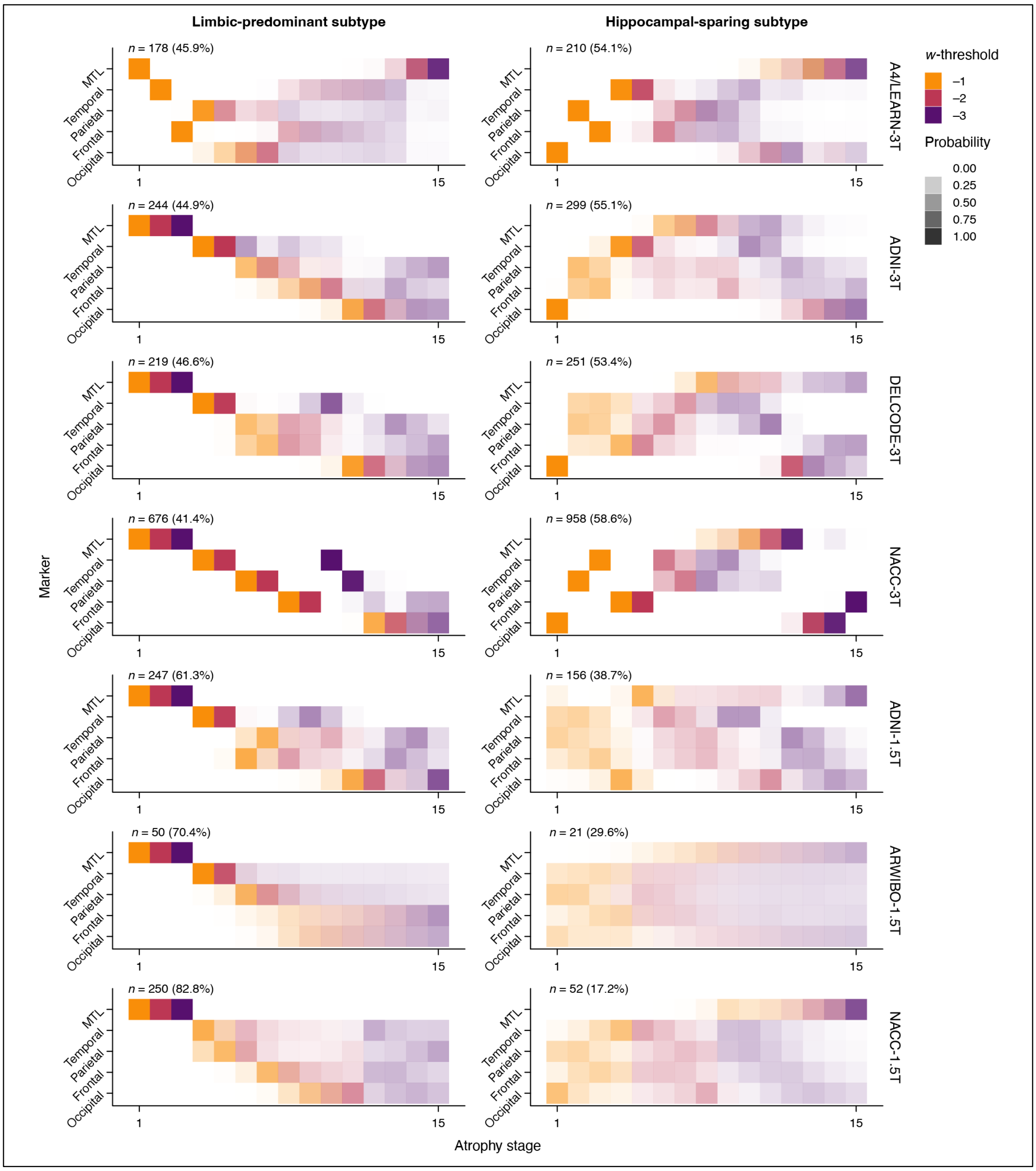


**Supplementary Figure 6 Positional variance plots showing the atrophy progression sequences identified from independent, within-sample modeling using all available data.** The proportions shown above each plot refer to participants with an assigned atrophy stage > 0.

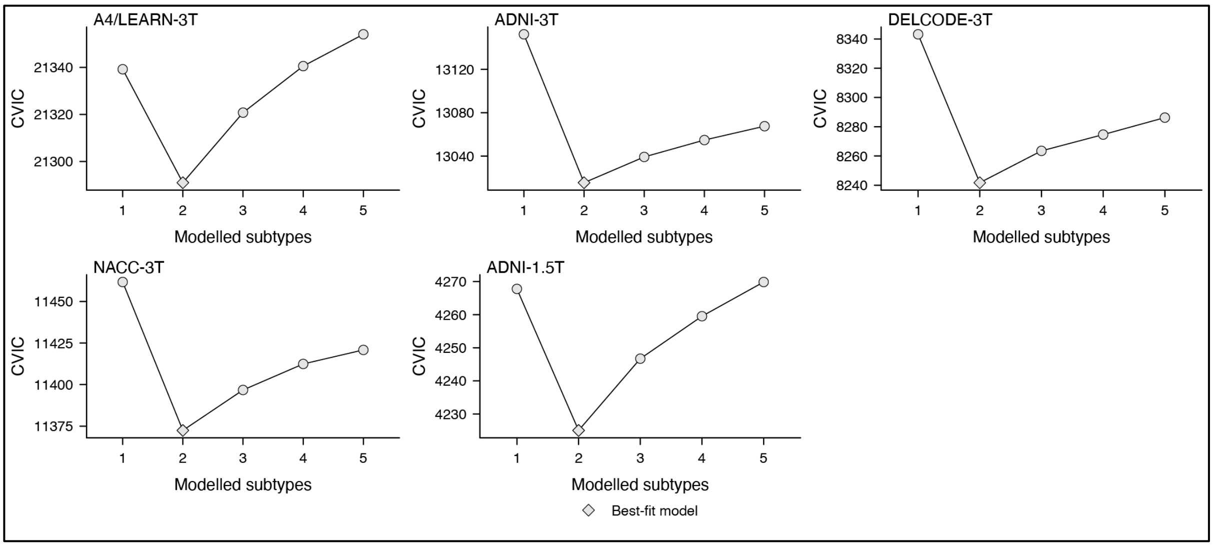


**Supplementary Figure 7 Model selection criteria derived from within-sample SuStaIn modeling, including only the Aβ-positive subsets.**

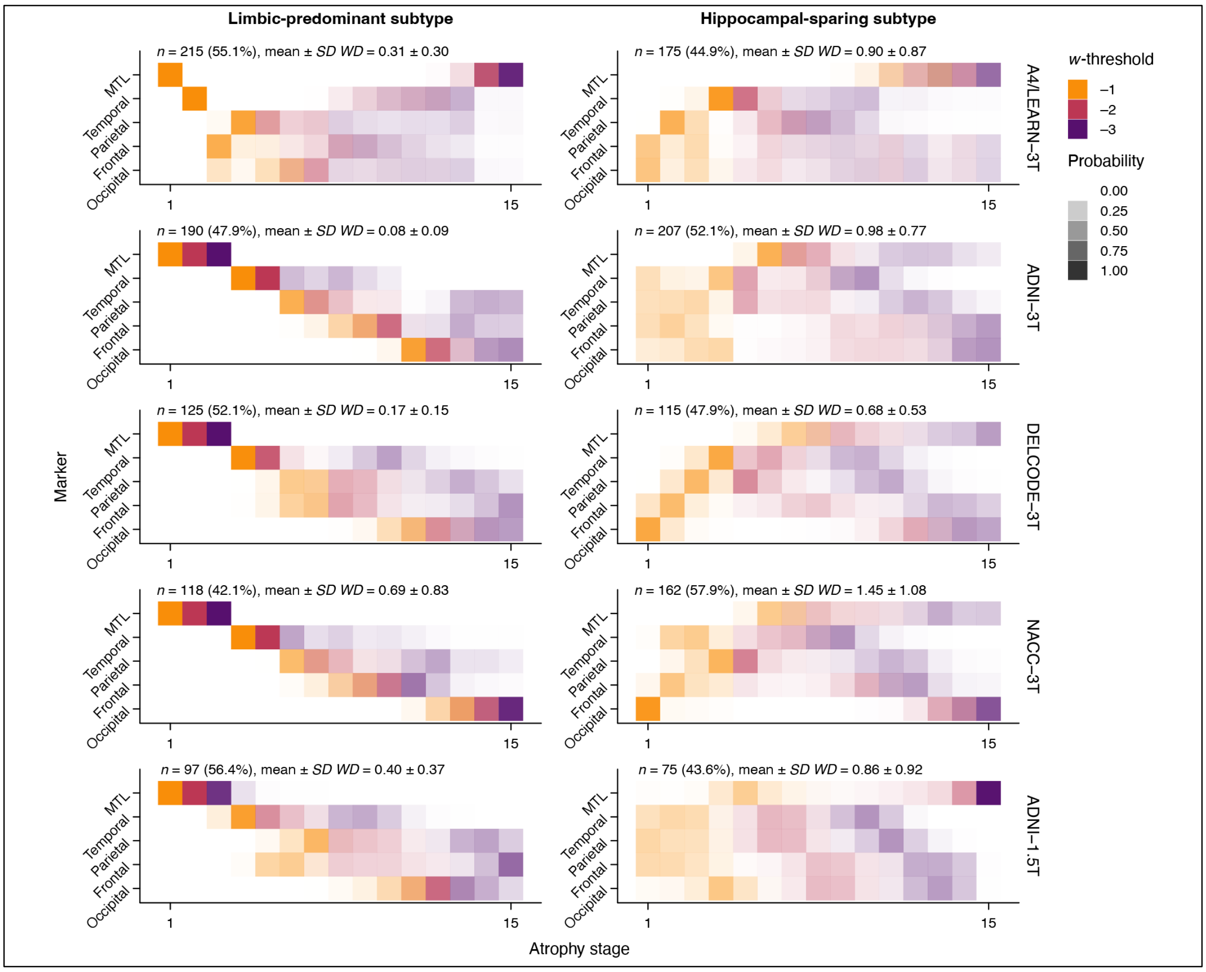


**Supplementary Figure 8 Positional variance plots showing the atrophy progression sequences identified from independent, within-sample modeling using Aβ-positive subsets.** The proportions shown above each plot refer to participants with an assigned atrophy stage > 0. *WD* was used to quantify the stage shift between sequences derived from Aβ-positive sub-samples and those from the full sample. The proportions shown above each plot refer to participants with an assigned atrophy stage > 0.

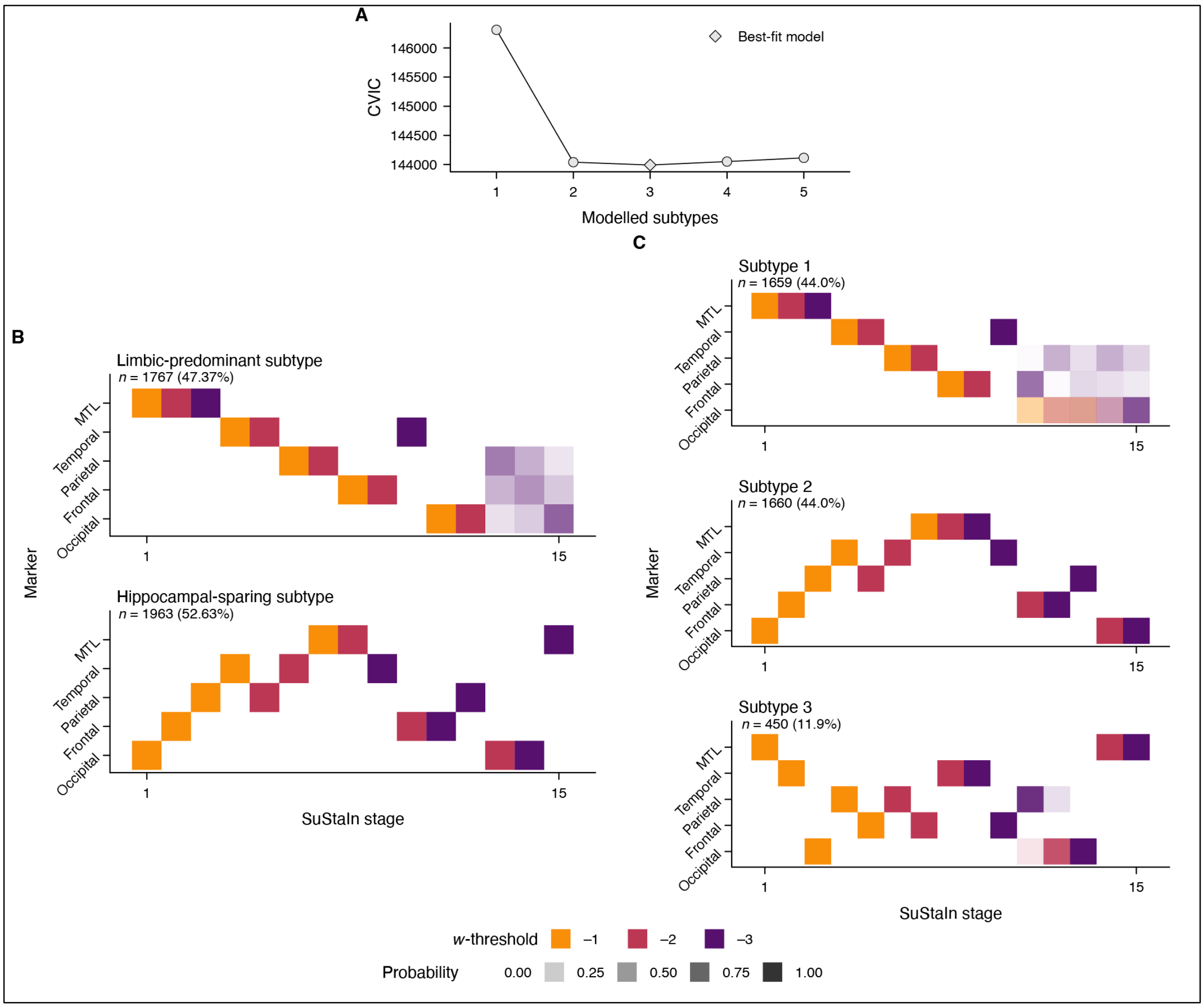


**Supplementary Figure 9 Results from atrophy subtyping and staging as part of PHASE–AD.** (**A**) Cross-validation information criterion (CVIC) from ten-fold cross-validation used to identify the most likely number of atrophy subtypes. (**B**) Positional variance diagram visualizing the two-subtype model. (**C**) Positional variance diagram visualizing the three-subtype model. The proportions shown above each plot in (***B***, **C**) refer to participants with an assigned atrophy stage > 0.

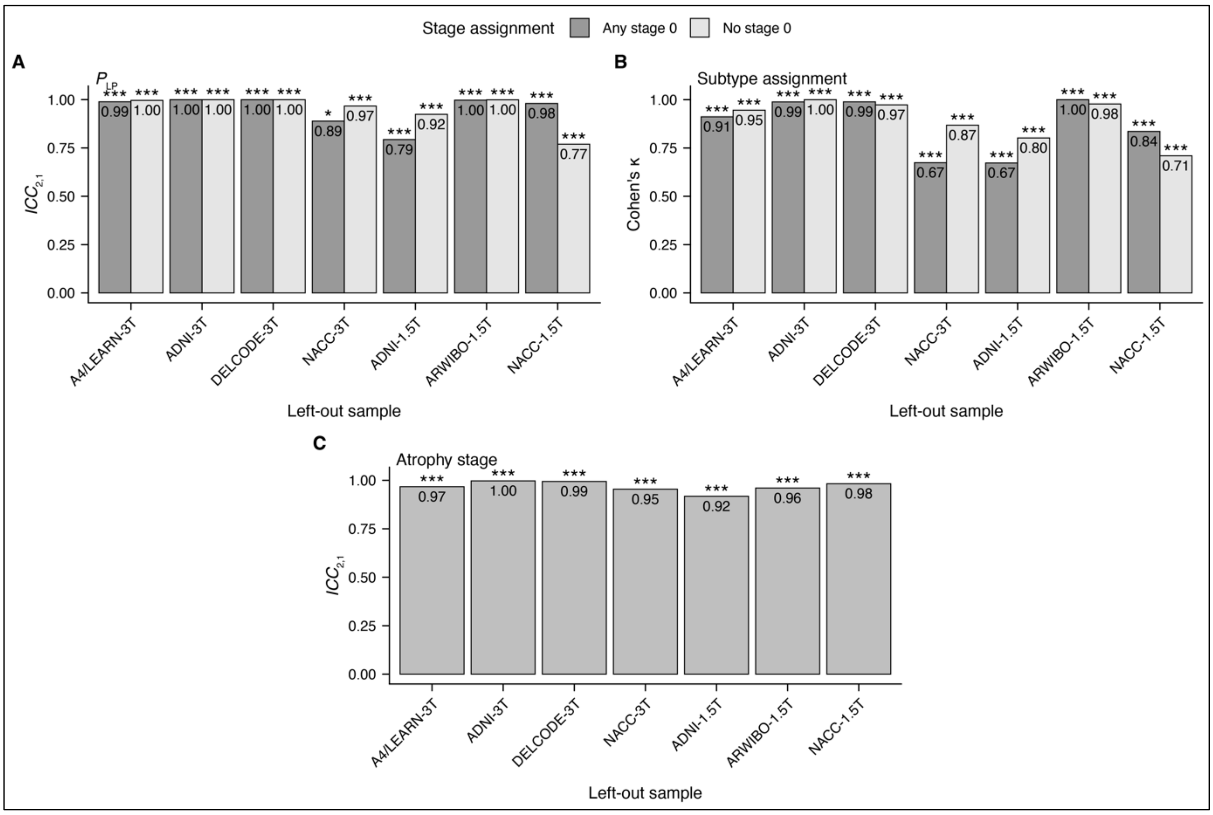


**Supplementary Figure 10 Robustness of PHASE–AD classifications to sample removal.** Leave-one-sample-out (LOSO) cross-validation results are shown for (**A**) probabilistic likelihood of progression ($P_{LP}$, $ICC_{2,1}$), (**B**) subtype assignment (Cohen's κ), and (**C**) atrophy stage ($ICC_{2,1}$). In (**A**,**B**), dark gray bars indicate agreement between observed and predicted classifications for participants assigned to stage 0 by either model, while light gray bars indicate agreement for all other participants. Significance levels: *$p$ < .05, **$p$ < .01, ***$p$ < .001

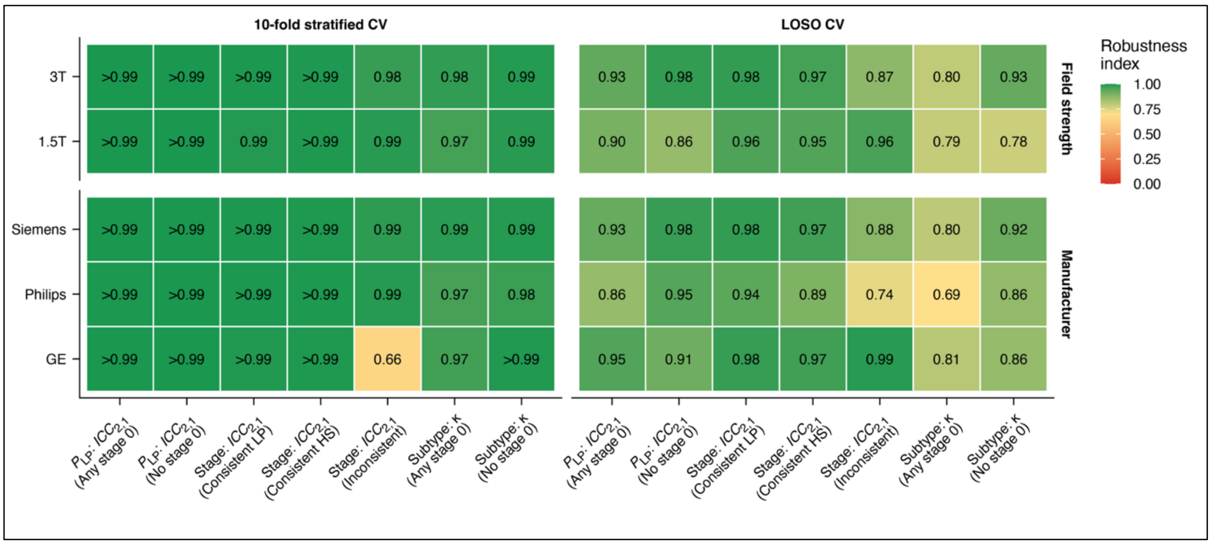


**Supplementary Figure 11 Robustness of PHASE–AD classifications across scanner manufacturers and field strengths.** Heatmap of robustness indices for PHASE–AD classifications stratified by scanner manufacturer and MRI field strength, shown separately for the applied 10-fold stratified and LOSO cross-validation schemes. Rows correspond to stratification groups, columns correspond to robustness metrics. Abbreviations: HS, hippocampal-sparing. LP, limbic-predominant.

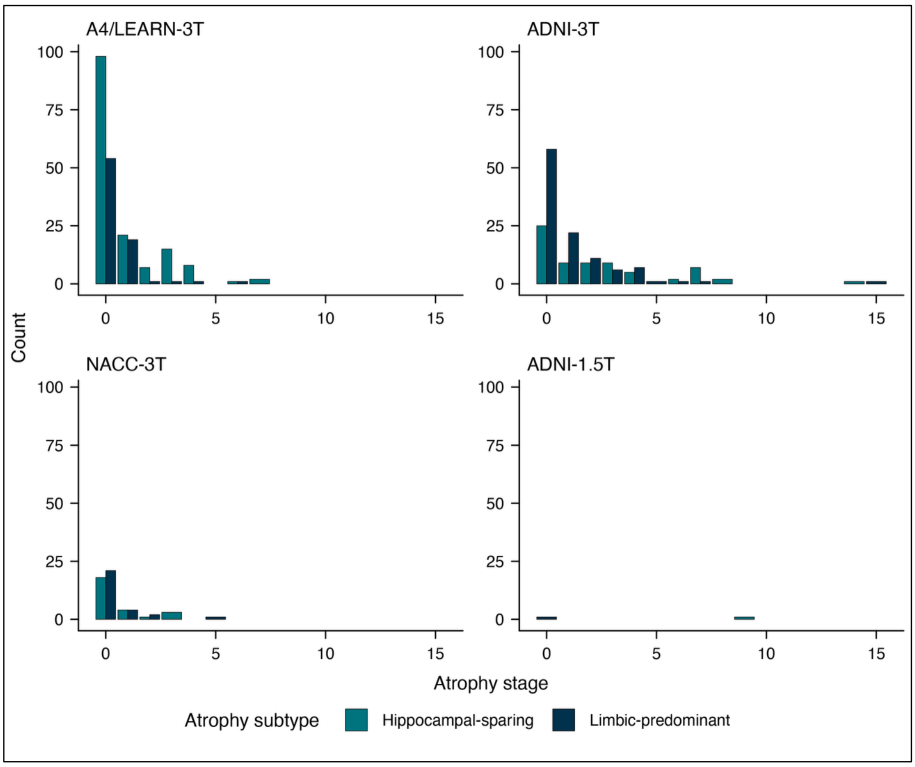


**Supplementary Figure 12 Distribution of atrophy subtypes and stages across samples in the tau PET sample.** Bar plots show the number of participants at each atrophy stage, stratified by subtype and sample. Only Aβ-positive individuals with available tau PET data are included.

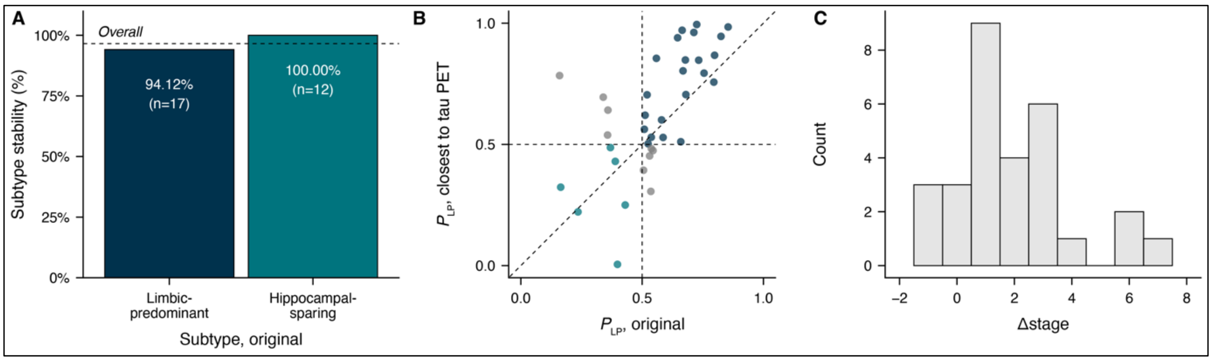


**Supplementary Figure 13 Stability of atrophy subtype and stage classifications across longitudinal MRI acquisitions in the tau PET analysis sample.** (**A**) Proportion of individuals with stable subtype classification between the original MRI and the MRI closest to tau PET, stratified by baseline subtype. The dashed line indicates overall stability across subtypes. (**B**) Agreement of limbic-predominant subtype probabilities ($P_{LP}$) between timepoints for individuals initially classified as stage 0. Points are colored by consistency of subtype assignment. (**C**) Distribution of change in atrophy stage (Δstage) between timepoints among individuals with baseline stage > 0. Negative values indicate violations of stage monotonicity. Participants classified as stage 0 at baseline were excluded, as stage decreases (violations of monotonicity) cannot occur from this baseline level.

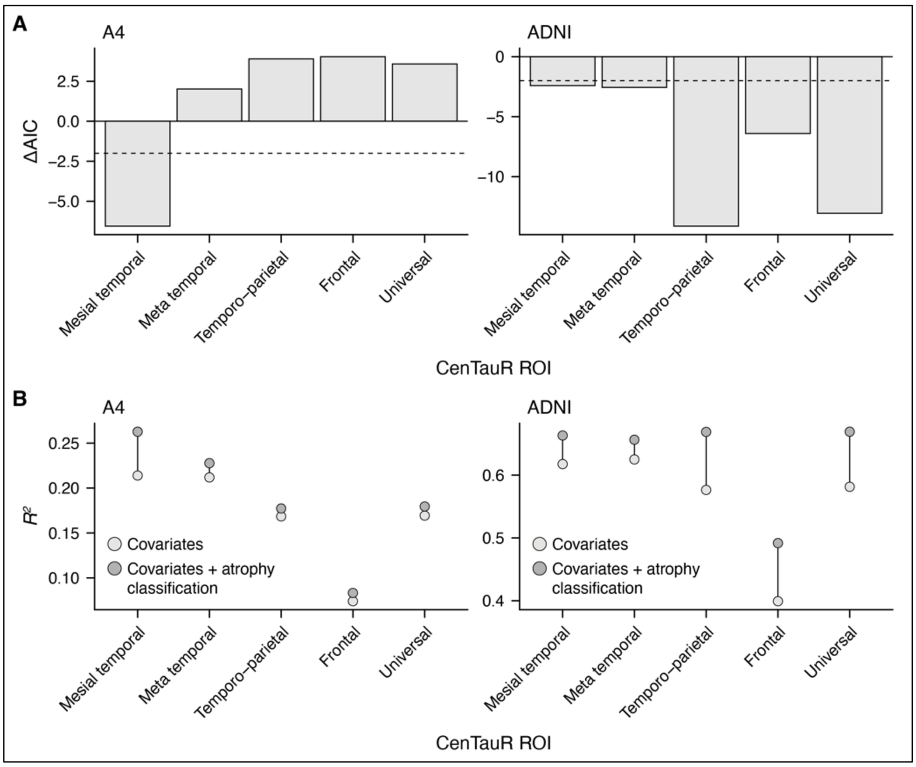


**Supplementary Figure 14 Associations between atrophy stage, subtype, and regional tau burden using plasma p-tau as covariate in Aβ-positive participants.** (**A**) Model comparison across CenTauR ROIs, showing ΔAIC for full models relative to covariates-only models. The dashed line indicates ΔAIC = −2, prespecified as evidence of improved model fit. (**B**) Total variance explained ($R^2$) for covariates-only and full models across CenTauR ROIs. Results are shown separately for A4 and ADNI samples.

### Supplementary tables

**Supplementary Table 1 FastSurfer cortical labels grouped into meta-regions used for subsequent analyses.**

| Meta-region | Included labels |
|---|---|
| MTL | entorhinal (from <lh/rh>.BA_exvivo.stats), hippocampus, amygdala (both from aseg+DKT.stats) |
| Temporal | inferiortemporal, fusiform, bankssts, middletemporal, temporalpole, superiortemporal, transversetemporal |
| Parietal | isthmuscingulate, inferiorparietal, precuneus, superiorparietal, supramarginal, posteriorcingulate, postcentral |
| Occipital | lateraloccipital, lingual, pericalcarine, cuneus |
| Frontal | caudalanteriorcingulate, caudalmiddlefrontal, lateralorbitofrontal, medialorbitofrontal, parsopercularis, parsorbitalis, paracentral, parstriangularis, precentral, rostralanteriorcingulate, rostralmiddlefrontal, superiorfrontal, insula, frontalpoles |

**Supplementary Table 2 Summary statistics of cross-sectional subtype effects derived from linear regression models predicting numeric outcome variables.**

| Sample | Sample | Subtype 1 (*n*) | Subtype 2 (*n*) | *b* | *SE* (*b*) | Cohen's *d* | *SE* (*d*) | 95% C.I. (*d*) | $p_{Tukey}$ | $\tau^2$ | $I^2$ | Pooled $p_{raw}$ | Pooled $p_{FDR}$ |
|---|---|---|---|---|---|---|---|---|---|---|---|---|---|
| **Outcome: age (years)** | | | | | | | | | | | | | |
| Pooled | Whole sample | HS (1963) | A– (4685) | | | 0.02 | 0.07 | [–0.12; 0.16] | | 0.03 | 81.44 | .763 | .842 |
| Pooled | Whole sample | LP (1767) | A– (4685) | | | 0.10 | 0.07 | [–0.04; 0.25] | | 0.03 | 75.44 | .168 | .231 |
| Pooled | Whole sample | LP (1767) | HS (1963) | | | 0.07 | 0.12 | [–0.17; 0.32] | | 0.09 | 90.60 | .553 | .676 |
| Pooled | Aβ-positive | HS (525) | A– (893) | | | –0.10 | 0.10 | [–0.29; 0.09] | | 0.03 | 58.68 | .293 | .410 |
| Pooled | Aβ-positive | LP (518) | A– (893) | | | 0.11 | 0.11 | [–0.10; 0.32] | | 0.03 | 57.34 | .302 | .410 |
| Pooled | Aβ-positive | LP (518) | HS (525) | | | 0.21 | 0.13 | [–0.05; 0.47] | | 0.06 | 72.95 | .107 | .161 |
| A4/LEARN-3T | Whole sample | LP (81) | HS (347) | 1.67 | 0.57 | 0.36 | 0.12 | [0.12; 0.61] | .009** | | | | |
| A4/LEARN-3T | Whole sample | LP (81) | A– (809) | 1.82 | 0.53 | 0.40 | 0.12 | [0.17; 0.63] | .002** | | | | |
| A4/LEARN-3T | Whole sample | HS (347) | A– (809) | 0.16 | 0.29 | 0.03 | 0.06 | [–0.09; 0.16] | .851 | | | | |
| A4/LEARN-3T | Aβ-positive | LP (56) | HS (191) | 1.81 | 0.72 | 0.38 | 0.15 | [0.08; 0.68] | .033* | | | | |
| A4/LEARN-3T | Aβ-positive | LP (56) | A– (458) | 2.16 | 0.67 | 0.46 | 0.14 | [0.18; 0.74] | .004** | | | | |
| A4/LEARN-3T | Aβ-positive | HS (191) | A– (458) | 0.35 | 0.41 | 0.08 | 0.09 | [–0.09; 0.24] | .658 | | | | |
| ADNI-3T | Whole sample | LP (225) | HS (289) | 0.65 | 0.64 | 0.10 | 0.09 | [–0.09; 0.28] | .565 | | | | |
| ADNI-3T | Whole sample | LP (225) | A– (538) | 1.22 | 0.62 | 0.18 | 0.09 | [0.00; 0.36] | .117 | | | | |
| ADNI-3T | Whole sample | HS (289) | A– (538) | 0.57 | 0.51 | 0.08 | 0.07 | [–0.06; 0.23] | .494 | | | | |
| ADNI-3T | Aβ-positive | LP (145) | HS (143) | 0.88 | 0.81 | 0.13 | 0.12 | [–0.11; 0.37] | .526 | | | | |
| ADNI-3T | Aβ-positive | LP (145) | A– (162) | 0.54 | 0.87 | 0.08 | 0.13 | [–0.17; 0.33] | .807 | | | | |
| ADNI-3T | Aβ-positive | HS (143) | A– (162) | –0.34 | 0.80 | –0.05 | 0.12 | [–0.29; 0.18] | .906 | | | | |
| DELCODE-3T | Whole sample | LP (265) | HS (254) | 2.55 | 0.55 | 0.44 | 0.09 | [0.25; 0.62] | < .001*** | | | | |
| DELCODE-3T | Whole sample | LP (265) | A– (392) | 1.61 | 0.53 | 0.28 | 0.09 | [0.10; 0.46] | .008** | | | | |
| DELCODE-3T | Whole sample | HS (254) | A– (392) | –0.95 | 0.48 | –0.16 | 0.08 | [–0.32; 0.00] | .115 | | | | |
| DELCODE-3T | Aβ-positive | LP (132) | HS (71) | 2.58 | 0.88 | 0.45 | 0.15 | [0.15; 0.75] | .010* | | | | |
| DELCODE-3T | Aβ-positive | LP (132) | A– (77) | –0.06 | 0.95 | –0.01 | 0.17 | [–0.34; 0.32] | .998 | | | | |
| DELCODE-3T | Aβ-positive | HS (71) | A– (77) | –2.64 | 0.97 | –0.46 | 0.17 | [–0.80; –0.13] | .019* | | | | |
| NACC-3T | Whole sample | LP (784) | HS (698) | 1.62 | 0.38 | 0.23 | 0.05 | [0.13; 0.34] | < .001*** | | | | |
| NACC-3T | Whole sample | LP (784) | A– (2319) | 0.17 | 0.33 | 0.02 | 0.05 | [–0.07; 0.12] | .863 | | | | |
| NACC-3T | Whole sample | HS (698) | A– (2319) | –1.45 | 0.31 | –0.21 | 0.04 | [–0.29; –0.12] | < .001*** | | | | |
| NACC-3T | Aβ-positive | LP (98) | HS (64) | 2.54 | 1.14 | 0.37 | 0.17 | [0.04; 0.71] | .069 | | | | |
| NACC-3T | Aβ-positive | LP (98) | A– (152) | 1.00 | 1.05 | 0.15 | 0.16 | [–0.16; 0.45] | .607 | | | | |
| NACC-3T | Aβ-positive | HS (64) | A– (152) | –1.53 | 1.04 | –0.23 | 0.15 | [–0.53; 0.08] | .303 | | | | |
| ADNI-1.5T | Whole sample | LP (200) | HS (170) | –1.61 | 0.64 | –0.27 | 0.11 | [–0.47; –0.06] | .033* | | | | |
| ADNI-1.5T | Whole sample | LP (200) | A– (236) | –0.39 | 0.67 | –0.06 | 0.11 | [–0.28; 0.15] | .834 | | | | |
| ADNI-1.5T | Whole sample | HS (170) | A– (236) | 1.23 | 0.65 | 0.20 | 0.11 | [–0.01; 0.41] | .145 | | | | |
| ADNI-1.5T | Aβ-positive | LP (87) | HS (56) | –1.76 | 1.02 | –0.30 | 0.17 | [–0.64; 0.04] | .197 | | | | |
| ADNI-1.5T | Aβ-positive | LP (87) | A– (44) | –1.33 | 1.18 | –0.23 | 0.20 | [–0.63; 0.17] | .498 | | | | |
| ADNI-1.5T | Aβ-positive | HS (56) | A– (44) | 0.43 | 1.23 | 0.07 | 0.21 | [–0.34; 0.49] | .935 | | | | |
| ARWIBO-1.5T | Whole sample | LP (54) | HS (38) | 0.37 | 1.44 | 0.06 | 0.24 | [–0.41; 0.53] | .964 | | | | |
| ARWIBO-1.5T | Whole sample | LP (54) | A– (59) | 0.36 | 1.28 | 0.06 | 0.21 | [–0.36; 0.48] | .958 | | | | |
| ARWIBO-1.5T | Whole sample | HS (38) | A– (59) | –0.02 | 1.27 | 0.00 | 0.21 | [–0.42; 0.41] | .999 | | | | |
| NACC-1.5T | Whole sample | LP (158) | HS (167) | –3.15 | 0.85 | –0.43 | 0.12 | [–0.67; –0.20] | < .001*** | | | | |
| NACC-1.5T | Whole sample | LP (158) | A– (332) | –1.22 | 0.82 | –0.17 | 0.11 | [–0.39; 0.05] | .299 | | | | |
| NACC-1.5T | Whole sample | HS (167) | A– (332) | 1.93 | 0.71 | 0.27 | 0.10 | [0.07; 0.46] | .018* | | | | |
| **Outcome: education (years)** | | | | | | | | | | | | | |
| Pooled | Whole sample | HS (1820) | A– (4224) | | | –0.05 | 0.04 | [–0.12; 0.03] | | 0.00 | 34.13 | .215 | .284 |
| Pooled | Whole sample | LP (1646) | A– (4224) | | | 0.00 | 0.03 | [–0.07; 0.06] | | 0.00 | 0.00 | .932 | .932 |
| Pooled | Whole sample | LP (1646) | HS (1820) | | | 0.04 | 0.04 | [–0.03; 0.12] | | 0.00 | 2.57 | .251 | .319 |
| Pooled | Aβ-positive | HS (518) | A– (869) | | | 0.01 | 0.06 | [–0.10; 0.13] | | 0.00 | 0.00 | .839 | .933 |
| Pooled | Aβ-positive | LP (507) | A– (869) | | | 0.01 | 0.08 | [–0.14; 0.16] | | 0.01 | 18.17 | .907 | .972 |
| Pooled | Aβ-positive | LP (507) | HS (518) | | | –0.03 | 0.07 | [–0.17; 0.10] | | 0.00 | 0.00 | .628 | .725 |
| A4/LEARN-3T | Whole sample | LP (81) | HS (347) | 0.21 | 0.34 | 0.08 | 0.12 | [–0.17; 0.32] | .811 | | | | |
| A4/LEARN-3T | Whole sample | LP (81) | A– (809) | 0.05 | 0.32 | 0.02 | 0.12 | [–0.21; 0.25] | .984 | | | | |
| A4/LEARN-3T | Whole sample | HS (347) | A– (809) | –0.15 | 0.18 | –0.06 | 0.06 | [–0.18; 0.07] | .655 | | | | |
| A4/LEARN-3T | Aβ-positive | LP (56) | HS (191) | –0.16 | 0.43 | –0.06 | 0.15 | [–0.36; 0.24] | .924 | | | | |
| A4/LEARN-3T | Aβ-positive | LP (56) | A– (458) | –0.23 | 0.40 | –0.08 | 0.14 | [–0.36; 0.20] | .838 | | | | |
| A4/LEARN-3T | Aβ-positive | HS (191) | A– (458) | –0.06 | 0.24 | –0.02 | 0.09 | [–0.19; 0.15] | .963 | | | | |
| ADNI-3T | Whole sample | LP (220) | HS (281) | –0.09 | 0.24 | –0.04 | 0.09 | [–0.22; 0.15] | .924 | | | | |

| Sample | Sample | Subtype 1 (*n*) | Subtype 2 (*n*) | *b* | *SE* (*b*) | Cohen's *d* | *SE* (*d*) | 95% C.I. (*d*) | $p_{Tukey}$ | $\tau^2$ | $I^2$ | Pooled $p_{raw}$ | Pooled $p_{FDR}$ |
|---|---|---|---|---|---|---|---|---|---|---|---|---|---|
| ADNI-3T | Whole sample | LP (220) | A– (501) | –0.20 | 0.24 | –0.08 | 0.09 | [–0.26; 0.10] | .659 | | | | |
| ADNI-3T | Whole sample | HS (281) | A– (501) | –0.11 | 0.19 | –0.04 | 0.08 | [–0.19; 0.10] | .829 | | | | |
| ADNI-3T | Aβ-positive | LP (143) | HS (140) | –0.20 | 0.32 | –0.08 | 0.12 | [–0.32; 0.16] | .809 | | | | |
| ADNI-3T | Aβ-positive | LP (143) | A– (154) | –0.46 | 0.34 | –0.18 | 0.13 | [–0.44; 0.08] | .365 | | | | |
| ADNI-3T | Aβ-positive | HS (140) | A– (154) | –0.27 | 0.32 | –0.10 | 0.12 | [–0.34; 0.14] | .679 | | | | |
| DELCODE-3T | Whole sample | LP (265) | HS (254) | –0.02 | 0.28 | –0.01 | 0.09 | [–0.19; 0.18] | .996 | | | | |
| DELCODE-3T | Whole sample | LP (265) | A– (392) | 0.21 | 0.27 | 0.07 | 0.09 | [–0.11; 0.25] | .716 | | | | |
| DELCODE-3T | Whole sample | HS (254) | A– (392) | 0.23 | 0.24 | 0.08 | 0.08 | [–0.08; 0.24] | .597 | | | | |
| DELCODE-3T | Aβ-positive | LP (132) | HS (71) | –0.27 | 0.47 | –0.09 | 0.15 | [–0.39; 0.21] | .834 | | | | |
| DELCODE-3T | Aβ-positive | LP (132) | A– (77) | 0.57 | 0.50 | 0.19 | 0.17 | [–0.14; 0.52] | .497 | | | | |
| DELCODE-3T | Aβ-positive | HS (71) | A– (77) | 0.84 | 0.51 | 0.28 | 0.17 | [–0.06; 0.61] | .235 | | | | |
| NACC-3T | Whole sample | LP (714) | HS (613) | 0.25 | 0.15 | 0.10 | 0.06 | [–0.01; 0.21] | .194 | | | | |
| NACC-3T | Whole sample | LP (714) | A– (2009) | 0.01 | 0.13 | 0.00 | 0.05 | [–0.09; 0.10] | .996 | | | | |
| NACC-3T | Whole sample | HS (613) | A– (2009) | –0.24 | 0.12 | –0.09 | 0.05 | [–0.19; 0.00] | .115 | | | | |
| NACC-3T | Aβ-positive | LP (90) | HS (60) | 0.16 | 0.43 | 0.06 | 0.18 | [–0.28; 0.41] | .931 | | | | |
| NACC-3T | Aβ-positive | LP (90) | A– (136) | 0.20 | 0.41 | 0.08 | 0.17 | [–0.24; 0.41] | .873 | | | | |
| NACC-3T | Aβ-positive | HS (60) | A– (136) | 0.05 | 0.39 | 0.02 | 0.16 | [–0.30; 0.33] | .992 | | | | |
| ADNI-1.5T | Whole sample | LP (199) | HS (168) | –0.30 | 0.32 | –0.10 | 0.11 | [–0.31; 0.11] | .612 | | | | |
| ADNI-1.5T | Whole sample | LP (199) | A– (234) | 0.07 | 0.34 | 0.02 | 0.11 | [–0.20; 0.24] | .979 | | | | |
| ADNI-1.5T | Whole sample | HS (168) | A– (234) | 0.37 | 0.33 | 0.12 | 0.11 | [–0.09; 0.33] | .493 | | | | |
| ADNI-1.5T | Aβ-positive | LP (86) | HS (56) | 0.20 | 0.54 | 0.06 | 0.17 | [–0.28; 0.41] | .930 | | | | |
| ADNI-1.5T | Aβ-positive | LP (86) | A– (44) | 0.62 | 0.63 | 0.20 | 0.20 | [–0.20; 0.60] | .586 | | | | |
| ADNI-1.5T | Aβ-positive | HS (56) | A– (44) | 0.42 | 0.65 | 0.14 | 0.21 | [–0.28; 0.55] | .792 | | | | |
| ARWIBO-1.5T | Whole sample | LP (26) | HS (20) | 1.25 | 1.64 | 0.27 | 0.35 | [–0.43; 0.97] | .728 | | | | |
| ARWIBO-1.5T | Whole sample | LP (26) | A– (35) | 0.36 | 1.51 | 0.08 | 0.32 | [–0.57; 0.72] | .969 | | | | |
| ARWIBO-1.5T | Whole sample | HS (20) | A– (35) | –0.88 | 1.33 | –0.19 | 0.28 | [–0.76; 0.38] | .784 | | | | |
| NACC-1.5T | Whole sample | LP (141) | HS (137) | 0.44 | 0.37 | 0.15 | 0.13 | [–0.10; 0.40] | .467 | | | | |
| NACC-1.5T | Whole sample | LP (141) | A– (244) | –0.33 | 0.36 | –0.11 | 0.12 | [–0.35; 0.13] | .644 | | | | |
| NACC-1.5T | Whole sample | HS (137) | A– (244) | –0.77 | 0.33 | –0.26 | 0.11 | [–0.47; –0.04] | .051 | | | | |
| **Outcome: FAQ (inv.)** | | | | | | | | | | | | | |
| Pooled | Whole sample | HS (1265) | A– (2970) | | | –0.13 | 0.04 | [–0.20; –0.05] | | 0.00 | 11.23 | < .001*** | .002** |
| Pooled | Whole sample | LP (1292) | A– (2970) | | | –0.41 | 0.04 | [–0.50; –0.32] | | 0.00 | 14.73 | < .001*** | < .001*** |
| Pooled | Whole sample | LP (1292) | HS (1265) | | | –0.28 | 0.04 | [–0.36; –0.20] | | 0.00 | 0.00 | < .001*** | < .001*** |
| Pooled | Aβ-positive | HS (317) | A– (394) | | | –0.29 | 0.08 | [–0.45; –0.13] | | 0.00 | 0.00 | < .001*** | < .001*** |
| Pooled | Aβ-positive | LP (431) | A– (394) | | | –0.43 | 0.08 | [–0.59; –0.27] | | 0.00 | 0.00 | < .001*** | < .001*** |
| Pooled | Aβ-positive | LP (431) | HS (317) | | | –0.14 | 0.08 | [–0.29; 0.01] | | 0.00 | 0.00 | .070 | .111 |
| ADNI-3T | Whole sample | HS (266) | A– (463) | –0.27 | 0.11 | –0.19 | 0.08 | [–0.34; –0.03] | .046* | | | | |
| ADNI-3T | Whole sample | LP (203) | A– (463) | –0.81 | 0.14 | –0.56 | 0.10 | [–0.75; –0.37] | < .001*** | | | | |
| ADNI-3T | Whole sample | LP (203) | HS (266) | –0.54 | 0.14 | –0.37 | 0.10 | [–0.56; –0.18] | < .001*** | | | | |
| ADNI-3T | Aβ-positive | HS (140) | A– (153) | –0.46 | 0.19 | –0.30 | 0.12 | [–0.54; –0.06] | .041* | | | | |
| ADNI-3T | Aβ-positive | LP (142) | A– (153) | –0.83 | 0.21 | –0.54 | 0.13 | [–0.80; –0.27] | < .001*** | | | | |
| ADNI-3T | Aβ-positive | LP (142) | HS (140) | –0.37 | 0.19 | –0.24 | 0.12 | [–0.48; 0.01] | .132 | | | | |
| DELCODE-3T | Whole sample | HS (248) | A– (381) | –0.12 | 0.11 | –0.09 | 0.08 | [–0.25; 0.07] | .527 | | | | |
| DELCODE-3T | Whole sample | LP (260) | A– (381) | –0.45 | 0.12 | –0.34 | 0.09 | [–0.52; –0.16] | < .001*** | | | | |
| DELCODE-3T | Whole sample | LP (260) | HS (248) | –0.33 | 0.13 | –0.25 | 0.10 | [–0.44; –0.06] | .025* | | | | |
| DELCODE-3T | Aβ-positive | HS (68) | A– (76) | –0.49 | 0.26 | –0.33 | 0.18 | [–0.68; 0.01] | .140 | | | | |
| DELCODE-3T | Aβ-positive | LP (130) | A– (76) | –0.52 | 0.24 | –0.36 | 0.17 | [–0.69; –0.03] | .084 | | | | |
| DELCODE-3T | Aβ-positive | LP (130) | HS (68) | –0.04 | 0.23 | –0.02 | 0.16 | [–0.34; 0.29] | .987 | | | | |
| NACC-3T | Whole sample | HS (514) | A– (1733) | –0.10 | 0.08 | –0.07 | 0.05 | [–0.17; 0.03] | .391 | | | | |
| NACC-3T | Whole sample | LP (560) | A– (1733) | –0.52 | 0.08 | –0.36 | 0.06 | [–0.47; –0.25] | < .001*** | | | | |
| NACC-3T | Whole sample | LP (560) | HS (514) | –0.43 | 0.09 | –0.29 | 0.06 | [–0.42; –0.17] | < .001*** | | | | |
| NACC-3T | Aβ-positive | HS (53) | A– (121) | –0.38 | 0.29 | –0.22 | 0.17 | [–0.56; 0.12] | .406 | | | | |
| NACC-3T | Aβ-positive | LP (73) | A– (121) | –0.58 | 0.31 | –0.34 | 0.18 | [–0.70; 0.02] | .152 | | | | |
| NACC-3T | Aβ-positive | LP (73) | HS (53) | –0.20 | 0.33 | –0.12 | 0.19 | [–0.50; 0.26] | .816 | | | | |
| ADNI-1.5T | Whole sample | HS (168) | A– (234) | –0.38 | 0.18 | –0.23 | 0.11 | [–0.44; –0.01] | .091 | | | | |
| ADNI-1.5T | Whole sample | LP (198) | A– (234) | –0.77 | 0.19 | –0.46 | 0.11 | [–0.68; –0.24] | < .001*** | | | | |
| ADNI-1.5T | Whole sample | LP (198) | HS (168) | –0.39 | 0.18 | –0.23 | 0.11 | [–0.45; –0.02] | .076 | | | | |
| ADNI-1.5T | Aβ-positive | HS (56) | A– (44) | –0.50 | 0.37 | –0.29 | 0.21 | [–0.71; 0.13] | .360 | | | | |

| Sample | Sample | Subtype 1 (*n*) | Subtype 2 (*n*) | *b* | *SE* (*b*) | Cohen's *d* | *SE* (*d*) | 95% C.I. (*d*) | $p_{Tukey}$ | $\tau^2$ | $I^2$ | Pooled $p_{raw}$ | Pooled $p_{FDR}$ |
|---|---|---|---|---|---|---|---|---|---|---|---|---|---|
| ADNI-1.5T | Aβ-positive | LP (86) | A– (44) | –0.69 | 0.35 | –0.40 | 0.21 | [–0.80; 0.01] | .128 | | | | |
| ADNI-1.5T | Aβ-positive | LP (86) | HS (56) | –0.19 | 0.30 | –0.11 | 0.18 | [–0.46; 0.24] | .810 | | | | |
| NACC-1.5T | Whole sample | HS (69) | A– (159) | –0.49 | 0.27 | –0.28 | 0.15 | [–0.58; 0.03] | .174 | | | | |
| NACC-1.5T | Whole sample | LP (71) | A– (159) | –0.79 | 0.30 | –0.45 | 0.17 | [–0.79; –0.11] | .025* | | | | |
| NACC-1.5T | Whole sample | LP (71) | HS (69) | –0.30 | 0.32 | –0.17 | 0.18 | [–0.53; 0.18] | .605 | | | | |
| **Outcome: MMSE** | | | | | | | | | | | | | |
| Pooled | Whole sample | HS (1790) | A– (4130) | | | –0.27 | 0.09 | [–0.45; –0.09] | | 0.05 | 87.08 | .003** | .006** |
| Pooled | Whole sample | LP (1605) | A– (4130) | | | –0.34 | 0.06 | [–0.46; –0.22] | | 0.01 | 57.67 | < .001*** | < .001*** |
| Pooled | Whole sample | LP (1605) | HS (1790) | | | –0.06 | 0.04 | [–0.14; 0.01] | | 0.00 | 0.00 | .090 | .129 |
| Pooled | Aβ-positive | HS (514) | A– (863) | | | –0.34 | 0.14 | [–0.61; –0.07] | | 0.07 | 78.45 | .013* | .024* |
| Pooled | Aβ-positive | LP (499) | A– (863) | | | –0.43 | 0.09 | [–0.60; –0.25] | | 0.01 | 34.73 | < .001*** | < .001*** |
| Pooled | Aβ-positive | LP (499) | HS (514) | | | –0.08 | 0.08 | [–0.24; 0.08] | | 0.01 | 23.89 | .328 | .410 |
| A4/LEARN-3T | Whole sample | LP (81) | HS (346) | –0.24 | 0.15 | –0.19 | 0.13 | [–0.44; 0.05] | .270 | | | | |
| A4/LEARN-3T | Whole sample | LP (81) | A– (809) | –0.18 | 0.14 | –0.14 | 0.12 | [–0.37; 0.09] | .441 | | | | |
| A4/LEARN-3T | Whole sample | HS (346) | A– (809) | 0.06 | 0.08 | 0.05 | 0.06 | [–0.08; 0.18] | .723 | | | | |
| A4/LEARN-3T | Aβ-positive | LP (56) | HS (191) | –0.33 | 0.19 | –0.28 | 0.15 | [–0.58; 0.03] | .178 | | | | |
| A4/LEARN-3T | Aβ-positive | LP (56) | A– (458) | –0.20 | 0.17 | –0.16 | 0.14 | [–0.44; 0.12] | .499 | | | | |
| A4/LEARN-3T | Aβ-positive | HS (191) | A– (458) | 0.14 | 0.11 | 0.11 | 0.09 | [–0.06; 0.28] | .391 | | | | |
| ADNI-3T | Whole sample | LP (220) | HS (281) | –0.16 | 0.11 | –0.13 | 0.10 | [–0.32; 0.05] | .347 | | | | |
| ADNI-3T | Whole sample | LP (220) | A– (501) | –0.48 | 0.11 | –0.40 | 0.09 | [–0.58; –0.22] | < .001*** | | | | |
| ADNI-3T | Whole sample | HS (281) | A– (501) | –0.32 | 0.09 | –0.27 | 0.08 | [–0.42; –0.12] | .001** | | | | |
| ADNI-3T | Aβ-positive | LP (143) | HS (140) | –0.21 | 0.14 | –0.18 | 0.12 | [–0.43; 0.06] | .297 | | | | |
| ADNI-3T | Aβ-positive | LP (143) | A– (154) | –0.70 | 0.15 | –0.60 | 0.13 | [–0.86; –0.34] | < .001*** | | | | |
| ADNI-3T | Aβ-positive | HS (140) | A– (154) | –0.49 | 0.14 | –0.42 | 0.12 | [–0.66; –0.18] | .002** | | | | |
| DELCODE-3T | Whole sample | LP (264) | HS (254) | –0.06 | 0.11 | –0.05 | 0.10 | [–0.24; 0.13] | .839 | | | | |
| DELCODE-3T | Whole sample | LP (264) | A– (392) | –0.28 | 0.11 | –0.24 | 0.09 | [–0.42; –0.06] | .026* | | | | |
| DELCODE-3T | Whole sample | HS (254) | A– (392) | –0.22 | 0.10 | –0.18 | 0.08 | [–0.35; –0.02] | .062 | | | | |
| DELCODE-3T | Aβ-positive | LP (131) | HS (71) | 0.18 | 0.18 | 0.16 | 0.16 | [–0.14; 0.47] | .548 | | | | |
| DELCODE-3T | Aβ-positive | LP (131) | A– (77) | –0.41 | 0.19 | –0.37 | 0.17 | [–0.70; –0.04] | .072 | | | | |
| DELCODE-3T | Aβ-positive | HS (71) | A– (77) | –0.60 | 0.19 | –0.53 | 0.17 | [–0.88; –0.19] | .006** | | | | |
| NACC-3T | Whole sample | LP (680) | HS (593) | –0.05 | 0.09 | –0.03 | 0.06 | [–0.14; 0.08] | .862 | | | | |
| NACC-3T | Whole sample | LP (680) | A– (1923) | –0.45 | 0.08 | –0.30 | 0.05 | [–0.40; –0.20] | < .001*** | | | | |
| NACC-3T | Whole sample | HS (593) | A– (1923) | –0.40 | 0.07 | –0.27 | 0.05 | [–0.37; –0.18] | < .001*** | | | | |
| NACC-3T | Aβ-positive | LP (83) | HS (56) | –0.07 | 0.28 | –0.04 | 0.19 | [–0.41; 0.32] | .970 | | | | |
| NACC-3T | Aβ-positive | LP (83) | A– (130) | –0.81 | 0.26 | –0.54 | 0.18 | [–0.89; –0.19] | .006** | | | | |
| NACC-3T | Aβ-positive | HS (56) | A– (130) | –0.74 | 0.25 | –0.50 | 0.17 | [–0.83; –0.17] | .008** | | | | |
| ADNI-1.5T | Whole sample | LP (199) | HS (168) | –0.10 | 0.13 | –0.08 | 0.11 | [–0.30; 0.13] | .722 | | | | |
| ADNI-1.5T | Whole sample | LP (199) | A– (234) | –0.47 | 0.13 | –0.40 | 0.11 | [–0.62; –0.18] | .001** | | | | |
| ADNI-1.5T | Whole sample | HS (168) | A– (234) | –0.38 | 0.13 | –0.31 | 0.11 | [–0.53; –0.10] | .011* | | | | |
| ADNI-1.5T | Aβ-positive | LP (86) | HS (56) | 0.02 | 0.22 | 0.02 | 0.18 | [–0.33; 0.37] | .994 | | | | |
| ADNI-1.5T | Aβ-positive | LP (86) | A– (44) | –0.61 | 0.26 | –0.49 | 0.21 | [–0.89; –0.08] | .048* | | | | |
| ADNI-1.5T | Aβ-positive | HS (56) | A– (44) | –0.64 | 0.27 | –0.50 | 0.21 | [–0.92; –0.08] | .048* | | | | |
| ARWIBO-1.5T | Whole sample | LP (21) | HS (12) | 0.25 | 0.76 | 0.14 | 0.43 | [–0.72; 1.00] | .944 | | | | |
| ARWIBO-1.5T | Whole sample | LP (21) | A– (29) | –0.10 | 0.67 | –0.05 | 0.38 | [–0.81; 0.70] | .988 | | | | |
| ARWIBO-1.5T | Whole sample | HS (12) | A– (29) | –0.34 | 0.62 | –0.19 | 0.35 | [–0.89; 0.51] | .845 | | | | |
| NACC-1.5T | Whole sample | LP (140) | HS (136) | 0.06 | 0.20 | 0.04 | 0.13 | [–0.21; 0.29] | .953 | | | | |
| NACC-1.5T | Whole sample | LP (140) | A– (242) | –1.07 | 0.20 | –0.67 | 0.13 | [–0.92; –0.43] | < .001*** | | | | |
| NACC-1.5T | Whole sample | HS (136) | A– (242) | –1.13 | 0.18 | –0.71 | 0.11 | [–0.93; –0.49] | < .001*** | | | | |
| **Outcome: Logical Memory Delayed Recall** | | | | | | | | | | | | | |
| Pooled | Whole sample | HS (1778) | A– (4154) | | | –0.16 | 0.04 | [–0.25; –0.08] | | 0.00 | 46.59 | < .001*** | < .001*** |
| Pooled | Whole sample | LP (1585) | A– (4154) | | | –0.52 | 0.03 | [–0.59; –0.45] | | 0.00 | 0.00 | < .001*** | < .001*** |
| Pooled | Whole sample | LP (1585) | HS (1778) | | | –0.37 | 0.04 | [–0.44; –0.29] | | 0.00 | 0.00 | < .001*** | < .001*** |
| Pooled | Aβ-positive | HS (512) | A– (859) | | | –0.34 | 0.08 | [–0.49; –0.18] | | 0.01 | 34.98 | < .001*** | < .001*** |
| Pooled | Aβ-positive | LP (503) | A– (859) | | | –0.81 | 0.14 | [–1.09; –0.54] | | 0.07 | 71.92 | < .001*** | < .001*** |
| Pooled | Aβ-positive | LP (503) | HS (512) | | | –0.44 | 0.07 | [–0.57; –0.30] | | 0.00 | 0.00 | < .001*** | < .001*** |
| A4/LEARN-3T | Whole sample | LP (80) | HS (341) | –1.17 | 0.43 | –0.34 | 0.13 | [–0.59; –0.09] | .018* | | | | |
| A4/LEARN-3T | Whole sample | LP (80) | A– (798) | –1.59 | 0.41 | –0.46 | 0.12 | [–0.70; –0.23] | < .001*** | | | | |
| A4/LEARN-3T | Whole sample | HS (341) | A– (798) | –0.42 | 0.22 | –0.12 | 0.07 | [–0.25; 0.01] | .148 | | | | |

| **Sample** | **Sample** | **Subtype 1 (*n*)** | **Subtype 2 (*n*)** | ***b*** | ***SE* (*b*)** | **Cohen's *d*** | ***SE* (*d*)** | **95% C.I. (*d*)** | $p_{\mathrm{Tukey}}$ | $\tau^2$ | $I^2$ | **Pooled** $p_{\mathrm{raw}}$ | **Pooled** $p_{\mathrm{FDR}}$ |
|---|---|---|---|---|---|---|---|---|---|---|---|---|---|
| A4/LEARN-3T | Aβ-positive | LP (56) | HS (188) | −1.37 | 0.55 | −0.39 | 0.16 | [−0.69; −0.08] | .035* | | | | |
| A4/LEARN-3T | Aβ-positive | LP (56) | A− (448) | −2.03 | 0.51 | −0.57 | 0.15 | [−0.86; −0.29] | < .001*** | | | | |
| A4/LEARN-3T | Aβ-positive | HS (188) | A− (448) | −0.66 | 0.31 | −0.19 | 0.09 | [−0.36; −0.01] | .083 | | | | |
| ADNI-3T | Whole sample | LP (219) | HS (281) | −1.34 | 0.28 | −0.45 | 0.10 | [−0.63; −0.26] | < .001*** | | | | |
| ADNI-3T | Whole sample | LP (219) | A− (500) | −1.74 | 0.28 | −0.58 | 0.09 | [−0.77; −0.40] | < .001*** | | | | |
| ADNI-3T | Whole sample | HS (281) | A− (500) | −0.41 | 0.23 | −0.14 | 0.08 | [−0.29; 0.01] | .173 | | | | |
| ADNI-3T | Aβ-positive | LP (143) | HS (140) | −1.22 | 0.36 | −0.42 | 0.12 | [−0.66; −0.18] | .002** | | | | |
| ADNI-3T | Aβ-positive | LP (143) | A− (154) | −2.37 | 0.38 | −0.82 | 0.14 | [−1.08; −0.55] | < .001*** | | | | |
| ADNI-3T | Aβ-positive | HS (140) | A− (154) | −1.16 | 0.35 | −0.40 | 0.12 | [−0.64; −0.16] | .003** | | | | |
| DELCODE-3T | Whole sample | LP (264) | HS (253) | −1.57 | 0.36 | −0.41 | 0.10 | [−0.60; −0.22] | < .001*** | | | | |
| DELCODE-3T | Whole sample | LP (264) | A− (392) | −1.72 | 0.35 | −0.45 | 0.09 | [−0.63; −0.27] | < .001*** | | | | |
| DELCODE-3T | Whole sample | HS (253) | A− (392) | −0.15 | 0.31 | −0.04 | 0.08 | [−0.20; 0.12] | .888 | | | | |
| DELCODE-3T | Aβ-positive | LP (132) | HS (70) | −1.01 | 0.61 | −0.26 | 0.16 | [−0.57; 0.05] | .227 | | | | |
| DELCODE-3T | Aβ-positive | LP (132) | A− (77) | −1.81 | 0.65 | −0.47 | 0.17 | [−0.80; −0.13] | .015* | | | | |
| DELCODE-3T | Aβ-positive | HS (70) | A− (77) | −0.80 | 0.67 | −0.21 | 0.17 | [−0.55; 0.14] | .463 | | | | |
| NACC-3T | Whole sample | LP (690) | HS (602) | −1.41 | 0.23 | −0.35 | 0.06 | [−0.47; −0.24] | < .001*** | | | | |
| NACC-3T | Whole sample | LP (690) | A− (1987) | −1.95 | 0.20 | −0.49 | 0.05 | [−0.59; −0.39] | < .001*** | | | | |
| NACC-3T | Whole sample | HS (602) | A− (1987) | −0.53 | 0.19 | −0.13 | 0.05 | [−0.23; −0.04] | .015* | | | | |
| NACC-3T | Aβ-positive | LP (86) | HS (59) | −2.61 | 0.70 | −0.68 | 0.18 | [−1.04; −0.31] | < .001*** | | | | |
| NACC-3T | Aβ-positive | LP (86) | A− (136) | −4.69 | 0.65 | −1.22 | 0.18 | [−1.56; −0.87] | < .001*** | | | | |
| NACC-3T | Aβ-positive | HS (59) | A− (136) | −2.09 | 0.63 | −0.54 | 0.16 | [−0.86; −0.22] | .003** | | | | |
| ADNI-1.5T | Whole sample | LP (198) | HS (167) | −1.23 | 0.31 | −0.43 | 0.11 | [−0.64; −0.22] | < .001*** | | | | |
| ADNI-1.5T | Whole sample | LP (198) | A− (234) | −2.08 | 0.32 | −0.72 | 0.11 | [−0.95; −0.50] | < .001*** | | | | |
| ADNI-1.5T | Whole sample | HS (167) | A− (234) | −0.84 | 0.31 | −0.29 | 0.11 | [−0.51; −0.08] | .020* | | | | |
| ADNI-1.5T | Aβ-positive | LP (86) | HS (55) | −1.19 | 0.39 | −0.54 | 0.18 | [−0.89; −0.19] | .007** | | | | |
| ADNI-1.5T | Aβ-positive | LP (86) | A− (44) | −2.35 | 0.45 | −1.06 | 0.21 | [−1.49; −0.64] | < .001*** | | | | |
| ADNI-1.5T | Aβ-positive | HS (55) | A− (44) | −1.16 | 0.47 | −0.53 | 0.21 | [−0.95; −0.10] | .038* | | | | |
| NACC-1.5T | Whole sample | LP (134) | HS (134) | −0.49 | 0.49 | −0.13 | 0.13 | [−0.38; 0.13] | .577 | | | | |
| NACC-1.5T | Whole sample | LP (134) | A− (243) | −2.09 | 0.47 | −0.56 | 0.13 | [−0.80; −0.31] | < .001*** | | | | |
| NACC-1.5T | Whole sample | HS (134) | A− (243) | −1.60 | 0.42 | −0.43 | 0.11 | [−0.65; −0.20] | < .001*** | | | | |
| **Outcome: TMT B–A (inv.)** | | | | | | | | | | | | | |
| Pooled | Whole sample | HS (1144) | A− (3098) | | | −0.21 | 0.04 | [−0.28; −0.14] | | 0.00 | 0.00 | < .001*** | < .001*** |
| Pooled | Whole sample | LP (1105) | A− (3098) | | | −0.01 | 0.04 | [−0.08; 0.07] | | 0.00 | 0.00 | .871 | .899 |
| Pooled | Whole sample | LP (1105) | HS (1144) | | | 0.20 | 0.04 | [0.12; 0.29] | | 0.00 | 0.00 | < .001*** | < .001*** |
| Pooled | Aβ-positive | HS (252) | A− (390) | | | −0.28 | 0.09 | [−0.45; −0.11] | | 0.00 | 2.70 | .001** | .002** |
| Pooled | Aβ-positive | LP (339) | A− (390) | | | 0.00 | 0.09 | [−0.17; 0.17] | | 0.00 | 0.00 | .977 | .990 |
| Pooled | Aβ-positive | LP (339) | HS (252) | | | 0.30 | 0.09 | [0.12; 0.48] | | 0.00 | 2.21 | .001** | .002** |
| ADNI-3T | Whole sample | HS (249) | A− (462) | −0.34 | 0.12 | −0.24 | 0.08 | [−0.39; −0.08] | .009** | | | | |
| ADNI-3T | Whole sample | LP (180) | A− (462) | 0.00 | 0.14 | 0.00 | 0.10 | [−0.20; 0.19] | .999 | | | | |
| ADNI-3T | Whole sample | LP (180) | HS (249) | 0.34 | 0.15 | 0.23 | 0.10 | [0.03; 0.44] | .063 | | | | |
| ADNI-3T | Aβ-positive | HS (123) | A− (151) | −0.57 | 0.19 | −0.37 | 0.13 | [−0.62; −0.12] | .009** | | | | |
| ADNI-3T | Aβ-positive | LP (122) | A− (151) | 0.04 | 0.21 | 0.03 | 0.14 | [−0.24; 0.30] | .978 | | | | |
| ADNI-3T | Aβ-positive | LP (122) | HS (123) | 0.61 | 0.20 | 0.40 | 0.13 | [0.14; 0.66] | .007** | | | | |
| DELCODE-3T | Whole sample | HS (234) | A− (383) | −0.19 | 0.11 | −0.15 | 0.08 | [−0.32; 0.01] | .170 | | | | |
| DELCODE-3T | Whole sample | LP (207) | A− (383) | −0.03 | 0.12 | −0.03 | 0.10 | [−0.22; 0.16] | .959 | | | | |
| DELCODE-3T | Whole sample | LP (207) | HS (234) | 0.16 | 0.13 | 0.12 | 0.10 | [−0.08; 0.33] | .445 | | | | |
| DELCODE-3T | Aβ-positive | HS (57) | A− (72) | −0.17 | 0.24 | −0.13 | 0.18 | [−0.49; 0.23] | .747 | | | | |
| DELCODE-3T | Aβ-positive | LP (89) | A− (72) | 0.00 | 0.24 | 0.00 | 0.18 | [−0.35; 0.35] | .999 | | | | |
| DELCODE-3T | Aβ-positive | LP (89) | HS (57) | 0.17 | 0.24 | 0.13 | 0.18 | [−0.22; 0.49] | .745 | | | | |
| NACC-3T | Whole sample | HS (455) | A− (1826) | −0.29 | 0.08 | −0.20 | 0.05 | [−0.31; −0.10] | < .001*** | | | | |
| NACC-3T | Whole sample | LP (473) | A− (1826) | 0.05 | 0.08 | 0.03 | 0.06 | [−0.08; 0.14] | .839 | | | | |
| NACC-3T | Whole sample | LP (473) | HS (455) | 0.34 | 0.10 | 0.23 | 0.07 | [0.10; 0.37] | .002** | | | | |
| NACC-3T | Aβ-positive | HS (33) | A− (123) | −0.10 | 0.28 | −0.07 | 0.20 | [−0.47; 0.33] | .935 | | | | |
| NACC-3T | Aβ-positive | LP (51) | A− (123) | −0.06 | 0.29 | −0.04 | 0.21 | [−0.45; 0.37] | .980 | | | | |
| NACC-3T | Aβ-positive | LP (51) | HS (33) | 0.04 | 0.36 | 0.03 | 0.26 | [−0.48; 0.54] | .992 | | | | |
| ADNI-1.5T | Whole sample | HS (123) | A− (233) | −0.37 | 0.15 | −0.28 | 0.12 | [−0.51; −0.05] | .042* | | | | |
| ADNI-1.5T | Whole sample | LP (171) | A− (233) | −0.13 | 0.15 | −0.09 | 0.12 | [−0.32; 0.13] | .694 | | | | |
| ADNI-1.5T | Whole sample | LP (171) | HS (123) | 0.24 | 0.16 | 0.19 | 0.12 | [−0.06; 0.43] | .296 | | | | |

| Sample | Sample | Subtype 1 ($n$) | Subtype 2 ($n$) | $b$ | $SE$ ($b$) | Cohen's $d$ | $SE$ ($d$) | 95% C.I. ($d$) | $p_{Tukey}$ | $\tau^2$ | $I^2$ | Pooled $p_{raw}$ | Pooled $p_{FDR}$ |
|---|---|---|---|---|---|---|---|---|---|---|---|---|---|
| ADNI-1.5T | Aβ-positive | HS (39) | A– (44) | −0.69 | 0.32 | −0.50 | 0.23 | [−0.95; −0.04] | .080 | | | | |
| ADNI-1.5T | Aβ-positive | LP (77) | A– (44) | −0.05 | 0.29 | −0.04 | 0.21 | [−0.45; 0.38] | .983 | | | | |
| ADNI-1.5T | Aβ-positive | LP (77) | HS (39) | 0.64 | 0.30 | 0.46 | 0.21 | [0.04; 0.88] | .081 | | | | |
| NACC-1.5T | Whole sample | HS (83) | A– (194) | −0.43 | 0.20 | −0.29 | 0.14 | [−0.56; −0.02] | .083 | | | | |
| NACC-1.5T | Whole sample | LP (74) | A– (194) | −0.15 | 0.24 | −0.10 | 0.16 | [−0.41; 0.21] | .806 | | | | |
| NACC-1.5T | Whole sample | LP (74) | HS (83) | 0.29 | 0.26 | 0.19 | 0.17 | [−0.15; 0.53] | .509 | | | | |

*$p$ < .05. **$p$ < .01. ***$p$ < .001. Abbreviations: A–, atrophy negative. HS, hippocampal-sparing. inv., inverted. LP, limbic-predominant.

**Supplementary Table 3 Summary statistics of cross-sectional stage effects derived from partial correlation predicting numeric outcome variables.**

| Sample | Sample | Subtype (*n*) | PCC | Fisher's *z*-transformed PCC | SE | 95% C.I. | *p* | $\tau^2$ | $I^2$ | Pooled $p_{raw}$ | Pooled $p_{FDR}$ |
|---|---|---|---|---|---|---|---|---|---|---|---|
| **Outcome: age (years)** | | | | | | | | | | | |
| Pooled | Whole sample | HS (1963) | | 0.09 | 0.05 | [−0.01; 0.19] | | 0.01 | 76.33 | < .001*** | .100 |
| Pooled | Whole sample | LP (1767) | | −0.05 | 0.05 | [−0.15; 0.05] | | 0.01 | 73.41 | < .001*** | .457 |
| Pooled | Aβ-positive | HS (525) | | 0.08 | 0.08 | [−0.08; 0.23] | | 0.02 | 65.11 | < .001*** | .426 |
| Pooled | Aβ-positive | LP (518) | | 0.02 | 0.10 | [−0.16; 0.21] | | 0.04 | 77.46 | < .001*** | .946 |
| A4/LEARN-3T | Whole sample | HS (347) | 0.18 | 0.18 | 0.05 | [0.08; 0.28] | < .001*** | | | | |
| A4/LEARN-3T | Whole sample | LP (81) | 0.29 | 0.29 | 0.11 | [0.07; 0.48] | .010** | | | | |
| A4/LEARN-3T | Aβ-positive | HS (191) | 0.26 | 0.27 | 0.07 | [0.12; 0.39] | < .001*** | | | | |
| A4/LEARN-3T | Aβ-positive | LP (56) | 0.28 | 0.29 | 0.14 | [0.02; 0.51] | .039* | | | | |
| ADNI-3T | Whole sample | HS (289) | 0.07 | 0.07 | 0.06 | [−0.05; 0.18] | .249 | | | | |
| ADNI-3T | Whole sample | LP (225) | −0.07 | −0.07 | 0.07 | [−0.20; 0.07] | .329 | | | | |
| ADNI-3T | Aβ-positive | HS (143) | 0.07 | 0.07 | 0.09 | [−0.09; 0.23] | .395 | | | | |
| ADNI-3T | Aβ-positive | LP (145) | −0.01 | −0.01 | 0.08 | [−0.18; 0.15] | .867 | | | | |
| DELCODE-3T | Whole sample | HS (254) | 0.13 | 0.13 | 0.06 | [0.00; 0.24] | .047* | | | | |
| DELCODE-3T | Whole sample | LP (265) | 0.00 | 0.00 | 0.06 | [−0.12; 0.12] | .972 | | | | |
| DELCODE-3T | Aβ-positive | HS (71) | 0.19 | 0.19 | 0.12 | [−0.05; 0.41] | .124 | | | | |
| DELCODE-3T | Aβ-positive | LP (132) | 0.01 | 0.01 | 0.09 | [−0.16; 0.18] | .902 | | | | |
| NACC-3T | Whole sample | HS (698) | −0.07 | −0.07 | 0.04 | [−0.14; 0.00] | .062 | | | | |
| NACC-3T | Whole sample | LP (784) | −0.18 | −0.18 | 0.04 | [−0.25; −0.11] | < .001*** | | | | |
| NACC-3T | Aβ-positive | HS (64) | −0.15 | −0.15 | 0.13 | [−0.38; 0.10] | .247 | | | | |
| NACC-3T | Aβ-positive | LP (98) | −0.29 | −0.30 | 0.10 | [−0.47; −0.10] | .004** | | | | |
| ADNI-1.5T | Whole sample | HS (170) | −0.03 | −0.03 | 0.08 | [−0.18; 0.12] | .672 | | | | |
| ADNI-1.5T | Whole sample | LP (200) | −0.04 | −0.04 | 0.07 | [−0.18; 0.10] | .573 | | | | |
| ADNI-1.5T | Aβ-positive | HS (56) | −0.10 | −0.10 | 0.14 | [−0.36; 0.17] | .469 | | | | |
| ADNI-1.5T | Aβ-positive | LP (87) | 0.19 | 0.19 | 0.11 | [−0.02; 0.39] | .083 | | | | |
| ARWIBO-1.5T | Whole sample | HS (38) | 0.09 | 0.09 | 0.17 | [−0.25; 0.40] | .611 | | | | |
| ARWIBO-1.5T | Whole sample | LP (54) | −0.25 | −0.25 | 0.14 | [−0.49; 0.03] | .079 | | | | |
| NACC-1.5T | Whole sample | HS (167) | 0.28 | 0.29 | 0.08 | [0.14; 0.42] | < .001*** | | | | |
| NACC-1.5T | Whole sample | LP (158) | −0.07 | −0.07 | 0.08 | [−0.22; 0.09] | .406 | | | | |
| **Outcome: education (years)** | | | | | | | | | | | |
| Pooled | Whole sample | HS (1820) | | −0.06 | 0.02 | [−0.11; −0.02] | | 0.00 | 0.00 | < .001*** | .015* |
| Pooled | Whole sample | LP (1646) | | −0.01 | 0.03 | [−0.07; 0.05] | | 0.00 | 19.17 | < .001*** | .867 |
| Pooled | Aβ-positive | HS (518) | | 0.01 | 0.04 | [−0.08; 0.09] | | 0.00 | 0.00 | < .001*** | .910 |
| Pooled | Aβ-positive | LP (507) | | 0.00 | 0.06 | [−0.13; 0.12] | | 0.01 | 47.36 | < .001*** | .986 |
| A4/LEARN-3T | Whole sample | HS (347) | 0.01 | 0.01 | 0.05 | [−0.10; 0.11] | .863 | | | | |
| A4/LEARN-3T | Whole sample | LP (81) | 0.20 | 0.21 | 0.11 | [−0.02; 0.40] | .071 | | | | |
| A4/LEARN-3T | Aβ-positive | HS (191) | 0.02 | 0.02 | 0.07 | [−0.12; 0.16] | .788 | | | | |
| A4/LEARN-3T | Aβ-positive | LP (56) | 0.28 | 0.29 | 0.14 | [0.02; 0.51] | .035* | | | | |
| ADNI-3T | Whole sample | HS (281) | −0.09 | −0.09 | 0.06 | [−0.20; 0.03] | .143 | | | | |
| ADNI-3T | Whole sample | LP (220) | −0.12 | −0.12 | 0.07 | [−0.24; 0.02] | .088 | | | | |
| ADNI-3T | Aβ-positive | HS (140) | −0.02 | −0.02 | 0.09 | [−0.19; 0.14] | .792 | | | | |
| ADNI-3T | Aβ-positive | LP (143) | −0.14 | −0.14 | 0.09 | [−0.29; 0.03] | .109 | | | | |
| DELCODE-3T | Whole sample | HS (254) | −0.07 | −0.07 | 0.06 | [−0.19; 0.05] | .271 | | | | |
| DELCODE-3T | Whole sample | LP (265) | −0.07 | −0.07 | 0.06 | [−0.19; 0.05] | .275 | | | | |
| DELCODE-3T | Aβ-positive | HS (71) | −0.01 | −0.01 | 0.12 | [−0.25; 0.23] | .930 | | | | |
| DELCODE-3T | Aβ-positive | LP (132) | −0.03 | −0.03 | 0.09 | [−0.21; 0.14] | .700 | | | | |
| NACC-3T | Whole sample | HS (613) | −0.08 | −0.09 | 0.04 | [−0.16; −0.01] | .036* | | | | |
| NACC-3T | Whole sample | LP (714) | −0.02 | −0.02 | 0.04 | [−0.09; 0.06] | .661 | | | | |
| NACC-3T | Aβ-positive | HS (60) | 0.05 | 0.05 | 0.13 | [−0.21; 0.30] | .710 | | | | |
| NACC-3T | Aβ-positive | LP (90) | −0.07 | −0.07 | 0.11 | [−0.28; 0.14] | .497 | | | | |
| ADNI-1.5T | Whole sample | HS (168) | 0.01 | 0.01 | 0.08 | [−0.14; 0.16] | .910 | | | | |
| ADNI-1.5T | Whole sample | LP (199) | 0.06 | 0.06 | 0.07 | [−0.08; 0.19] | .428 | | | | |
| ADNI-1.5T | Aβ-positive | HS (56) | 0.00 | 0.00 | 0.14 | [−0.27; 0.27] | .984 | | | | |
| ADNI-1.5T | Aβ-positive | LP (86) | 0.07 | 0.07 | 0.11 | [−0.15; 0.28] | .544 | | | | |
| ARWIBO-1.5T | Whole sample | HS (20) | 0.22 | 0.22 | 0.25 | [−0.26; 0.61] | .374 | | | | |
| ARWIBO-1.5T | Whole sample | LP (26) | 0.01 | 0.01 | 0.22 | [−0.39; 0.41] | .951 | | | | |
| NACC-1.5T | Whole sample | HS (137) | −0.20 | −0.20 | 0.09 | [−0.35; −0.03] | .023* | | | | |

| Sample | Sample | Subtype (*n*) | PCC | Fisher's *z*-transformed PCC | SE | 95% C.I. | *p* | $\tau^2$ | $I^2$ | Pooled $p_{raw}$ | Pooled $p_{FDR}$ |
|---|---|---|---|---|---|---|---|---|---|---|---|
| NACC-1.5T | Whole sample | LP (141) | 0.06 | 0.06 | 0.09 | [−0.11; 0.22] | .489 | | | | |
| **Outcome: FAQ (inv.)** | | | | | | | | | | | |
| Pooled | Whole sample | HS (1265) | | −0.09 | 0.05 | [−0.18; 0.00] | | 0.01 | 53.16 | < .001*** | .059 |
| Pooled | Whole sample | LP (1292) | | −0.19 | 0.03 | [−0.25; −0.14] | | 0.00 | 0.00 | < .001*** | < .001*** |
| Pooled | Aβ-positive | HS (317) | | −0.23 | 0.06 | [−0.35; −0.11] | | 0.00 | 0.00 | < .001*** | < .001*** |
| Pooled | Aβ-positive | LP (431) | | −0.10 | 0.06 | [−0.22; 0.03] | | 0.01 | 36.47 | < .001*** | .247 |
| ADNI-3T | Whole sample | HS (266) | −0.23 | −0.23 | 0.06 | [−0.34; −0.11] | < .001*** | | | | |
| ADNI-3T | Whole sample | LP (203) | −0.22 | −0.23 | 0.07 | [−0.35; −0.09] | .002** | | | | |
| ADNI-3T | Aβ-positive | HS (140) | −0.21 | −0.21 | 0.09 | [−0.36; −0.04] | .017* | | | | |
| ADNI-3T | Aβ-positive | LP (142) | −0.20 | −0.20 | 0.09 | [−0.36; −0.03] | .020* | | | | |
| DELCODE-3T | Whole sample | HS (248) | −0.10 | −0.10 | 0.06 | [−0.22; 0.03] | .125 | | | | |
| DELCODE-3T | Whole sample | LP (260) | −0.18 | −0.18 | 0.06 | [−0.29; −0.05] | .005** | | | | |
| DELCODE-3T | Aβ-positive | HS (68) | −0.18 | −0.19 | 0.13 | [−0.41; 0.07] | .153 | | | | |
| DELCODE-3T | Aβ-positive | LP (130) | −0.12 | −0.12 | 0.09 | [−0.29; 0.06] | .200 | | | | |
| NACC-3T | Whole sample | HS (514) | −0.08 | −0.08 | 0.04 | [−0.16; 0.01] | .075 | | | | |
| NACC-3T | Whole sample | LP (560) | −0.21 | −0.21 | 0.04 | [−0.28; −0.12] | < .001*** | | | | |
| NACC-3T | Aβ-positive | HS (53) | −0.29 | −0.30 | 0.15 | [−0.53; 0.00] | .048* | | | | |
| NACC-3T | Aβ-positive | LP (73) | 0.13 | 0.13 | 0.12 | [−0.11; 0.36] | .283 | | | | |
| ADNI-1.5T | Whole sample | HS (168) | −0.05 | −0.05 | 0.08 | [−0.20; 0.11] | .553 | | | | |
| ADNI-1.5T | Whole sample | LP (198) | −0.11 | −0.11 | 0.07 | [−0.25; 0.03] | .114 | | | | |
| ADNI-1.5T | Aβ-positive | HS (56) | −0.27 | −0.27 | 0.15 | [−0.51; 0.01] | .062 | | | | |
| ADNI-1.5T | Aβ-positive | LP (86) | −0.13 | −0.13 | 0.11 | [−0.34; 0.09] | .241 | | | | |
| NACC-1.5T | Whole sample | HS (69) | 0.15 | 0.15 | 0.13 | [−0.10; 0.38] | .250 | | | | |
| NACC-1.5T | Whole sample | LP (71) | −0.22 | −0.22 | 0.13 | [−0.44; 0.03] | .080 | | | | |
| **Outcome: MMSE** | | | | | | | | | | | |
| Pooled | Whole sample | HS (1790) | | −0.10 | 0.04 | [−0.18; −0.01] | | 0.01 | 61.20 | < .001*** | .034* |
| Pooled | Whole sample | LP (1605) | | −0.20 | 0.05 | [−0.30; −0.10] | | 0.01 | 67.26 | < .001*** | < .001*** |
| Pooled | Aβ-positive | HS (514) | | −0.10 | 0.08 | [−0.26; 0.05] | | 0.02 | 61.10 | < .001*** | .271 |
| Pooled | Aβ-positive | LP (499) | | −0.19 | 0.05 | [−0.28; −0.10] | | 0.00 | 0.00 | < .001*** | < .001*** |
| A4/LEARN-3T | Whole sample | HS (346) | 0.03 | 0.03 | 0.05 | [−0.08; 0.13] | .642 | | | | |
| A4/LEARN-3T | Whole sample | LP (81) | −0.02 | −0.02 | 0.12 | [−0.24; 0.21] | .879 | | | | |
| A4/LEARN-3T | Aβ-positive | HS (191) | 0.06 | 0.06 | 0.07 | [−0.08; 0.21] | .389 | | | | |
| A4/LEARN-3T | Aβ-positive | LP (56) | −0.07 | −0.07 | 0.14 | [−0.34; 0.21] | .614 | | | | |
| ADNI-3T | Whole sample | HS (281) | −0.24 | −0.24 | 0.06 | [−0.35; −0.12] | < .001*** | | | | |
| ADNI-3T | Whole sample | LP (220) | −0.14 | −0.14 | 0.07 | [−0.27; −0.01] | .036* | | | | |
| ADNI-3T | Aβ-positive | HS (140) | −0.32 | −0.33 | 0.09 | [−0.46; −0.16] | < .001*** | | | | |
| ADNI-3T | Aβ-positive | LP (143) | −0.21 | −0.22 | 0.09 | [−0.37; −0.05] | .012* | | | | |
| DELCODE-3T | Whole sample | HS (254) | −0.05 | −0.05 | 0.06 | [−0.17; 0.08] | .439 | | | | |
| DELCODE-3T | Whole sample | LP (264) | −0.15 | −0.15 | 0.06 | [−0.27; −0.03] | .014* | | | | |
| DELCODE-3T | Aβ-positive | HS (71) | −0.06 | −0.06 | 0.13 | [−0.30; 0.18] | .608 | | | | |
| DELCODE-3T | Aβ-positive | LP (131) | −0.13 | −0.13 | 0.09 | [−0.30; 0.04] | .136 | | | | |
| NACC-3T | Whole sample | HS (593) | −0.15 | −0.15 | 0.04 | [−0.23; −0.07] | < .001*** | | | | |
| NACC-3T | Whole sample | LP (680) | −0.29 | −0.30 | 0.04 | [−0.36; −0.22] | < .001*** | | | | |
| NACC-3T | Aβ-positive | HS (56) | −0.10 | −0.10 | 0.15 | [−0.37; 0.18] | .474 | | | | |
| NACC-3T | Aβ-positive | LP (83) | −0.22 | −0.22 | 0.12 | [−0.42; 0.01] | .056 | | | | |
| ADNI-1.5T | Whole sample | HS (168) | −0.01 | −0.01 | 0.08 | [−0.16; 0.15] | .930 | | | | |
| ADNI-1.5T | Whole sample | LP (199) | −0.08 | −0.08 | 0.07 | [−0.22; 0.06] | .253 | | | | |
| ADNI-1.5T | Aβ-positive | HS (56) | −0.08 | −0.08 | 0.15 | [−0.35; 0.20] | .573 | | | | |
| ADNI-1.5T | Aβ-positive | LP (86) | −0.25 | −0.26 | 0.11 | [−0.45; −0.03] | .024* | | | | |
| ARWIBO-1.5T | Whole sample | HS (12) | −0.19 | −0.19 | 0.45 | [−0.79; 0.60] | .658 | | | | |
| ARWIBO-1.5T | Whole sample | LP (21) | −0.54 | −0.61 | 0.28 | [−0.82; −0.06] | .030* | | | | |
| NACC-1.5T | Whole sample | HS (136) | −0.15 | −0.15 | 0.09 | [−0.32; 0.02] | .083 | | | | |
| NACC-1.5T | Whole sample | LP (140) | −0.36 | −0.38 | 0.09 | [−0.50; −0.21] | < .001*** | | | | |
| **Outcome: Logical Memory Delayed Recall** | | | | | | | | | | | |
| Pooled | Whole sample | HS (1778) | | −0.13 | 0.02 | [−0.18; −0.09] | | 0.00 | 0.90 | < .001*** | < .001*** |
| Pooled | Whole sample | LP (1585) | | −0.10 | 0.03 | [−0.15; −0.05] | | 0.00 | 0.00 | < .001*** | < .001*** |
| Pooled | Aβ-positive | HS (512) | | −0.18 | 0.05 | [−0.27; −0.08] | | 0.00 | 0.02 | < .001*** | < .001*** |
| Pooled | Aβ-positive | LP (503) | | −0.04 | 0.05 | [−0.13; 0.06] | | 0.00 | 3.33 | < .001*** | .767 |

| Sample | Sample | Subtype (*n*) | PCC | Fisher's *z*-transformed PCC | SE | 95% C.I. | *p* | $\tau^2$ | $I^2$ | Pooled $p_{raw}$ | Pooled $p_{FDR}$ |
|---|---|---|---|---|---|---|---|---|---|---|---|
| A4/LEARN-3T | Whole sample | HS (341) | –0.06 | –0.06 | 0.05 | [–0.16; 0.05] | .306 | | | | |
| A4/LEARN-3T | Whole sample | LP (80) | –0.09 | –0.09 | 0.12 | [–0.31; 0.14] | .464 | | | | |
| A4/LEARN-3T | Aβ-positive | HS (188) | –0.12 | –0.12 | 0.07 | [–0.26; 0.02] | .094 | | | | |
| A4/LEARN-3T | Aβ-positive | LP (56) | –0.06 | –0.06 | 0.14 | [–0.33; 0.22] | .675 | | | | |
| ADNI-3T | Whole sample | HS (281) | –0.10 | –0.10 | 0.06 | [–0.22; 0.02] | .098 | | | | |
| ADNI-3T | Whole sample | LP (219) | –0.13 | –0.13 | 0.07 | [–0.26; 0.00] | .051 | | | | |
| ADNI-3T | Aβ-positive | HS (140) | –0.21 | –0.22 | 0.09 | [–0.37; –0.05] | .013* | | | | |
| ADNI-3T | Aβ-positive | LP (143) | –0.13 | –0.13 | 0.09 | [–0.29; 0.03] | .120 | | | | |
| DELCODE-3T | Whole sample | HS (253) | –0.19 | –0.19 | 0.06 | [–0.31; –0.07] | .002** | | | | |
| DELCODE-3T | Whole sample | LP (264) | –0.08 | –0.08 | 0.06 | [–0.20; 0.04] | .199 | | | | |
| DELCODE-3T | Aβ-positive | HS (70) | –0.27 | –0.27 | 0.13 | [–0.48; –0.02] | .033* | | | | |
| DELCODE-3T | Aβ-positive | LP (132) | 0.04 | 0.04 | 0.09 | [–0.14; 0.21] | .672 | | | | |
| NACC-3T | Whole sample | HS (602) | –0.17 | –0.17 | 0.04 | [–0.24; –0.09] | < .001*** | | | | |
| NACC-3T | Whole sample | LP (690) | –0.13 | –0.13 | 0.04 | [–0.20; –0.06] | < .001*** | | | | |
| NACC-3T | Aβ-positive | HS (59) | 0.01 | 0.01 | 0.14 | [–0.26; 0.28] | .957 | | | | |
| NACC-3T | Aβ-positive | LP (86) | –0.09 | –0.09 | 0.11 | [–0.31; 0.13] | .417 | | | | |
| ADNI-1.5T | Whole sample | HS (167) | –0.14 | –0.14 | 0.08 | [–0.29; 0.02] | .084 | | | | |
| ADNI-1.5T | Whole sample | LP (198) | –0.03 | –0.03 | 0.07 | [–0.17; 0.11] | .678 | | | | |
| ADNI-1.5T | Aβ-positive | HS (55) | –0.31 | –0.32 | 0.15 | [–0.54; –0.03] | .031* | | | | |
| ADNI-1.5T | Aβ-positive | LP (86) | 0.09 | 0.09 | 0.11 | [–0.13; 0.31] | .420 | | | | |
| NACC-1.5T | Whole sample | HS (134) | –0.14 | –0.14 | 0.09 | [–0.30; 0.04] | .127 | | | | |
| NACC-1.5T | Whole sample | LP (134) | –0.06 | –0.06 | 0.09 | [–0.23; 0.11] | .503 | | | | |
| **Outcome: TMT B–A (inv.)** | | | | | | | | | | | |
| Pooled | Whole sample | HS (1144) | | –0.04 | 0.05 | [–0.15; 0.07] | | 0.01 | 64.48 | < .001*** | .510 |
| Pooled | Whole sample | LP (1105) | | –0.09 | 0.03 | [–0.15; –0.03] | | 0.00 | 0.02 | < .001*** | .008** |
| Pooled | Aβ-positive | HS (252) | | –0.12 | 0.09 | [–0.30; 0.06] | | 0.01 | 35.40 | < .001*** | .271 |
| Pooled | Aβ-positive | LP (339) | | –0.11 | 0.07 | [–0.25; 0.03] | | 0.01 | 30.09 | < .001*** | .247 |
| ADNI-3T | Whole sample | HS (249) | –0.20 | –0.20 | 0.06 | [–0.32; –0.07] | .002** | | | | |
| ADNI-3T | Whole sample | LP (180) | –0.20 | –0.20 | 0.08 | [–0.34; –0.05] | .008** | | | | |
| ADNI-3T | Aβ-positive | HS (123) | –0.22 | –0.22 | 0.09 | [–0.39; –0.04] | .018* | | | | |
| ADNI-3T | Aβ-positive | LP (122) | –0.22 | –0.22 | 0.09 | [–0.39; –0.04] | .018* | | | | |
| DELCODE-3T | Whole sample | HS (234) | –0.07 | –0.07 | 0.07 | [–0.20; 0.06] | .315 | | | | |
| DELCODE-3T | Whole sample | LP (207) | –0.02 | –0.02 | 0.07 | [–0.15; 0.12] | .814 | | | | |
| DELCODE-3T | Aβ-positive | HS (57) | 0.12 | 0.12 | 0.14 | [–0.17; 0.38] | .419 | | | | |
| DELCODE-3T | Aβ-positive | LP (89) | 0.06 | 0.06 | 0.11 | [–0.16; 0.27] | .619 | | | | |
| NACC-3T | Whole sample | HS (455) | 0.00 | 0.00 | 0.05 | [–0.10; 0.09] | .932 | | | | |
| NACC-3T | Whole sample | LP (473) | –0.09 | –0.09 | 0.05 | [–0.18; 0.00] | .061 | | | | |
| NACC-3T | Aβ-positive | HS (33) | –0.27 | –0.28 | 0.20 | [–0.59; 0.12] | .172 | | | | |
| NACC-3T | Aβ-positive | LP (51) | –0.20 | –0.20 | 0.15 | [–0.46; 0.10] | .195 | | | | |
| ADNI-1.5T | Whole sample | HS (123) | –0.02 | –0.02 | 0.09 | [–0.21; 0.16] | .791 | | | | |
| ADNI-1.5T | Whole sample | LP (171) | –0.12 | –0.12 | 0.08 | [–0.27; 0.04] | .131 | | | | |
| ADNI-1.5T | Aβ-positive | HS (39) | –0.10 | –0.10 | 0.18 | [–0.43; 0.25] | .585 | | | | |
| ADNI-1.5T | Aβ-positive | LP (77) | –0.08 | –0.08 | 0.12 | [–0.31; 0.15] | .496 | | | | |
| NACC-1.5T | Whole sample | HS (83) | 0.19 | 0.19 | 0.12 | [–0.04; 0.40] | .101 | | | | |
| NACC-1.5T | Whole sample | LP (74) | 0.07 | 0.07 | 0.12 | [–0.17; 0.30] | .586 | | | | |

*$p < .05$. **$p < .01$. ***$p < .001$. Abbreviations: HS, hippocampal-sparing. inv., inverted. LP, limbic-predominant.

**Supplementary Table 4 Summary statistics of cross-sectional subtype effects derived from logistic regression models predicting binary outcome variables.**

| Sample | Sample | Subtype 1 (*n*) | Subtype 2 (*n*) | Used Firth's penalization | log(*OR*) | *SE* | *OR* | *SE* | 95% C.I. | *p* | $\tau^2$ | $I^2$ | Pooled $p_{raw}$ | Pooled $p_{FDR}$ |
|---|---|---|---|---|---|---|---|---|---|---|---|---|---|---|
| **Outcome: female *versus* male sex** | | | | | | | | | | | | | | |
| Pooled | Whole sample | HS (1963) | A– (4685) | no | 0.03 | 0.11 | 1.03 | 1.11 | [0.84; 1.28] | | 0.05 | 65.50 | .766 | .842 |
| Pooled | Whole sample | LP (1767) | A– (4685) | no | 0.05 | 0.15 | 1.05 | 1.16 | [0.79; 1.40] | | 0.10 | 72.36 | .715 | .842 |
| Pooled | Whole sample | LP (1767) | HS (1963) | no | 0.05 | 0.19 | 1.05 | 1.21 | [0.72; 1.53] | | 0.20 | 82.95 | .814 | .866 |
| Pooled | Aβ-positive | HS (525) | A– (893) | no | 0.27 | 0.12 | 1.31 | 1.12 | [1.04; 1.65] | | 0.00 | 0.00 | .020* | .033* |
| Pooled | Aβ-positive | LP (518) | A– (893) | no | 0.00 | 0.24 | 1.00 | 1.28 | [0.62; 1.62] | | 0.18 | 62.79 | .990 | .990 |
| Pooled | Aβ-positive | LP (518) | HS (525) | no | −0.23 | 0.29 | 0.80 | 1.33 | [0.45; 1.40] | | 0.31 | 75.32 | .431 | .517 |
| A4/LEARN-3T | Whole sample | HS (347) | A– (809) | no | 0.40 | 0.13 | 1.49 | 1.14 | [1.10; 2.02] | .006** | | | | |
| A4/LEARN-3T | Whole sample | LP (81) | A– (809) | no | −0.57 | 0.27 | 0.56 | 1.31 | [0.30; 1.06] | .082 | | | | |
| A4/LEARN-3T | Whole sample | LP (81) | HS (347) | no | −0.97 | 0.28 | 0.38 | 1.32 | [0.20; 0.73] | .001** | | | | |
| A4/LEARN-3T | Aβ-positive | HS (191) | A– (458) | no | 0.38 | 0.17 | 1.46 | 1.19 | [0.97; 2.20] | .073 | | | | |
| A4/LEARN-3T | Aβ-positive | LP (56) | A– (458) | no | −0.77 | 0.34 | 0.46 | 1.40 | [0.21; 1.02] | .056 | | | | |
| A4/LEARN-3T | Aβ-positive | LP (56) | HS (191) | no | −1.16 | 0.35 | 0.31 | 1.42 | [0.14; 0.72] | .003** | | | | |
| ADNI-3T | Whole sample | HS (289) | A– (538) | no | −0.13 | 0.15 | 0.88 | 1.16 | [0.62; 1.25] | .670 | | | | |
| ADNI-3T | Whole sample | LP (225) | A– (538) | no | 0.31 | 0.18 | 1.36 | 1.20 | [0.89; 2.09] | .209 | | | | |
| ADNI-3T | Whole sample | LP (225) | HS (289) | no | 0.44 | 0.19 | 1.55 | 1.21 | [0.99; 2.42] | .054 | | | | |
| ADNI-3T | Aβ-positive | HS (143) | A– (162) | no | 0.09 | 0.24 | 1.09 | 1.28 | [0.62; 1.94] | .929 | | | | |
| ADNI-3T | Aβ-positive | LP (145) | A– (162) | no | 0.55 | 0.26 | 1.74 | 1.30 | [0.94; 3.23] | .090 | | | | |
| ADNI-3T | Aβ-positive | LP (145) | HS (143) | no | 0.47 | 0.25 | 1.59 | 1.28 | [0.89; 2.83] | .141 | | | | |
| DELCODE-3T | Whole sample | HS (254) | A– (392) | no | −0.17 | 0.16 | 0.84 | 1.18 | [0.57; 1.23] | .533 | | | | |
| DELCODE-3T | Whole sample | LP (265) | A– (392) | no | −0.16 | 0.18 | 0.85 | 1.20 | [0.55; 1.31] | .663 | | | | |
| DELCODE-3T | Whole sample | LP (265) | HS (254) | no | 0.02 | 0.19 | 1.02 | 1.21 | [0.65; 1.58] | .996 | | | | |
| DELCODE-3T | Aβ-positive | HS (71) | A– (77) | no | −0.02 | 0.35 | 0.98 | 1.42 | [0.43; 2.22] | .998 | | | | |
| DELCODE-3T | Aβ-positive | LP (132) | A– (77) | no | 0.07 | 0.34 | 1.08 | 1.41 | [0.48; 2.39] | .975 | | | | |
| DELCODE-3T | Aβ-positive | LP (132) | HS (71) | no | 0.09 | 0.31 | 1.10 | 1.37 | [0.53; 2.28] | .953 | | | | |
| NACC-3T | Whole sample | HS (698) | A– (2319) | no | 0.20 | 0.09 | 1.23 | 1.10 | [0.99; 1.53] | .074 | | | | |
| NACC-3T | Whole sample | LP (784) | A– (2319) | no | 0.03 | 0.10 | 1.03 | 1.10 | [0.82; 1.29] | .963 | | | | |
| NACC-3T | Whole sample | LP (784) | HS (698) | no | −0.18 | 0.11 | 0.84 | 1.12 | [0.64; 1.09] | .244 | | | | |
| NACC-3T | Aβ-positive | HS (64) | A– (152) | no | 0.37 | 0.32 | 1.45 | 1.37 | [0.69; 3.04] | .470 | | | | |
| NACC-3T | Aβ-positive | LP (98) | A– (152) | no | −0.23 | 0.31 | 0.79 | 1.37 | [0.38; 1.65] | .738 | | | | |
| NACC-3T | Aβ-positive | LP (98) | HS (64) | no | −0.60 | 0.34 | 0.55 | 1.41 | [0.24; 1.22] | .185 | | | | |
| ADNI-1.5T | Whole sample | HS (170) | A– (236) | no | 0.00 | 0.23 | 1.00 | 1.26 | [0.59; 1.71] | .999 | | | | |
| ADNI-1.5T | Whole sample | LP (200) | A– (236) | no | 0.62 | 0.23 | 1.86 | 1.26 | [1.08; 3.20] | .020* | | | | |
| ADNI-1.5T | Whole sample | LP (200) | HS (170) | no | 0.62 | 0.22 | 1.86 | 1.25 | [1.11; 3.11] | .014* | | | | |
| ADNI-1.5T | Aβ-positive | HS (56) | A– (44) | no | 0.46 | 0.46 | 1.59 | 1.58 | [0.54; 4.63] | .568 | | | | |
| ADNI-1.5T | Aβ-positive | LP (87) | A– (44) | no | 0.40 | 0.44 | 1.49 | 1.56 | [0.53; 4.21] | .639 | | | | |
| ADNI-1.5T | Aβ-positive | LP (87) | HS (56) | no | −0.06 | 0.36 | 0.94 | 1.43 | [0.41; 2.17] | .983 | | | | |
| ARWIBO-1.5T | Whole sample | HS (38) | A– (59) | no | 0.23 | 0.42 | 1.25 | 1.53 | [0.47; 3.38] | .853 | | | | |
| ARWIBO-1.5T | Whole sample | LP (54) | A– (59) | no | 0.52 | 0.43 | 1.68 | 1.54 | [0.61; 4.61] | .451 | | | | |
| ARWIBO-1.5T | Whole sample | LP (54) | HS (38) | no | 0.29 | 0.48 | 1.34 | 1.62 | [0.43; 4.16] | .819 | | | | |
| NACC-1.5T | Whole sample | HS (167) | A– (332) | no | −0.34 | 0.20 | 0.71 | 1.22 | [0.45; 1.14] | .205 | | | | |
| NACC-1.5T | Whole sample | LP (158) | A– (332) | no | −0.22 | 0.23 | 0.81 | 1.26 | [0.47; 1.38] | .615 | | | | |
| NACC-1.5T | Whole sample | LP (158) | HS (167) | no | 0.12 | 0.24 | 1.13 | 1.27 | [0.65; 1.96] | .860 | | | | |
| **Outcome: APOE ε4 carrier *versus* non-carrier** | | | | | | | | | | | | | | |
| Pooled | Whole sample | HS (1767) | A– (4127) | no | 0.11 | 0.06 | 1.11 | 1.06 | [0.99; 1.26] | | 0.00 | 0.00 | .082 | .129 |
| Pooled | Whole sample | LP (1519) | A– (4127) | no | 0.54 | 0.08 | 1.72 | 1.08 | [1.47; 2.01] | | 0.01 | 11.46 | < .001*** | < .001*** |
| Pooled | Whole sample | LP (1519) | HS (1767) | no | 0.43 | 0.08 | 1.53 | 1.08 | [1.32; 1.78] | | 0.00 | 0.00 | < .001*** | < .001*** |
| Pooled | Aβ-positive | HS (509) | A– (873) | no | 0.12 | 0.12 | 1.13 | 1.13 | [0.89; 1.43] | | 0.00 | 0.00 | .317 | .410 |
| Pooled | Aβ-positive | LP (495) | A– (873) | no | 0.61 | 0.15 | 1.84 | 1.16 | [1.37; 2.48] | | 0.00 | 0.00 | < .001*** | < .001*** |
| Pooled | Aβ-positive | LP (495) | HS (509) | no | 0.51 | 0.15 | 1.66 | 1.16 | [1.23; 2.23] | | 0.00 | 0.00 | < .001*** | .002** |
| A4/LEARN-3T | Whole sample | HS (347) | A– (809) | no | 0.07 | 0.13 | 1.07 | 1.14 | [0.79; 1.46] | .861 | | | | |
| A4/LEARN-3T | Whole sample | LP (81) | A– (809) | no | 0.37 | 0.24 | 1.45 | 1.27 | [0.83; 2.55] | .263 | | | | |
| A4/LEARN-3T | Whole sample | LP (81) | HS (347) | no | 0.30 | 0.25 | 1.36 | 1.29 | [0.75; 2.46] | .453 | | | | |
| A4/LEARN-3T | Aβ-positive | HS (191) | A– (458) | no | 0.04 | 0.17 | 1.04 | 1.19 | [0.69; 1.57] | .968 | | | | |
| A4/LEARN-3T | Aβ-positive | LP (56) | A– (458) | no | 0.37 | 0.30 | 1.44 | 1.35 | [0.72; 2.89] | .435 | | | | |
| A4/LEARN-3T | Aβ-positive | LP (56) | HS (191) | no | 0.32 | 0.32 | 1.38 | 1.37 | [0.66; 2.91] | .566 | | | | |
| ADNI-3T | Whole sample | HS (273) | A– (501) | no | 0.26 | 0.16 | 1.30 | 1.17 | [0.89; 1.89] | .234 | | | | |

| Sample | Sample | Subtype 1 (*n*) | Subtype 2 (*n*) | Used Firth's penalization | log(*OR*) | *SE* | *OR* | *SE* | 95% C.I. | *p* | $\tau^2$ | $I^2$ | Pooled $p_{raw}$ | Pooled $p_{FDR}$ |
|---|---|---|---|---|---|---|---|---|---|---|---|---|---|---|
| ADNI-3T | Whole sample | LP (207) | A– (501) | no | 0.74 | 0.19 | 2.10 | 1.21 | [1.33; 3.32] | < .001*** | | | | |
| ADNI-3T | Whole sample | LP (207) | HS (273) | no | 0.48 | 0.20 | 1.62 | 1.22 | [1.01; 2.60] | .044* | | | | |
| ADNI-3T | Aβ-positive | HS (141) | A– (162) | no | 0.30 | 0.25 | 1.35 | 1.29 | [0.75; 2.45] | .454 | | | | |
| ADNI-3T | Aβ-positive | LP (144) | A– (162) | no | 0.77 | 0.29 | 2.17 | 1.34 | [1.09; 4.31] | .022* | | | | |
| ADNI-3T | Aβ-positive | LP (144) | HS (141) | no | 0.47 | 0.28 | 1.60 | 1.33 | [0.82; 3.12] | .219 | | | | |
| DELCODE-3T | Whole sample | HS (254) | A– (392) | no | 0.03 | 0.18 | 1.04 | 1.20 | [0.68; 1.58] | .980 | | | | |
| DELCODE-3T | Whole sample | LP (265) | A– (392) | no | 0.51 | 0.19 | 1.67 | 1.21 | [1.07; 2.62] | .021* | | | | |
| DELCODE-3T | Whole sample | LP (265) | HS (254) | no | 0.48 | 0.20 | 1.61 | 1.22 | [1.02; 2.56] | .039* | | | | |
| DELCODE-3T | Aβ-positive | HS (71) | A– (77) | no | 0.20 | 0.35 | 1.22 | 1.42 | [0.54; 2.77] | .838 | | | | |
| DELCODE-3T | Aβ-positive | LP (132) | A– (77) | no | 0.67 | 0.36 | 1.95 | 1.43 | [0.85; 4.48] | .146 | | | | |
| DELCODE-3T | Aβ-positive | LP (132) | HS (71) | no | 0.47 | 0.33 | 1.60 | 1.39 | [0.74; 3.44] | .323 | | | | |
| NACC-3T | Whole sample | HS (555) | A– (1852) | no | 0.03 | 0.10 | 1.03 | 1.11 | [0.80; 1.31] | .963 | | | | |
| NACC-3T | Whole sample | LP (611) | A– (1852) | no | 0.40 | 0.11 | 1.49 | 1.11 | [1.16; 1.93] | < .001*** | | | | |
| NACC-3T | Whole sample | LP (611) | HS (555) | no | 0.37 | 0.13 | 1.45 | 1.13 | [1.08; 1.95] | .008** | | | | |
| NACC-3T | Aβ-positive | HS (50) | A– (132) | no | 0.33 | 0.35 | 1.39 | 1.43 | [0.61; 3.19] | .623 | | | | |
| NACC-3T | Aβ-positive | LP (76) | A– (132) | no | 0.51 | 0.36 | 1.67 | 1.44 | [0.71; 3.91] | .340 | | | | |
| NACC-3T | Aβ-positive | LP (76) | HS (50) | no | 0.18 | 0.41 | 1.20 | 1.51 | [0.46; 3.14] | .898 | | | | |
| ADNI-1.5T | Whole sample | HS (170) | A– (236) | no | 0.32 | 0.22 | 1.38 | 1.25 | [0.82; 2.33] | .321 | | | | |
| ADNI-1.5T | Whole sample | LP (200) | A– (236) | no | 0.81 | 0.23 | 2.26 | 1.26 | [1.31; 3.89] | .001** | | | | |
| ADNI-1.5T | Whole sample | LP (200) | HS (170) | no | 0.49 | 0.22 | 1.64 | 1.25 | [0.97; 2.77] | .072 | | | | |
| ADNI-1.5T | Aβ-positive | HS (56) | A– (44) | no | –0.39 | 0.44 | 0.68 | 1.55 | [0.24; 1.90] | .650 | | | | |
| ADNI-1.5T | Aβ-positive | LP (87) | A– (44) | no | 0.86 | 0.47 | 2.37 | 1.59 | [0.79; 7.08] | .154 | | | | |
| ADNI-1.5T | Aβ-positive | LP (87) | HS (56) | no | 1.25 | 0.41 | 3.50 | 1.50 | [1.35; 9.09] | .006** | | | | |
| ARWIBO-1.5T | Whole sample | HS (8) | A– (16) | no | 0.73 | 1.29 | 2.07 | 3.65 | [0.10; 42.99] | .841 | | | | |
| ARWIBO-1.5T | Whole sample | LP (4) | A– (16) | no | 2.34 | 1.33 | 10.33 | 3.79 | [0.45; 234.98] | .186 | | | | |
| ARWIBO-1.5T | Whole sample | LP (4) | HS (8) | no | 1.61 | 1.38 | 5.00 | 3.97 | [0.20; 126.47] | .473 | | | | |
| NACC-1.5T | Whole sample | HS (160) | A– (321) | no | 0.15 | 0.21 | 1.17 | 1.23 | [0.72; 1.89] | .735 | | | | |
| NACC-1.5T | Whole sample | LP (151) | A– (321) | no | 0.62 | 0.24 | 1.86 | 1.27 | [1.07; 3.25] | .024* | | | | |
| NACC-1.5T | Whole sample | LP (151) | HS (160) | no | 0.47 | 0.24 | 1.60 | 1.28 | [0.90; 2.84] | .135 | | | | |
| **Outcome: Aβ positive *versus* negative** | | | | | | | | | | | | | | |
| Pooled | Whole sample | HS (1037) | A– (2123) | no | 0.30 | 0.17 | 1.35 | 1.19 | [0.96; 1.90] | | 0.10 | 70.39 | .090 | .129 |
| Pooled | Whole sample | LP (727) | A– (2123) | no | 0.64 | 0.14 | 1.89 | 1.15 | [1.43; 2.48] | | 0.02 | 25.42 | < .001*** | < .001*** |
| Pooled | Whole sample | LP (727) | HS (1037) | no | 0.31 | 0.13 | 1.37 | 1.13 | [1.07; 1.75] | | 0.00 | 0.00 | .013* | .022* |
| A4/LEARN-3T | Whole sample | HS (347) | A– (809) | no | –0.12 | 0.16 | 0.89 | 1.17 | [0.61; 1.29] | .737 | | | | |
| A4/LEARN-3T | Whole sample | LP (81) | A– (809) | no | 0.30 | 0.30 | 1.34 | 1.35 | [0.66; 2.72] | .589 | | | | |
| A4/LEARN-3T | Whole sample | LP (81) | HS (347) | no | 0.41 | 0.32 | 1.51 | 1.37 | [0.72; 3.19] | .396 | | | | |
| ADNI-3T | Whole sample | HS (268) | A– (490) | no | 0.55 | 0.17 | 1.73 | 1.18 | [1.17; 2.56] | .003** | | | | |
| ADNI-3T | Whole sample | LP (200) | A– (490) | no | 0.83 | 0.21 | 2.29 | 1.24 | [1.40; 3.76] | < .001*** | | | | |
| ADNI-3T | Whole sample | LP (200) | HS (268) | no | 0.28 | 0.22 | 1.32 | 1.25 | [0.79; 2.22] | .418 | | | | |
| DELCODE-3T | Whole sample | HS (211) | A– (309) | no | 0.21 | 0.22 | 1.23 | 1.24 | [0.74; 2.05] | .593 | | | | |
| DELCODE-3T | Whole sample | LP (212) | A– (309) | no | 0.68 | 0.23 | 1.97 | 1.26 | [1.15; 3.37] | .009** | | | | |
| DELCODE-3T | Whole sample | LP (212) | HS (211) | no | 0.47 | 0.24 | 1.60 | 1.27 | [0.92; 2.78] | .119 | | | | |
| NACC-3T | Whole sample | HS (139) | A– (403) | no | 0.10 | 0.23 | 1.11 | 1.26 | [0.65; 1.91] | .893 | | | | |
| NACC-3T | Whole sample | LP (129) | A– (403) | no | 0.22 | 0.30 | 1.24 | 1.35 | [0.61; 2.52] | .749 | | | | |
| NACC-3T | Whole sample | LP (129) | HS (139) | no | 0.11 | 0.34 | 1.12 | 1.40 | [0.51; 2.46] | .938 | | | | |
| ADNI-1.5T | Whole sample | HS (72) | A– (112) | no | 1.05 | 0.37 | 2.87 | 1.45 | [1.20; 6.90] | .013* | | | | |
| ADNI-1.5T | Whole sample | LP (105) | A– (112) | no | 1.14 | 0.36 | 3.12 | 1.44 | [1.33; 7.32] | .005** | | | | |
| ADNI-1.5T | Whole sample | LP (105) | HS (72) | no | 0.08 | 0.41 | 1.09 | 1.50 | [0.42; 2.83] | .977 | | | | |
| **Outcome: MCI *versus* CU diagnosis** | | | | | | | | | | | | | | |
| Pooled | Whole sample | HS (1323) | A– (3724) | no | 0.68 | 0.13 | 1.97 | 1.14 | [1.51; 2.55] | | 0.05 | 54.89 | < .001*** | < .001*** |
| Pooled | Whole sample | LP (1003) | A– (3724) | no | 1.95 | 0.13 | 7.05 | 1.14 | [5.49; 9.05] | | 0.03 | 34.93 | < .001*** | < .001*** |
| Pooled | Whole sample | LP (1003) | HS (1323) | no | 1.24 | 0.10 | 3.47 | 1.10 | [2.86; 4.20] | | 0.00 | 0.00 | < .001*** | < .001*** |
| Pooled | Aβ-positive | HS (242) | A– (409) | no | 0.70 | 0.18 | 2.02 | 1.20 | [1.41; 2.88] | | 0.00 | 0.00 | < .001*** | < .001*** |
| Pooled | Aβ-positive | LP (243) | A– (409) | no | 2.00 | 0.22 | 7.36 | 1.24 | [4.81; 11.25] | | 0.00 | 0.00 | < .001*** | < .001*** |
| Pooled | Aβ-positive | LP (243) | HS (242) | no | 1.28 | 0.27 | 3.60 | 1.32 | [2.10; 6.16] | | 0.08 | 26.30 | < .001*** | < .001*** |
| ADNI-3T | Whole sample | HS (247) | A– (523) | no | 0.43 | 0.16 | 1.54 | 1.17 | [1.06; 2.23] | .020* | | | | |
| ADNI-3T | Whole sample | LP (134) | A– (523) | no | 1.97 | 0.25 | 7.17 | 1.28 | [4.01; 12.83] | < .001*** | | | | |
| ADNI-3T | Whole sample | LP (134) | HS (247) | no | 1.54 | 0.27 | 4.67 | 1.30 | [2.50; 8.70] | < .001*** | | | | |

| Sample | Sample | Subtype 1 (*n*) | Subtype 2 (*n*) | Used Firth's penalization | log(*OR*) | *SE* | *OR* | *SE* | 95% C.I. | *p* | $\tau^2$ | $I^2$ | Pooled $p_{raw}$ | Pooled $p_{FDR}$ |
|---|---|---|---|---|---|---|---|---|---|---|---|---|---|---|
| ADNI-3T | Aβ-positive | HS (105) | A– (150) | no | 0.84 | 0.28 | 2.32 | 1.32 | [1.21; 4.44] | .007** | | | | |
| ADNI-3T | Aβ-positive | LP (75) | A– (150) | no | 2.28 | 0.41 | 9.75 | 1.50 | [3.76; 25.29] | < .001*** | | | | |
| ADNI-3T | Aβ-positive | LP (75) | HS (105) | no | 1.44 | 0.43 | 4.20 | 1.53 | [1.55; 11.43] | .002** | | | | |
| DELCODE-3T | Whole sample | HS (234) | A– (389) | no | 0.88 | 0.25 | 2.42 | 1.29 | [1.34; 4.36] | .001** | | | | |
| DELCODE-3T | Whole sample | LP (185) | A– (389) | no | 2.01 | 0.24 | 7.45 | 1.27 | [4.28; 12.97] | < .001*** | | | | |
| DELCODE-3T | Whole sample | LP (185) | HS (234) | no | 1.13 | 0.23 | 3.08 | 1.26 | [1.79; 5.31] | < .001*** | | | | |
| DELCODE-3T | Aβ-positive | HS (56) | A– (77) | no | 0.85 | 0.42 | 2.33 | 1.52 | [0.87; 6.22] | .108 | | | | |
| DELCODE-3T | Aβ-positive | LP (72) | A– (77) | no | 1.55 | 0.38 | 4.72 | 1.46 | [1.93; 11.52] | < .001*** | | | | |
| DELCODE-3T | Aβ-positive | LP (72) | HS (56) | no | 0.71 | 0.38 | 2.03 | 1.47 | [0.83; 4.96] | .154 | | | | |
| NACC-3T | Whole sample | HS (573) | A– (2255) | no | 0.61 | 0.12 | 1.84 | 1.12 | [1.40; 2.43] | < .001*** | | | | |
| NACC-3T | Whole sample | LP (492) | A– (2255) | no | 1.75 | 0.11 | 5.78 | 1.12 | [4.47; 7.48] | < .001*** | | | | |
| NACC-3T | Whole sample | LP (492) | HS (573) | no | 1.14 | 0.14 | 3.14 | 1.15 | [2.28; 4.32] | < .001*** | | | | |
| NACC-3T | Aβ-positive | HS (46) | A– (142) | no | 0.42 | 0.36 | 1.52 | 1.44 | [0.65; 3.54] | .483 | | | | |
| NACC-3T | Aβ-positive | LP (47) | A– (142) | no | 2.22 | 0.41 | 9.18 | 1.51 | [3.51; 24.03] | < .001*** | | | | |
| NACC-3T | Aβ-positive | LP (47) | HS (46) | no | 1.80 | 0.48 | 6.06 | 1.62 | [1.95; 18.82] | < .001*** | | | | |
| ADNI-1.5T | Whole sample | HS (121) | A– (224) | no | 1.17 | 0.25 | 3.24 | 1.28 | [1.82; 5.75] | < .001*** | | | | |
| ADNI-1.5T | Whole sample | LP (115) | A– (224) | no | 2.28 | 0.31 | 9.82 | 1.37 | [4.72; 20.46] | < .001*** | | | | |
| ADNI-1.5T | Whole sample | LP (115) | HS (121) | no | 1.11 | 0.34 | 3.04 | 1.41 | [1.35; 6.80] | .004** | | | | |
| ADNI-1.5T | Aβ-positive | HS (35) | A– (40) | no | 0.58 | 0.51 | 1.78 | 1.67 | [0.54; 5.92] | .494 | | | | |
| ADNI-1.5T | Aβ-positive | LP (49) | A– (40) | no | 2.01 | 0.64 | 7.49 | 1.89 | [1.68; 33.49] | .005** | | | | |
| ADNI-1.5T | Aβ-positive | LP (49) | HS (35) | no | 1.43 | 0.68 | 4.20 | 1.97 | [0.85; 20.66] | .088 | | | | |
| ARWIBO-1.5T | Whole sample | HS (34) | A– (52) | no | –0.59 | 0.58 | 0.56 | 1.79 | [0.14; 2.19] | .573 | | | | |
| ARWIBO-1.5T | Whole sample | LP (26) | A– (52) | no | 1.08 | 0.55 | 2.94 | 1.73 | [0.81; 10.67] | .123 | | | | |
| ARWIBO-1.5T | Whole sample | LP (26) | HS (34) | no | 1.67 | 0.65 | 5.29 | 1.91 | [1.16; 24.00] | .027* | | | | |
| NACC-1.5T | Whole sample | HS (114) | A– (281) | no | 0.81 | 0.26 | 2.25 | 1.30 | [1.22; 4.16] | .005** | | | | |
| NACC-1.5T | Whole sample | LP (51) | A– (281) | no | 2.56 | 0.36 | 12.93 | 1.44 | [5.53; 30.26] | < .001*** | | | | |
| NACC-1.5T | Whole sample | LP (51) | HS (114) | no | 1.75 | 0.38 | 5.74 | 1.47 | [2.33; 14.13] | < .001*** | | | | |
| **Outcome: DAT *versus* CU/MCI diagnosis** | | | | | | | | | | | | | | |
| Pooled | Whole sample | HS (1616) | A– (3876) | no | 1.55 | 0.32 | 4.70 | 1.38 | [2.50; 8.83] | | 0.48 | 84.59 | < .001*** | < .001*** |
| Pooled | Whole sample | LP (1686) | A– (3876) | no | 2.77 | 0.18 | 15.98 | 1.20 | [11.19; 22.83] | | 0.10 | 56.74 | < .001*** | < .001*** |
| Pooled | Whole sample | LP (1686) | HS (1616) | no | 1.19 | 0.17 | 3.28 | 1.19 | [2.34; 4.59] | | 0.11 | 68.78 | < .001*** | < .001*** |
| Pooled | Aβ-positive | HS (334) | A– (435) | no | 1.62 | 0.24 | 5.03 | 1.27 | [3.14; 8.07] | | 0.00 | 0.00 | < .001*** | < .001*** |
| Pooled | Aβ-positive | LP (462) | A– (435) | no | 2.47 | 0.23 | 11.85 | 1.25 | [7.60; 18.48] | | 0.00 | 0.00 | < .001*** | < .001*** |
| Pooled | Aβ-positive | LP (462) | HS (334) | no | 0.81 | 0.16 | 2.24 | 1.17 | [1.64; 3.06] | | 0.00 | 0.00 | < .001*** | < .001*** |
| ADNI-3T | Whole sample | HS (289) | A– (538) | no | 1.72 | 0.31 | 5.61 | 1.36 | [2.73; 11.50] | < .001*** | | | | |
| ADNI-3T | Whole sample | LP (225) | A– (538) | no | 3.08 | 0.29 | 21.68 | 1.34 | [10.95; 42.92] | < .001*** | | | | |
| ADNI-3T | Whole sample | LP (225) | HS (289) | no | 1.35 | 0.22 | 3.87 | 1.24 | [2.33; 6.43] | < .001*** | | | | |
| ADNI-3T | Aβ-positive | HS (143) | A– (162) | no | 1.50 | 0.35 | 4.46 | 1.42 | [1.96; 10.14] | < .001*** | | | | |
| ADNI-3T | Aβ-positive | LP (145) | A– (162) | no | 2.40 | 0.34 | 11.06 | 1.40 | [5.00; 24.48] | < .001*** | | | | |
| ADNI-3T | Aβ-positive | LP (145) | HS (143) | no | 0.91 | 0.26 | 2.48 | 1.29 | [1.36; 4.51] | .001** | | | | |
| DELCODE-3T | Whole sample | HS (254) | A– (392) | no | 2.36 | 0.58 | 10.60 | 1.79 | [2.71; 41.53] | < .001*** | | | | |
| DELCODE-3T | Whole sample | LP (265) | A– (392) | no | 3.68 | 0.55 | 39.68 | 1.74 | [10.88; 144.74] | < .001*** | | | | |
| DELCODE-3T | Whole sample | LP (265) | HS (254) | no | 1.32 | 0.27 | 3.74 | 1.32 | [1.97; 7.13] | < .001*** | | | | |
| DELCODE-3T | Aβ-positive | HS (71) | A– (77) | no | 3.95 | 1.45 | 51.78 | 4.24 | [1.75; 1531.30] | .017* | | | | |
| DELCODE-3T | Aβ-positive | LP (132) | A– (77) | no | 4.78 | 1.42 | 119.27 | 4.15 | [4.25; 3349.95] | .002** | | | | |
| DELCODE-3T | Aβ-positive | LP (132) | HS (71) | no | 0.83 | 0.35 | 2.30 | 1.42 | [1.01; 5.27] | .047* | | | | |
| NACC-3T | Whole sample | HS (698) | A– (2319) | no | 2.07 | 0.16 | 7.95 | 1.17 | [5.45; 11.60] | < .001*** | | | | |
| NACC-3T | Whole sample | LP (784) | A– (2319) | no | 3.00 | 0.15 | 20.16 | 1.16 | [14.30; 28.43] | < .001*** | | | | |
| NACC-3T | Whole sample | LP (784) | HS (698) | no | 0.93 | 0.12 | 2.53 | 1.13 | [1.89; 3.40] | < .001*** | | | | |
| NACC-3T | Aβ-positive | HS (64) | A– (152) | no | 1.62 | 0.42 | 5.06 | 1.53 | [1.87; 13.67] | < .001*** | | | | |
| NACC-3T | Aβ-positive | LP (98) | A– (152) | no | 2.68 | 0.38 | 14.60 | 1.46 | [5.99; 35.58] | < .001*** | | | | |
| NACC-3T | Aβ-positive | LP (98) | HS (64) | no | 1.06 | 0.35 | 2.89 | 1.42 | [1.26; 6.62] | .008** | | | | |
| ADNI-1.5T | Whole sample | HS (170) | A– (236) | no | 2.04 | 0.34 | 7.66 | 1.41 | [3.44; 17.07] | < .001*** | | | | |
| ADNI-1.5T | Whole sample | LP (200) | A– (236) | no | 2.62 | 0.33 | 13.68 | 1.39 | [6.32; 29.63] | < .001*** | | | | |
| ADNI-1.5T | Whole sample | LP (200) | HS (170) | no | 0.58 | 0.23 | 1.79 | 1.25 | [1.05; 3.04] | .028* | | | | |
| ADNI-1.5T | Aβ-positive | HS (56) | A– (44) | no | 1.57 | 0.57 | 4.78 | 1.77 | [1.25; 18.31] | .017* | | | | |
| ADNI-1.5T | Aβ-positive | LP (87) | A– (44) | no | 1.87 | 0.55 | 6.52 | 1.73 | [1.80; 23.56] | .002** | | | | |
| ADNI-1.5T | Aβ-positive | LP (87) | HS (56) | no | 0.31 | 0.36 | 1.36 | 1.43 | [0.59; 3.17] | .665 | | | | |

| **Sample** | **Sample** | **Subtype 1 (*n*)** | **Subtype 2 (*n*)** | **Used Firth's penalization** | **log(*OR*)** | ***SE*** | ***OR*** | ***SE*** | **95% C.I.** | ***p*** | **$\tau^2$** | **$I^2$** | **Pooled $p_{raw}$** | **Pooled $p_{FDR}$** |
|---|---|---|---|---|---|---|---|---|---|---|---|---|---|---|
| ARWIBO-1.5T | Whole sample | HS (38) | A– (59) | no | –0.07 | 0.64 | 0.93 | 1.89 | [0.21; 4.17] | .994 | | | | |
| ARWIBO-1.5T | Whole sample | LP (54) | A– (59) | no | 1.89 | 0.48 | 6.63 | 1.62 | [2.14; 20.49] | < .001*** | | | | |
| ARWIBO-1.5T | Whole sample | LP (54) | HS (38) | no | 1.96 | 0.58 | 7.10 | 1.78 | [1.84; 27.46] | .002** | | | | |
| NACC-1.5T | Whole sample | HS (167) | A– (332) | no | 0.84 | 0.23 | 2.31 | 1.26 | [1.35; 3.94] | < .001*** | | | | |
| NACC-1.5T | Whole sample | LP (158) | A– (332) | no | 2.40 | 0.23 | 11.07 | 1.26 | [6.46; 18.96] | < .001*** | | | | |
| NACC-1.5T | Whole sample | LP (158) | HS (167) | no | 1.57 | 0.24 | 4.80 | 1.27 | [2.72; 8.48] | < .001*** | | | | |

*$p < .05$. **$p < .01$. ***$p < .001$. Abbreviations: A–, atrophy negative. HS, hippocampal-sparing. LP, limbic-predominant.

**Supplementary Table 5 Summary statistics of cross-sectional stage effects derived from logistic regression models predicting binary outcome variables.**

| Sample | Sample | Subtype (*n*) | Used Firth's penalization | log(*OR*) | *SE* | *OR* | *SE* | 95% C.I. | *p* | τ² | I² | Pooled $p_{raw}$ | Pooled $p_{FDR}$ |
|---|---|---|---|---|---|---|---|---|---|---|---|---|---|
| **Outcome: female *versus* male sex** | | | | | | | | | | | | | |
| Pooled | Whole sample | HS (3926) | | –0.01 | 0.01 | 0.99 | 1.01 | [0.96; 1.02] | | 0.00 | 0.00 | .604 | .604 |
| Pooled | Whole sample | LP (3534) | | –0.02 | 0.02 | 0.98 | 1.02 | [0.95; 1.01] | | 0.00 | 0.00 | .282 | .442 |
| Pooled | Aβ-positive | HS (1050) | | 0.02 | 0.03 | 1.02 | 1.03 | [0.96; 1.09] | | 0.00 | 0.00 | .439 | .488 |
| Pooled | Aβ-positive | LP (1036) | | 0.01 | 0.03 | 1.01 | 1.03 | [0.95; 1.06] | | 0.00 | 0.00 | .851 | .946 |
| A4/LEARN-3T | Whole sample | HS (347) | no | 0.16 | 0.08 | 1.17 | 1.08 | [1.00; 1.37] | .051 | | | | |
| A4/LEARN-3T | Whole sample | HS (347) | no | 0.16 | 0.08 | 1.17 | 1.08 | [1.00; 1.37] | .051 | | | | |
| A4/LEARN-3T | Whole sample | LP (81) | no | 0.00 | 0.28 | 1.00 | 1.33 | [0.57; 1.73] | .991 | | | | |
| A4/LEARN-3T | Whole sample | LP (81) | no | 0.00 | 0.28 | 1.00 | 1.33 | [0.57; 1.73] | .991 | | | | |
| A4/LEARN-3T | Aβ-positive | HS (191) | no | 0.14 | 0.10 | 1.15 | 1.11 | [0.94; 1.41] | .166 | | | | |
| A4/LEARN-3T | Aβ-positive | HS (191) | no | 0.14 | 0.10 | 1.15 | 1.11 | [0.94; 1.41] | .166 | | | | |
| A4/LEARN-3T | Aβ-positive | LP (56) | no | 0.03 | 0.31 | 1.03 | 1.36 | [0.56; 1.87] | .933 | | | | |
| A4/LEARN-3T | Aβ-positive | LP (56) | no | 0.03 | 0.31 | 1.03 | 1.36 | [0.56; 1.87] | .933 | | | | |
| ADNI-3T | Whole sample | HS (289) | no | –0.04 | 0.06 | 0.96 | 1.07 | [0.85; 1.09] | .569 | | | | |
| ADNI-3T | Whole sample | HS (289) | no | –0.04 | 0.06 | 0.96 | 1.07 | [0.85; 1.09] | .569 | | | | |
| ADNI-3T | Whole sample | LP (225) | no | 0.03 | 0.05 | 1.03 | 1.06 | [0.93; 1.15] | .550 | | | | |
| ADNI-3T | Whole sample | LP (225) | no | 0.03 | 0.05 | 1.03 | 1.06 | [0.93; 1.15] | .550 | | | | |
| ADNI-3T | Aβ-positive | HS (143) | no | –0.04 | 0.08 | 0.96 | 1.08 | [0.83; 1.12] | .640 | | | | |
| ADNI-3T | Aβ-positive | HS (143) | no | –0.04 | 0.08 | 0.96 | 1.08 | [0.83; 1.12] | .640 | | | | |
| ADNI-3T | Aβ-positive | LP (145) | no | 0.02 | 0.06 | 1.02 | 1.06 | [0.90; 1.15] | .735 | | | | |
| ADNI-3T | Aβ-positive | LP (145) | no | 0.02 | 0.06 | 1.02 | 1.06 | [0.90; 1.15] | .735 | | | | |
| DELCODE-3T | Whole sample | HS (254) | no | –0.05 | 0.07 | 0.95 | 1.08 | [0.82; 1.10] | .531 | | | | |
| DELCODE-3T | Whole sample | HS (254) | no | –0.05 | 0.07 | 0.95 | 1.08 | [0.82; 1.10] | .531 | | | | |
| DELCODE-3T | Whole sample | LP (265) | no | –0.05 | 0.06 | 0.95 | 1.06 | [0.84; 1.08] | .440 | | | | |
| DELCODE-3T | Whole sample | LP (265) | no | –0.05 | 0.06 | 0.95 | 1.06 | [0.84; 1.08] | .440 | | | | |
| DELCODE-3T | Aβ-positive | HS (71) | no | –0.07 | 0.12 | 0.93 | 1.13 | [0.74; 1.18] | .569 | | | | |
| DELCODE-3T | Aβ-positive | HS (71) | no | –0.07 | 0.12 | 0.93 | 1.13 | [0.74; 1.18] | .569 | | | | |
| DELCODE-3T | Aβ-positive | LP (132) | no | –0.03 | 0.08 | 0.97 | 1.09 | [0.83; 1.15] | .763 | | | | |
| DELCODE-3T | Aβ-positive | LP (132) | no | –0.03 | 0.08 | 0.97 | 1.09 | [0.83; 1.15] | .763 | | | | |
| NACC-3T | Whole sample | HS (698) | no | 0.01 | 0.04 | 1.01 | 1.04 | [0.93; 1.10] | .799 | | | | |
| NACC-3T | Whole sample | HS (698) | no | 0.01 | 0.04 | 1.01 | 1.04 | [0.93; 1.10] | .799 | | | | |
| NACC-3T | Whole sample | LP (784) | no | –0.02 | 0.03 | 0.98 | 1.03 | [0.92; 1.04] | .501 | | | | |
| NACC-3T | Whole sample | LP (784) | no | –0.02 | 0.03 | 0.98 | 1.03 | [0.92; 1.04] | .501 | | | | |
| NACC-3T | Aβ-positive | HS (64) | no | –0.07 | 0.12 | 0.94 | 1.13 | [0.74; 1.19] | .598 | | | | |
| NACC-3T | Aβ-positive | HS (64) | no | –0.07 | 0.12 | 0.94 | 1.13 | [0.74; 1.19] | .598 | | | | |
| NACC-3T | Aβ-positive | LP (98) | no | 0.09 | 0.11 | 1.10 | 1.12 | [0.88; 1.36] | .402 | | | | |
| NACC-3T | Aβ-positive | LP (98) | no | 0.09 | 0.11 | 1.10 | 1.12 | [0.88; 1.36] | .402 | | | | |
| ADNI–1.5T | Whole sample | HS (170) | no | 0.02 | 0.06 | 1.02 | 1.06 | [0.92; 1.14] | .702 | | | | |
| ADNI–1.5T | Whole sample | HS (170) | no | 0.02 | 0.06 | 1.02 | 1.06 | [0.92; 1.14] | .702 | | | | |
| ADNI–1.5T | Whole sample | LP (200) | no | –0.05 | 0.06 | 0.95 | 1.06 | [0.84; 1.07] | .375 | | | | |
| ADNI–1.5T | Whole sample | LP (200) | no | –0.05 | 0.06 | 0.95 | 1.06 | [0.84; 1.07] | .375 | | | | |
| ADNI–1.5T | Aβ-positive | HS (56) | no | 0.12 | 0.09 | 1.13 | 1.10 | [0.94; 1.35] | .192 | | | | |
| ADNI–1.5T | Aβ-positive | HS (56) | no | 0.12 | 0.09 | 1.13 | 1.10 | [0.94; 1.35] | .192 | | | | |
| ADNI–1.5T | Aβ-positive | LP (87) | yes | –0.06 | 0.09 | 0.94 | 1.10 | [0.78; 1.13] | .541 | | | | |
| ADNI–1.5T | Aβ-positive | LP (87) | yes | –0.06 | 0.09 | 0.94 | 1.10 | [0.78; 1.13] | .541 | | | | |
| ARWIBO–1.5T | Whole sample | HS (38) | no | –0.03 | 0.20 | 0.97 | 1.22 | [0.65; 1.43] | .867 | | | | |
| ARWIBO–1.5T | Whole sample | HS (38) | no | –0.03 | 0.20 | 0.97 | 1.22 | [0.65; 1.43] | .867 | | | | |
| ARWIBO–1.5T | Whole sample | LP (54) | no | –0.07 | 0.13 | 0.93 | 1.13 | [0.73; 1.19] | .582 | | | | |
| ARWIBO–1.5T | Whole sample | LP (54) | no | –0.07 | 0.13 | 0.93 | 1.13 | [0.73; 1.19] | .582 | | | | |
| NACC–1.5T | Whole sample | HS (167) | no | –0.04 | 0.03 | 0.96 | 1.03 | [0.90; 1.02] | .210 | | | | |
| NACC–1.5T | Whole sample | HS (167) | no | –0.04 | 0.03 | 0.96 | 1.03 | [0.90; 1.02] | .210 | | | | |
| NACC–1.5T | Whole sample | LP (158) | no | 0.03 | 0.07 | 1.03 | 1.07 | [0.90; 1.17] | .683 | | | | |
| NACC–1.5T | Whole sample | LP (158) | no | 0.03 | 0.07 | 1.03 | 1.07 | [0.90; 1.17] | .683 | | | | |
| **Outcome: APOE ε4 carrier *versus* non-carrier** | | | | | | | | | | | | | |
| Pooled | Whole sample | HS (3526) | | 0.06 | 0.02 | 1.06 | 1.02 | [1.03; 1.09] | | 0.00 | 0.00 | < .001*** | < .001*** |
| Pooled | Whole sample | LP (3030) | | 0.01 | 0.02 | 1.01 | 1.02 | [0.97; 1.05] | | 0.00 | 14.75 | .530 | .648 |
| Pooled | Aβ-positive | HS (1018) | | 0.05 | 0.03 | 1.05 | 1.03 | [0.98; 1.12] | | 0.00 | 0.02 | .134 | .269 |

| Sample | Sample | Subtype (*n*) | Used Firth's penalization | log(*OR*) | *SE* | *OR* | *SE* | 95% C.I. | *p* | $\tau^2$ | $I^2$ | Pooled $p_{raw}$ | Pooled $p_{FDR}$ |
|---|---|---|---|---|---|---|---|---|---|---|---|---|---|
| Pooled | Aβ-positive | LP (990) | | 0.01 | 0.04 | 1.01 | 1.04 | [0.94; 1.08] | | 0.00 | 0.01 | .747 | .946 |
| A4/LEARN-3T | Whole sample | HS (347) | no | 0.06 | 0.08 | 1.06 | 1.08 | [0.91; 1.24] | .463 | | | | |
| A4/LEARN-3T | Whole sample | HS (347) | no | 0.06 | 0.08 | 1.06 | 1.08 | [0.91; 1.24] | .463 | | | | |
| A4/LEARN-3T | Whole sample | LP (81) | no | 0.42 | 0.30 | 1.52 | 1.35 | [0.84; 2.75] | .168 | | | | |
| A4/LEARN-3T | Whole sample | LP (81) | no | 0.42 | 0.30 | 1.52 | 1.35 | [0.84; 2.75] | .168 | | | | |
| A4/LEARN-3T | Aβ-positive | HS (191) | no | 0.00 | 0.10 | 1.00 | 1.10 | [0.82; 1.22] | .986 | | | | |
| A4/LEARN-3T | Aβ-positive | HS (191) | no | 0.00 | 0.10 | 1.00 | 1.10 | [0.82; 1.22] | .986 | | | | |
| A4/LEARN-3T | Aβ-positive | LP (56) | no | 0.66 | 0.49 | 1.94 | 1.63 | [0.75; 5.03] | .175 | | | | |
| A4/LEARN-3T | Aβ-positive | LP (56) | no | 0.66 | 0.49 | 1.94 | 1.63 | [0.75; 5.03] | .175 | | | | |
| ADNI-3T | Whole sample | HS (273) | no | 0.11 | 0.07 | 1.11 | 1.07 | [0.98; 1.27] | .111 | | | | |
| ADNI-3T | Whole sample | HS (273) | no | 0.11 | 0.07 | 1.11 | 1.07 | [0.98; 1.27] | .111 | | | | |
| ADNI-3T | Whole sample | LP (207) | no | 0.06 | 0.06 | 1.06 | 1.07 | [0.94; 1.21] | .337 | | | | |
| ADNI-3T | Whole sample | LP (207) | no | 0.06 | 0.06 | 1.06 | 1.07 | [0.94; 1.21] | .337 | | | | |
| ADNI-3T | Aβ-positive | HS (141) | no | 0.07 | 0.09 | 1.07 | 1.09 | [0.90; 1.27] | .434 | | | | |
| ADNI-3T | Aβ-positive | HS (141) | no | 0.07 | 0.09 | 1.07 | 1.09 | [0.90; 1.27] | .434 | | | | |
| ADNI-3T | Aβ-positive | LP (144) | no | 0.02 | 0.08 | 1.02 | 1.09 | [0.87; 1.20] | .793 | | | | |
| ADNI-3T | Aβ-positive | LP (144) | no | 0.02 | 0.08 | 1.02 | 1.09 | [0.87; 1.20] | .793 | | | | |
| DELCODE-3T | Whole sample | HS (254) | no | 0.19 | 0.08 | 1.21 | 1.08 | [1.04; 1.41] | .013* | | | | |
| DELCODE-3T | Whole sample | HS (254) | no | 0.19 | 0.08 | 1.21 | 1.08 | [1.04; 1.41] | .013* | | | | |
| DELCODE-3T | Whole sample | LP (265) | no | −0.04 | 0.06 | 0.96 | 1.07 | [0.85; 1.09] | .545 | | | | |
| DELCODE-3T | Whole sample | LP (265) | no | −0.04 | 0.06 | 0.96 | 1.07 | [0.85; 1.09] | .545 | | | | |
| DELCODE-3T | Aβ-positive | HS (71) | no | 0.24 | 0.13 | 1.27 | 1.14 | [0.98; 1.64] | .071 | | | | |
| DELCODE-3T | Aβ-positive | HS (71) | no | 0.24 | 0.13 | 1.27 | 1.14 | [0.98; 1.64] | .071 | | | | |
| DELCODE-3T | Aβ-positive | LP (132) | no | 0.00 | 0.09 | 1.00 | 1.09 | [0.83; 1.19] | .973 | | | | |
| DELCODE-3T | Aβ-positive | LP (132) | no | 0.00 | 0.09 | 1.00 | 1.09 | [0.83; 1.19] | .973 | | | | |
| NACC-3T | Whole sample | HS (555) | no | 0.00 | 0.05 | 1.00 | 1.05 | [0.92; 1.10] | .918 | | | | |
| NACC-3T | Whole sample | HS (555) | no | 0.00 | 0.05 | 1.00 | 1.05 | [0.92; 1.10] | .918 | | | | |
| NACC-3T | Whole sample | LP (611) | no | 0.04 | 0.04 | 1.04 | 1.04 | [0.96; 1.12] | .328 | | | | |
| NACC-3T | Whole sample | LP (611) | no | 0.04 | 0.04 | 1.04 | 1.04 | [0.96; 1.12] | .328 | | | | |
| NACC-3T | Aβ-positive | HS (50) | no | −0.21 | 0.14 | 0.81 | 1.15 | [0.61; 1.08] | .149 | | | | |
| NACC-3T | Aβ-positive | HS (50) | no | −0.21 | 0.14 | 0.81 | 1.15 | [0.61; 1.08] | .149 | | | | |
| NACC-3T | Aβ-positive | LP (76) | no | 0.07 | 0.13 | 1.07 | 1.14 | [0.83; 1.38] | .602 | | | | |
| NACC-3T | Aβ-positive | LP (76) | no | 0.07 | 0.13 | 1.07 | 1.14 | [0.83; 1.38] | .602 | | | | |
| ADNI–1.5T | Whole sample | HS (170) | no | 0.07 | 0.05 | 1.07 | 1.05 | [0.96; 1.19] | .208 | | | | |
| ADNI–1.5T | Whole sample | HS (170) | no | 0.07 | 0.05 | 1.07 | 1.05 | [0.96; 1.19] | .208 | | | | |
| ADNI–1.5T | Whole sample | LP (200) | no | −0.11 | 0.07 | 0.89 | 1.07 | [0.79; 1.02] | .088 | | | | |
| ADNI–1.5T | Whole sample | LP (200) | no | −0.11 | 0.07 | 0.89 | 1.07 | [0.79; 1.02] | .088 | | | | |
| ADNI–1.5T | Aβ-positive | HS (56) | no | 0.09 | 0.10 | 1.09 | 1.10 | [0.91; 1.32] | .360 | | | | |
| ADNI–1.5T | Aβ-positive | HS (56) | no | 0.09 | 0.10 | 1.09 | 1.10 | [0.91; 1.32] | .360 | | | | |
| ADNI–1.5T | Aβ-positive | LP (87) | yes | −0.09 | 0.13 | 0.92 | 1.14 | [0.71; 1.22] | .525 | | | | |
| ADNI–1.5T | Aβ-positive | LP (87) | yes | −0.09 | 0.13 | 0.92 | 1.14 | [0.71; 1.22] | .525 | | | | |
| ARWIBO–1.5T | Whole sample | HS (8) | no | −0.41 | 0.83 | 0.66 | 2.30 | [0.13; 3.38] | .619 | | | | |
| NACC–1.5T | Whole sample | HS (160) | no | 0.04 | 0.04 | 1.04 | 1.04 | [0.97; 1.12] | .260 | | | | |
| NACC–1.5T | Whole sample | HS (160) | no | 0.04 | 0.04 | 1.04 | 1.04 | [0.97; 1.12] | .260 | | | | |
| NACC–1.5T | Whole sample | LP (151) | no | 0.08 | 0.08 | 1.08 | 1.08 | [0.93; 1.25] | .321 | | | | |
| NACC–1.5T | Whole sample | LP (151) | no | 0.08 | 0.08 | 1.08 | 1.08 | [0.93; 1.25] | .321 | | | | |
| **Outcome: Aβ positive *versus* negative** | | | | | | | | | | | | | |
| Pooled | Whole sample | HS (2074) | | 0.18 | 0.03 | 1.20 | 1.03 | [1.13; 1.28] | | 0.00 | 0.00 | < .001*** | < .001*** |
| Pooled | Whole sample | LP (1454) | | 0.00 | 0.03 | 1.00 | 1.03 | [0.94; 1.07] | | 0.00 | 0.00 | .908 | .908 |
| A4/LEARN-3T | Whole sample | HS (347) | no | 0.11 | 0.10 | 1.12 | 1.11 | [0.91; 1.37] | .279 | | | | |
| A4/LEARN-3T | Whole sample | HS (347) | no | 0.11 | 0.10 | 1.12 | 1.11 | [0.91; 1.37] | .279 | | | | |
| A4/LEARN-3T | Whole sample | LP (81) | no | −0.24 | 0.37 | 0.79 | 1.45 | [0.38; 1.63] | .525 | | | | |
| A4/LEARN-3T | Whole sample | LP (81) | no | −0.24 | 0.37 | 0.79 | 1.45 | [0.38; 1.63] | .525 | | | | |
| ADNI-3T | Whole sample | HS (268) | no | 0.27 | 0.09 | 1.31 | 1.09 | [1.10; 1.55] | .002** | | | | |
| ADNI-3T | Whole sample | HS (268) | no | 0.27 | 0.09 | 1.31 | 1.09 | [1.10; 1.55] | .002** | | | | |
| ADNI-3T | Whole sample | LP (200) | no | 0.09 | 0.08 | 1.09 | 1.08 | [0.94; 1.26] | .244 | | | | |
| ADNI-3T | Whole sample | LP (200) | no | 0.09 | 0.08 | 1.09 | 1.08 | [0.94; 1.26] | .244 | | | | |
| DELCODE-3T | Whole sample | HS (211) | no | 0.20 | 0.09 | 1.23 | 1.10 | [1.02; 1.47] | .029* | | | | |

| Sample | Sample | Subtype ($n$) | Used Firth's penalization | log($OR$) | $SE$ | $OR$ | $SE$ | 95% C.I. | $p$ | $\tau^2$ | $I^2$ | Pooled $p_{raw}$ | Pooled $p_{FDR}$ |
|---|---|---|---|---|---|---|---|---|---|---|---|---|---|
| DELCODE-3T | Whole sample | HS (211) | no | 0.20 | 0.09 | 1.23 | 1.10 | [1.02; 1.47] | .029* | | | | |
| DELCODE-3T | Whole sample | LP (212) | no | –0.01 | 0.08 | 0.99 | 1.08 | [0.85; 1.16] | .920 | | | | |
| DELCODE-3T | Whole sample | LP (212) | no | –0.01 | 0.08 | 0.99 | 1.08 | [0.85; 1.16] | .920 | | | | |
| NACC-3T | Whole sample | HS (139) | no | 0.25 | 0.13 | 1.28 | 1.14 | [1.00; 1.66] | .053 | | | | |
| NACC-3T | Whole sample | HS (139) | no | 0.25 | 0.13 | 1.28 | 1.14 | [1.00; 1.66] | .053 | | | | |
| NACC-3T | Whole sample | LP (129) | no | –0.11 | 0.14 | 0.90 | 1.15 | [0.68; 1.19] | .451 | | | | |
| NACC-3T | Whole sample | LP (129) | no | –0.11 | 0.14 | 0.90 | 1.15 | [0.68; 1.19] | .451 | | | | |
| ADNI–1.5T | Whole sample | HS (72) | no | 0.05 | 0.11 | 1.05 | 1.11 | [0.85; 1.30] | .658 | | | | |
| ADNI–1.5T | Whole sample | HS (72) | no | 0.05 | 0.11 | 1.05 | 1.11 | [0.85; 1.30] | .658 | | | | |
| ADNI–1.5T | Whole sample | LP (105) | no | –0.09 | 0.13 | 0.91 | 1.14 | [0.71; 1.17] | .472 | | | | |
| ADNI–1.5T | Whole sample | LP (105) | no | –0.09 | 0.13 | 0.91 | 1.14 | [0.71; 1.17] | .472 | | | | |
| **Outcome: MCI *versus* CU diagnosis** | | | | | | | | | | | | | |
| Pooled | Whole sample | HS (2646) | | 0.22 | 0.04 | 1.24 | 1.05 | [1.14; 1.35] | | 0.01 | 72.60 | < .001*** | < .001*** |
| Pooled | Whole sample | LP (2006) | | 0.38 | 0.06 | 1.46 | 1.06 | [1.31; 1.64] | | 0.01 | 36.17 | < .001*** | < .001*** |
| Pooled | Aβ-positive | HS (484) | | 0.26 | 0.06 | 1.29 | 1.06 | [1.15; 1.45] | | 0.00 | 0.00 | < .001*** | < .001*** |
| Pooled | Aβ-positive | LP (486) | | 0.24 | 0.07 | 1.27 | 1.08 | [1.09; 1.47] | | 0.00 | 0.00 | .002** | .005** |
| ADNI-3T | Whole sample | HS (247) | no | 0.28 | 0.09 | 1.32 | 1.09 | [1.11; 1.56] | .001** | | | | |
| ADNI-3T | Whole sample | HS (247) | no | 0.28 | 0.09 | 1.32 | 1.09 | [1.11; 1.56] | .001** | | | | |
| ADNI-3T | Whole sample | LP (134) | no | 0.21 | 0.14 | 1.23 | 1.15 | [0.93; 1.63] | .144 | | | | |
| ADNI-3T | Whole sample | LP (134) | no | 0.21 | 0.14 | 1.23 | 1.15 | [0.93; 1.63] | .144 | | | | |
| ADNI-3T | Aβ-positive | HS (105) | no | 0.28 | 0.13 | 1.33 | 1.13 | [1.04; 1.70] | .025* | | | | |
| ADNI-3T | Aβ-positive | HS (105) | no | 0.28 | 0.13 | 1.33 | 1.13 | [1.04; 1.70] | .025* | | | | |
| ADNI-3T | Aβ-positive | LP (75) | no | 0.17 | 0.18 | 1.19 | 1.20 | [0.83; 1.70] | .349 | | | | |
| ADNI-3T | Aβ-positive | LP (75) | no | 0.17 | 0.18 | 1.19 | 1.20 | [0.83; 1.70] | .349 | | | | |
| DELCODE-3T | Whole sample | HS (234) | no | 0.29 | 0.10 | 1.33 | 1.10 | [1.10; 1.61] | .003** | | | | |
| DELCODE-3T | Whole sample | HS (234) | no | 0.29 | 0.10 | 1.33 | 1.10 | [1.10; 1.61] | .003** | | | | |
| DELCODE-3T | Whole sample | LP (185) | no | 0.55 | 0.12 | 1.73 | 1.13 | [1.37; 2.19] | < .001*** | | | | |
| DELCODE-3T | Whole sample | LP (185) | no | 0.55 | 0.12 | 1.73 | 1.13 | [1.37; 2.19] | < .001*** | | | | |
| DELCODE-3T | Aβ-positive | HS (56) | no | 0.09 | 0.15 | 1.09 | 1.17 | [0.81; 1.48] | .553 | | | | |
| DELCODE-3T | Aβ-positive | HS (56) | no | 0.09 | 0.15 | 1.09 | 1.17 | [0.81; 1.48] | .553 | | | | |
| DELCODE-3T | Aβ-positive | LP (72) | no | 0.23 | 0.14 | 1.25 | 1.15 | [0.96; 1.64] | .097 | | | | |
| DELCODE-3T | Aβ-positive | LP (72) | no | 0.23 | 0.14 | 1.25 | 1.15 | [0.96; 1.64] | .097 | | | | |
| NACC-3T | Whole sample | HS (573) | no | 0.25 | 0.06 | 1.28 | 1.06 | [1.14; 1.45] | < .001*** | | | | |
| NACC-3T | Whole sample | HS (573) | no | 0.25 | 0.06 | 1.28 | 1.06 | [1.14; 1.45] | < .001*** | | | | |
| NACC-3T | Whole sample | LP (492) | no | 0.40 | 0.08 | 1.49 | 1.09 | [1.26; 1.75] | < .001*** | | | | |
| NACC-3T | Whole sample | LP (492) | no | 0.40 | 0.08 | 1.49 | 1.09 | [1.26; 1.75] | < .001*** | | | | |
| NACC-3T | Aβ-positive | HS (46) | no | 0.39 | 0.20 | 1.47 | 1.22 | [0.99; 2.18] | .055 | | | | |
| NACC-3T | Aβ-positive | HS (46) | no | 0.39 | 0.20 | 1.47 | 1.22 | [0.99; 2.18] | .055 | | | | |
| NACC-3T | Aβ-positive | LP (47) | no | 1.47 | 0.87 | 4.34 | 2.39 | [0.79; 23.89] | .091 | | | | |
| NACC-3T | Aβ-positive | LP (47) | no | 1.47 | 0.87 | 4.34 | 2.39 | [0.79; 23.89] | .091 | | | | |
| ADNI–1.5T | Whole sample | HS (121) | no | 0.35 | 0.10 | 1.41 | 1.11 | [1.15; 1.73] | < .001*** | | | | |
| ADNI–1.5T | Whole sample | HS (121) | no | 0.35 | 0.10 | 1.41 | 1.11 | [1.15; 1.73] | < .001*** | | | | |
| ADNI–1.5T | Whole sample | LP (115) | no | 1.09 | 0.49 | 2.97 | 1.62 | [1.15; 7.68] | .025* | | | | |
| ADNI–1.5T | Whole sample | LP (115) | no | 1.09 | 0.49 | 2.97 | 1.62 | [1.15; 7.68] | .025* | | | | |
| ADNI–1.5T | Aβ-positive | HS (35) | no | 0.37 | 0.23 | 1.45 | 1.25 | [0.93; 2.25] | .101 | | | | |
| ADNI–1.5T | Aβ-positive | HS (35) | no | 0.37 | 0.23 | 1.45 | 1.25 | [0.93; 2.25] | .101 | | | | |
| ADNI–1.5T | Aβ-positive | LP (49) | no | 0.44 | 0.50 | 1.55 | 1.65 | [0.59; 4.12] | .376 | | | | |
| ADNI–1.5T | Aβ-positive | LP (49) | no | 0.44 | 0.50 | 1.55 | 1.65 | [0.59; 4.12] | .376 | | | | |
| ARWIBO–1.5T | Whole sample | HS (34) | no | 0.31 | 0.28 | 1.36 | 1.32 | [0.79; 2.35] | .269 | | | | |
| ARWIBO–1.5T | Whole sample | HS (34) | no | 0.31 | 0.28 | 1.36 | 1.32 | [0.79; 2.35] | .269 | | | | |
| ARWIBO–1.5T | Whole sample | LP (26) | no | –0.01 | 0.21 | 0.99 | 1.24 | [0.65; 1.49] | .945 | | | | |
| ARWIBO–1.5T | Whole sample | LP (26) | no | –0.01 | 0.21 | 0.99 | 1.24 | [0.65; 1.49] | .945 | | | | |
| NACC–1.5T | Whole sample | HS (114) | no | –0.01 | 0.04 | 0.99 | 1.04 | [0.91; 1.08] | .816 | | | | |
| NACC–1.5T | Whole sample | HS (114) | no | –0.01 | 0.04 | 0.99 | 1.04 | [0.91; 1.08] | .816 | | | | |
| NACC–1.5T | Whole sample | LP (51) | no | 0.73 | 0.39 | 2.07 | 1.47 | [0.97; 4.42] | .061 | | | | |
| NACC–1.5T | Whole sample | LP (51) | no | 0.73 | 0.39 | 2.07 | 1.47 | [0.97; 4.42] | .061 | | | | |
| **Outcome: DAT *versus* CU/MCI diagnosis** | | | | | | | | | | | | | |
| Pooled | Whole sample | HS (3232) | | 0.46 | 0.08 | 1.58 | 1.09 | [1.34; 1.86] | | 0.07 | 94.15 | < .001*** | < .001*** |

| **Sample** | **Sample** | **Subtype (*n*)** | **Used Firth's penalization** | **log(*OR*)** | ***SE*** | ***OR*** | ***SE*** | **95% C.I.** | ***p*** | **$\tau^2$** | **$I^2$** | **Pooled $p_{raw}$** | **Pooled $p_{FDR}$** |
|---|---|---|---|---|---|---|---|---|---|---|---|---|---|
| Pooled | Whole sample | LP (3372) | | 0.33 | 0.04 | 1.39 | 1.04 | [1.29; 1.50] | | 0.01 | 71.35 | < .001*** | < .001*** |
| Pooled | Aβ-positive | HS (668) | | 0.42 | 0.06 | 1.53 | 1.06 | [1.36; 1.71] | | 0.01 | 40.77 | < .001*** | < .001*** |
| Pooled | Aβ-positive | LP (924) | | 0.27 | 0.04 | 1.31 | 1.04 | [1.21; 1.42] | | 0.00 | 34.12 | < .001*** | < .001*** |
| ADNI-3T | Whole sample | HS (289) | no | 0.60 | 0.09 | 1.82 | 1.09 | [1.53; 2.16] | < .001*** | | | | |
| ADNI-3T | Whole sample | HS (289) | no | 0.60 | 0.09 | 1.82 | 1.09 | [1.53; 2.16] | < .001*** | | | | |
| ADNI-3T | Whole sample | LP (225) | no | 0.20 | 0.06 | 1.22 | 1.06 | [1.09; 1.37] | < .001*** | | | | |
| ADNI-3T | Whole sample | LP (225) | no | 0.20 | 0.06 | 1.22 | 1.06 | [1.09; 1.37] | < .001*** | | | | |
| ADNI-3T | Aβ-positive | HS (143) | no | 0.49 | 0.10 | 1.63 | 1.10 | [1.35; 1.96] | < .001*** | | | | |
| ADNI-3T | Aβ-positive | HS (143) | no | 0.49 | 0.10 | 1.63 | 1.10 | [1.35; 1.96] | < .001*** | | | | |
| ADNI-3T | Aβ-positive | LP (145) | no | 0.16 | 0.07 | 1.17 | 1.07 | [1.03; 1.34] | .017* | | | | |
| ADNI-3T | Aβ-positive | LP (145) | no | 0.16 | 0.07 | 1.17 | 1.07 | [1.03; 1.34] | .017* | | | | |
| DELCODE-3T | Whole sample | HS (254) | no | 0.90 | 0.16 | 2.45 | 1.17 | [1.80; 3.33] | < .001*** | | | | |
| DELCODE-3T | Whole sample | HS (254) | no | 0.90 | 0.16 | 2.45 | 1.17 | [1.80; 3.33] | < .001*** | | | | |
| DELCODE-3T | Whole sample | LP (265) | no | 0.41 | 0.07 | 1.51 | 1.07 | [1.31; 1.73] | < .001*** | | | | |
| DELCODE-3T | Whole sample | LP (265) | no | 0.41 | 0.07 | 1.51 | 1.07 | [1.31; 1.73] | < .001*** | | | | |
| DELCODE-3T | Aβ-positive | HS (71) | no | 0.91 | 0.25 | 2.48 | 1.29 | [1.51; 4.08] | < .001*** | | | | |
| DELCODE-3T | Aβ-positive | HS (71) | no | 0.91 | 0.25 | 2.48 | 1.29 | [1.51; 4.08] | < .001*** | | | | |
| DELCODE-3T | Aβ-positive | LP (132) | no | 0.39 | 0.10 | 1.48 | 1.10 | [1.22; 1.80] | < .001*** | | | | |
| DELCODE-3T | Aβ-positive | LP (132) | no | 0.39 | 0.10 | 1.48 | 1.10 | [1.22; 1.80] | < .001*** | | | | |
| NACC-3T | Whole sample | HS (698) | no | 0.68 | 0.06 | 1.97 | 1.06 | [1.76; 2.21] | < .001*** | | | | |
| NACC-3T | Whole sample | HS (698) | no | 0.68 | 0.06 | 1.97 | 1.06 | [1.76; 2.21] | < .001*** | | | | |
| NACC-3T | Whole sample | LP (784) | no | 0.51 | 0.05 | 1.67 | 1.05 | [1.52; 1.83] | < .001*** | | | | |
| NACC-3T | Whole sample | LP (784) | no | 0.51 | 0.05 | 1.67 | 1.05 | [1.52; 1.83] | < .001*** | | | | |
| NACC-3T | Aβ-positive | HS (64) | no | 0.37 | 0.14 | 1.45 | 1.15 | [1.10; 1.92] | .008** | | | | |
| NACC-3T | Aβ-positive | HS (64) | no | 0.37 | 0.14 | 1.45 | 1.15 | [1.10; 1.92] | .008** | | | | |
| NACC-3T | Aβ-positive | LP (98) | no | 0.24 | 0.13 | 1.28 | 1.14 | [0.99; 1.64] | .059 | | | | |
| NACC-3T | Aβ-positive | LP (98) | no | 0.24 | 0.13 | 1.28 | 1.14 | [0.99; 1.64] | .059 | | | | |
| ADNI–1.5T | Whole sample | HS (170) | no | 0.25 | 0.06 | 1.29 | 1.06 | [1.16; 1.44] | < .001*** | | | | |
| ADNI–1.5T | Whole sample | HS (170) | no | 0.25 | 0.06 | 1.29 | 1.06 | [1.16; 1.44] | < .001*** | | | | |
| ADNI–1.5T | Whole sample | LP (200) | no | 0.33 | 0.07 | 1.40 | 1.07 | [1.22; 1.60] | < .001*** | | | | |
| ADNI–1.5T | Whole sample | LP (200) | no | 0.33 | 0.07 | 1.40 | 1.07 | [1.22; 1.60] | < .001*** | | | | |
| ADNI–1.5T | Aβ-positive | HS (56) | no | 0.26 | 0.10 | 1.30 | 1.10 | [1.07; 1.57] | .007** | | | | |
| ADNI–1.5T | Aβ-positive | HS (56) | no | 0.26 | 0.10 | 1.30 | 1.10 | [1.07; 1.57] | .007** | | | | |
| ADNI–1.5T | Aβ-positive | LP (87) | no | 0.35 | 0.11 | 1.42 | 1.12 | [1.14; 1.78] | .002** | | | | |
| ADNI–1.5T | Aβ-positive | LP (87) | no | 0.35 | 0.11 | 1.42 | 1.12 | [1.14; 1.78] | .002** | | | | |
| ARWIBO–1.5T | Whole sample | HS (38) | no | 0.23 | 0.23 | 1.26 | 1.25 | [0.81; 1.96] | .312 | | | | |
| ARWIBO–1.5T | Whole sample | HS (38) | no | 0.23 | 0.23 | 1.26 | 1.25 | [0.81; 1.96] | .312 | | | | |
| ARWIBO–1.5T | Whole sample | LP (54) | no | 0.24 | 0.14 | 1.27 | 1.15 | [0.97; 1.66] | .085 | | | | |
| ARWIBO–1.5T | Whole sample | LP (54) | no | 0.24 | 0.14 | 1.27 | 1.15 | [0.97; 1.66] | .085 | | | | |
| NACC–1.5T | Whole sample | HS (167) | no | 0.12 | 0.04 | 1.13 | 1.04 | [1.05; 1.22] | .002** | | | | |
| NACC–1.5T | Whole sample | HS (167) | no | 0.12 | 0.04 | 1.13 | 1.04 | [1.05; 1.22] | .002** | | | | |
| NACC–1.5T | Whole sample | LP (158) | no | 0.21 | 0.09 | 1.23 | 1.10 | [1.03; 1.47] | .024* | | | | |
| NACC–1.5T | Whole sample | LP (158) | no | 0.21 | 0.09 | 1.23 | 1.10 | [1.03; 1.47] | .024* | | | | |

*$p < .05$. **$p < .01$. ***$p < .001$. Abbreviations: HS, hippocampal-sparing. LP, limbic-predominant.

**Supplementary Table 6 Summary statistics of atrophy subtype and stage effects on regional CenTauR scores, extracted from linear regression models.**

| ROI | Predictor | Estimate | 95% C.I. | *SE* | *t* | *p* |
|---|---|---|---|---|---|---|
| Frontal | Atrophy subtype (LP) | 0.44 | [–5.83; 6.71] | 3.19 | 0.14 | .891 |
| | Atrophy stage (spline 1) | 26.87 | [9.28; 44.46] | 8.95 | 3.00 | .003** |
| | Atrophy stage (spline 2) | 71.40 | [44.82; 97.98] | 13.53 | 5.28 | < .001*** |
| | Atrophy subtype (LP) × atrophy stage (spline 1) | 90.73 | [54.64; 126.83] | 18.37 | 4.94 | < .001*** |
| | Atrophy subtype (LP) × atrophy stage (spline 2) | 122.86 | [49.93; 195.78] | 37.10 | 3.31 | .001** |
| Mesial temporal | Atrophy subtype (LP) | 3.06 | [–3.00; 9.12] | 3.08 | 0.99 | .321 |
| | Atrophy stage (spline 1) | 23.51 | [6.52; 40.51] | 8.65 | 2.72 | .007** |
| | Atrophy stage (spline 2) | 34.42 | [8.73; 60.10] | 13.07 | 2.63 | .009** |
| | Atrophy subtype (LP) × atrophy stage (spline 1) | 58.90 | [24.03; 93.77] | 17.74 | 3.32 | < .001*** |
| | Atrophy subtype (LP) × atrophy stage (spline 2) | –8.20 | [–78.65; 62.25] | 35.85 | –0.23 | .819 |
| Meta temporal | Atrophy subtype (LP) | 1.89 | [–3.87; 7.66] | 2.93 | 0.65 | .519 |
| | Atrophy stage (spline 1) | 29.05 | [12.89; 45.21] | 8.22 | 3.53 | < .001*** |
| | Atrophy stage (spline 2) | 47.86 | [23.44; 72.29] | 12.43 | 3.85 | < .001*** |
| | Atrophy subtype (LP) × atrophy stage (spline 1) | 77.21 | [44.05; 110.38] | 16.88 | 4.58 | < .001*** |
| | Atrophy subtype (LP) × atrophy stage (spline 2) | 85.75 | [18.74; 152.76] | 34.09 | 2.52 | .012* |
| Temporo–parietal | Atrophy subtype (LP) | 1.76 | [–3.42; 6.94] | 2.63 | 0.67 | .505 |
| | Atrophy stage (spline 1) | 22.54 | [8.02; 37.06] | 7.39 | 3.05 | .002** |
| | Atrophy stage (spline 2) | 49.05 | [27.11; 70.99] | 11.16 | 4.39 | < .001*** |
| | Atrophy subtype (LP) × atrophy stage (spline 1) | 94.39 | [64.59; 124.18] | 15.16 | 6.23 | < .001*** |
| | Atrophy subtype (LP) × atrophy stage (spline 2) | 142.49 | [82.30; 202.69] | 30.63 | 4.65 | < .001*** |
| Universal | Atrophy subtype (LP) | 1.36 | [–3.87; 6.59] | 2.66 | 0.51 | .609 |
| | Atrophy stage (spline 1) | 21.79 | [7.13; 36.45] | 7.46 | 2.92 | .004** |
| | Atrophy stage (spline 2) | 46.93 | [24.78; 69.08] | 11.27 | 4.16 | < .001*** |
| | Atrophy subtype (LP) × atrophy stage (spline 1) | 92.92 | [62.84; 123.01] | 15.31 | 6.07 | < .001*** |
| | Atrophy subtype (LP) × atrophy stage (spline 2) | 133.61 | [72.84; 194.38] | 30.92 | 4.32 | < .001*** |

Models additionally included age, sex, APOE ε4 status, diagnosis, treatment, and PET–MRI time interval as covariates (not shown). *$p < .05$. **$p < .01$. ***$p < .001$. Abbreviations: LP, limbic-predominant.

**Supplementary Table 7 Cohort-wise summary statistics of atrophy subtype and stage effects on regional CenTauR scores while controlling for plasma p-tau biomarkers, extracted from linear regression models.**

| Cohort | ROI | Predictor | Estimate | 95% C.I. | *SE* | *t* | *p* |
|---|---|---|---|---|---|---|---|
| A4 | Frontal | Atrophy subtype (LP) | −1.16 | [−6.73; 4.41] | 2.82 | −0.41 | .681 |
| | | Atrophy stage | −0.57 | [−2.37; 1.22] | 0.91 | −0.63 | .530 |
| | | Atrophy subtype (LP) × atrophy stage | 2.92 | [−1.37; 7.21] | 2.17 | 1.34 | .181 |
| | Mesial temporal | Atrophy subtype (LP) | 4.90 | [−1.25; 11.05] | 3.12 | 1.57 | .118 |
| | | Atrophy stage | 0.47 | [−1.52; 2.45] | 1.00 | 0.46 | .643 |
| | | Atrophy subtype (LP) × atrophy stage | 4.73 | [0.00; 9.47] | 2.40 | 1.97 | .050 |
| | Meta temporal | Atrophy subtype (LP) | 1.51 | [−2.71; 5.73] | 2.14 | 0.70 | .482 |
| | | Atrophy stage | 0.05 | [−1.31; 1.41] | 0.69 | 0.08 | .939 |
| | | Atrophy subtype (LP) × atrophy stage | 2.09 | [−1.16; 5.34] | 1.65 | 1.27 | .205 |
| | Temporo–parietal | Atrophy subtype (LP) | 0.41 | [−3.45; 4.28] | 1.96 | 0.21 | .832 |
| | | Atrophy stage | −0.53 | [−1.77; 0.72] | 0.63 | −0.83 | .405 |
| | | Atrophy subtype (LP) × atrophy stage | 1.57 | [−1.40; 4.55] | 1.51 | 1.04 | .298 |
| | Universal | Atrophy subtype (LP) | 0.37 | [−3.60; 4.35] | 2.01 | 0.19 | .853 |
| | | Atrophy stage | −0.59 | [−1.87; 0.69] | 0.65 | −0.91 | .365 |
| | | Atrophy subtype (LP) × atrophy stage | 1.77 | [−1.29; 4.84] | 1.55 | 1.14 | .254 |
| ADNI | Frontal | Atrophy subtype (LP) | 3.21 | [−18.70; 25.11] | 11.02 | 0.29 | .772 |
| | | Atrophy stage (spline 1) | 30.44 | [−25.64; 86.53] | 28.21 | 1.08 | .283 |
| | | Atrophy stage (spline 2) | 61.74 | [5.83; 117.64] | 28.12 | 2.20 | .031* |
| | | Atrophy subtype (LP) × atrophy stage (spline 1) | 93.75 | [5.46; 182.04] | 44.41 | 2.11 | .038* |
| | | Atrophy subtype (LP) × atrophy stage (spline 2) | 127.20 | [−13.54; 267.94] | 70.79 | 1.80 | .076 |
| | Mesial temporal | Atrophy subtype (LP) | 4.05 | [−13.36; 21.45] | 8.75 | 0.46 | .645 |
| | | Atrophy stage (spline 1) | 20.85 | [−23.72; 65.41] | 22.41 | 0.93 | .355 |
| | | Atrophy stage (spline 2) | 35.46 | [−8.96; 79.88] | 22.34 | 1.59 | .116 |
| | | Atrophy subtype (LP) × atrophy stage (spline 1) | 32.55 | [−37.61; 102.70] | 35.28 | 0.92 | .359 |
| | Meta temporal | Atrophy subtype (LP) × atrophy stage (spline 2) | −64.15 | [−175.98; 47.68] | 56.25 | −1.14 | .257 |
| | | Atrophy stage (spline 1) | 3.13 | [−12.75; 19.00] | 7.99 | 0.39 | .696 |
| | | Atrophy stage (spline 2) | 4.04 | [0.03; 8.05] | 2.02 | 2.00 | .049* |
| | | Atrophy subtype (LP) × atrophy stage (spline 1) | 3.31 | [−3.58; 10.20] | 3.47 | 0.95 | .343 |
| | Temporo–parietal | Atrophy subtype (LP) × atrophy stage (spline 2) | −1.41 | [−18.04; 15.21] | 8.36 | −0.17 | .866 |
| | | Atrophy stage (spline 1) | 13.02 | [−29.56; 55.59] | 21.41 | 0.61 | .545 |
| | | Atrophy stage (spline 2) | 53.39 | [10.95; 95.82] | 21.34 | 2.50 | .014* |
| | | Atrophy subtype (LP) × atrophy stage (spline 1) | 105.49 | [38.47; 172.51] | 33.71 | 3.13 | .002** |
| | | Atrophy subtype (LP) × atrophy stage (spline 2) | 137.47 | [30.63; 244.30] | 53.73 | 2.56 | .012* |
| | Universal | Atrophy subtype (LP) | −0.44 | [−17.08; 16.20] | 8.37 | −0.05 | .958 |
| | | Atrophy stage (spline 1) | 16.10 | [−26.51; 58.70] | 21.43 | 0.75 | .455 |
| | | Atrophy stage (spline 2) | 50.49 | [8.02; 92.96] | 21.36 | 2.36 | .020* |
| | | Atrophy subtype (LP) × atrophy stage (spline 1) | 104.51 | [37.44; 171.58] | 33.73 | 3.10 | .003** |
| | | Atrophy subtype (LP) × atrophy stage (spline 2) | 128.10 | [21.18; 235.02] | 53.77 | 2.38 | .019* |

Models additionally included age, sex, APOE ε4 status, diagnosis, treatment, PET–MRI time interval, and plasma p-tau as covariates (not shown). *$p < .05$. **$p < .01$. ***$p < .001$. Abbreviations: LP, limbic-predominant.

**Supplementary Table 8 Summary statistics of atrophy subtype and stage effects on tau burden group in two samples, extracted from the best-fit ordinal logistic regression model.**

| Sample | Predictor | Estimate | 95% C.I. | *SE* | *z* | *p* |
|---|---|---|---|---|---|---|
| Whole sample<br>ΔAIC = −12.02<br>$\Delta R^2_{\text{Nagelkerke}}$ = 0.04 | Age at MRI | −0.01 | [−0.04; 0.03] | 0.02 | −0.42 | .676 |
| | Sex (female) | 0.20 | [−0.27; 0.67] | 0.24 | 0.85 | .397 |
| | APOE ε4 carriership (carrier) | 0.19 | [−0.31; 0.70] | 0.26 | 0.73 | .466 |
| | PET–MRI interval | 0.00 | [−0.01; 0.00] | 0.00 | −0.45 | .656 |
| | Diagnosis (MCI) | 2.08 | [1.46; 2.72] | 0.32 | 6.45 | < .001*** |
| | Diagnosis (DAT) | 2.33 | [1.45; 3.23] | 0.45 | 5.16 | < .001*** |
| | Treatment (solanezumab) | −0.11 | [−0.77; 0.51] | 0.33 | −0.35 | .729 |
| | Atrophy stage | 0.21 | [0.06; 0.36] | 0.08 | 2.80 | .005** |
| | Atrophy subtype (LP) | 0.12 | [−0.47; 0.72] | 0.30 | 0.41 | .683 |
| | Atrophy subtype (LP) × atrophy stage | 0.24 | [−0.03; 0.51] | 0.14 | 1.74 | .082 |
| ATT-eligible sample<br>ΔAIC = −6.42<br>$\Delta R^2_{\text{Nagelkerke}}$ = 0.17 | Age at MRI | −0.06 | [−0.13; 0.00] | 0.03 | −1.86 | .063 |
| | Sex (female) | 0.07 | [−0.87; 1.01] | 0.48 | 0.15 | .883 |
| | APOE ε4 carriership (carrier) | 0.39 | [−0.66; 1.46] | 0.54 | 0.73 | .464 |
| | PET–MRI interval | 0.00 | [−0.01; 0.01] | 0.01 | −0.76 | .450 |
| | Diagnosis (DAT) | −0.38 | [−1.47; 0.68] | 0.54 | −0.70 | .482 |
| | Atrophy stage | 0.39 | [0.11; 0.69] | 0.15 | 2.68 | .007** |
| | Atrophy subtype (LP) | 0.32 | [−1.10; 1.77] | 0.73 | 0.44 | .662 |
| | Atrophy subtype (LP) × atrophy stage | 0.07 | [−0.36; 0.50] | 0.22 | 0.32 | .751 |

*$p < .05$. **$p < .01$. ***$p < .001$. Abbreviations: LP, limbic-predominant.

**Supplementary Table 9 Summary statistics of LMMs predicting longitudinal PACC–5 scores using baseline atrophy subtype and stage in the preclinical AD sample.**

| Predictor | Estimate | *SE* | df | *t* value | p |
|---|---|---|---|---|---|
| Intercept | −2.10 | 0.99 | 1556.14 | −2.13 | .033* |
| Years (spline 1) | 16.45 | 3.48 | 849.35 | 4.73 | < .001*** |
| Years (spline 2) | 7.56 | 2.26 | 886.37 | 3.35 | < .001*** |
| Age at MRI | 0.03 | 0.01 | 1551.65 | 2.01 | .045* |
| Sex (female) | −0.02 | 0.12 | 1501.30 | −0.19 | .853 |
| APOE ε4 carriership (carrier) | 0.01 | 0.12 | 1516.57 | 0.06 | .949 |
| Treatment (solanezumab) | 0.09 | 0.12 | 1515.32 | 0.82 | .414 |
| Years of education | −0.01 | 0.02 | 1571.44 | −0.67 | .501 |
| Version B: Martha Jackson | −0.26 | 0.04 | 5364.18 | −6.73 | < .001*** |
| Version SC: Anna Thompson | −0.82 | 0.16 | 5839.51 | −5.16 | < .001*** |
| Version SC: Robert Miller | −0.90 | 0.04 | 5399.67 | −20.38 | < .001*** |
| Baseline atrophy subtype (LP) | 0.12 | 0.14 | 1516.34 | 0.83 | .409 |
| Baseline atrophy stage | 0.08 | 0.05 | 1592.84 | 1.65 | .098 |
| Years (spline 1) × age at MRI | −0.19 | 0.05 | 848.32 | −4.26 | < .001*** |
| Years (spline 2) × age at MRI | −0.11 | 0.03 | 900.60 | −3.64 | < .001*** |
| Years (spline 1) × sex (female) | 0.65 | 0.43 | 825.07 | 1.50 | .133 |
| Years (spline 2) × sex (female) | 0.57 | 0.27 | 822.37 | 2.08 | .038* |
| Years (spline 1) × APOE ε4 carriership (carrier) | −0.94 | 0.42 | 831.23 | −2.25 | .024* |
| Years (spline 2) × APOE ε4 carriership (carrier) | −0.91 | 0.27 | 837.09 | −3.41 | < .001*** |
| Years (spline 1) × treatment (solanezumab | −0.32 | 0.41 | 828.91 | −0.80 | .426 |
| Years (spline 2) × treatment (solanezumab) | −0.27 | 0.26 | 831.26 | −1.06 | .289 |
| Years (spline 1) × years of education | −0.05 | 0.08 | 849.42 | −0.61 | .543 |
| Years (spline 2) × years of education | 0.02 | 0.05 | 884.62 | 0.41 | .681 |
| Years (spline 1) × baseline atrophy subtype (LP) | −0.32 | 0.49 | 826.25 | −0.66 | .510 |
| Years (spline 2) × baseline atrophy subtype (LP) | −0.81 | 0.31 | 840.67 | −2.61 | .009** |
| Years (spline 1) × baseline atrophy stage | −0.93 | 0.17 | 852.96 | −5.58 | < .001*** |
| Years (spline 2) × baseline atrophy stage | −0.77 | 0.11 | 944.64 | −6.97 | < .001*** |
| Baseline atrophy subtype (LP) × baseline atrophy stage | 0.14 | 0.14 | 1519.31 | 1.01 | .312 |
| Years (spline 1) × baseline atrophy subtype (LP) × baseline atrophy stage | −1.84 | 0.46 | 824.21 | −3.98 | < .001*** |
| Years (spline 2) × baseline atrophy subtype (LP) × baseline atrophy stage | −1.13 | 0.30 | 906.25 | −3.77 | < .001*** |

Formula: PACC-5 ~ ns(years, *df* = 2) × age at MRI + ns(years, *df* = 2) × sex + ns(years, *df* = 2) × APOE ε4 carriership + ns(years, *df* = 2) × treatment + ns(years, *df* = 2) × years of education + version + ns(years, *df* = 2) × baseline atrophy subtype × baseline atrophy stage + (years | participant). $R^2_{\text{marginal}}$ (REML/ML) = 0.27/0.16. *$p < .05$. **$p < .01$. ***$p < .001$. Abbreviations: LP, limbic-predominant.

**Supplementary Table 10 Summary statistics of LMMs predicting longitudinal CDR–SB scores using baseline atrophy subtype and stage in the mild symptomatic AD sample.**

| **Predictor** | **Estimate** | ***SE*** | **df** | ***t* value** | **p** |
|---|---|---|---|---|---|
| Intercept | 3.59 | 2.00 | 1739.69 | 1.79 | .074 |
| Years (spline 1) | –5.33 | 3.79 | 1416.18 | –1.41 | .160 |
| Years (spline 2) | 1.45 | 2.10 | 1422.79 | 0.69 | .489 |
| Age at MRI | –0.03 | 0.02 | 1744.53 | –1.17 | .244 |
| Sex (female) | –0.09 | 0.30 | 1749.84 | –0.32 | .751 |
| Years of education | –0.03 | 0.05 | 1708.32 | –0.67 | .504 |
| APOE ε4 carriership (carrier) | –0.41 | 0.34 | 1737.39 | –1.22 | .224 |
| Diagnosis (DAT) | 1.78 | 0.33 | 1772.42 | 5.34 | < .001*** |
| Baseline atrophy subtype (LP) | 0.30 | 0.52 | 1738.90 | 0.58 | .564 |
| Baseline atrophy stage (spline 1) | 0.28 | 1.11 | 1749.30 | 0.25 | .802 |
| Baseline atrophy stage (spline 2) | –0.92 | 1.10 | 1727.06 | –0.84 | .403 |
| Years (spline 1) × age at MRI | 0.09 | 0.04 | 1399.07 | 2.23 | .026* |
| Years (spline 2) × age at MRI | 0.01 | 0.02 | 1426.04 | 0.27 | .787 |
| Years (spline 1) × sex (female) | –0.29 | 0.56 | 1412.83 | –0.51 | .612 |
| Years (spline 2) × sex (female) | –0.64 | 0.31 | 1421.90 | –2.08 | .038* |
| Years (spline 1) × years of education | –0.03 | 0.10 | 1468.13 | –0.28 | .779 |
| Years (spline 2) × years of education | –0.09 | 0.05 | 1414.41 | –1.65 | .099 |
| Years (spline 1) × APOE ε4 carriership (carrier) | 0.57 | 0.64 | 1400.28 | 0.89 | .372 |
| Years (spline 2) × APOE ε4 carriership (carrier) | –0.39 | 0.33 | 1417.00 | –1.19 | .234 |
| Years (spline 1) × diagnosis (DAT) | 3.21 | 0.62 | 1404.48 | 5.20 | < .001*** |
| Years (spline 2) × diagnosis (DAT) | 1.44 | 0.52 | 1473.55 | 2.78 | .005** |
| Years (spline 1) × baseline atrophy subtype (LP) | 0.62 | 0.98 | 1397.12 | 0.63 | .526 |
| Years (spline 2) × baseline atrophy subtype (LP) | 0.88 | 0.45 | 1400.09 | 1.97 | .049* |
| Years (spline 1) × baseline atrophy stage (spline 1) | 9.75 | 2.09 | 1400.86 | 4.66 | < .001*** |
| Years (spline 2) × baseline atrophy stage (spline 1) | 8.20 | 1.10 | 1412.18 | 7.45 | < .001*** |
| Years (spline 1) × baseline atrophy stage (spline 2) | 6.36 | 2.06 | 1412.38 | 3.09 | .002** |
| Years (spline 2) × baseline atrophy stage (spline 2) | 1.45 | 1.41 | 1432.67 | 1.03 | .302 |
| Baseline atrophy subtype (LP) × baseline atrophy stage (spline 1) | 1.08 | 1.40 | 1742.45 | 0.77 | .439 |
| Baseline atrophy subtype (LP) × baseline atrophy stage (spline 2) | 2.26 | 1.77 | 1743.41 | 1.28 | .202 |
| Years (spline 1) × baseline atrophy subtype (LP) × baseline atrophy stage (spline 1) | –1.44 | 2.65 | 1419.44 | –0.55 | .586 |
| Years (spline 2) × baseline atrophy subtype (LP) × baseline atrophy stage (spline 1) | 3.00 | 1.52 | 1421.61 | 1.98 | .048* |
| Years (spline 1) × baseline atrophy subtype (LP) × baseline atrophy stage (spline 2) | –3.69 | 3.34 | 1404.66 | –1.11 | .269 |
| Years (spline 2) × baseline atrophy subtype (LP) × baseline atrophy stage (spline 2) | 1.61 | 2.52 | 1441.72 | 0.64 | .525 |

Formula: CDR-SB ~ ns(years, *df* = 2) × age at MRI + ns(years, *df* = 2) × sex + ns(years, *df* = 2) × years of education + ns(years, *df* = 2) × APOE ε4 carriership + ns(years, *df* = 2) × baseline diagnosis + ns(years, *df* = 2) × baseline atrophy subtype × ns(baseline atrophy stage, knot = 3) + (1 | participant). $R^2_{\text{marginal}}$ (REML/ML) = 0.48/0.49. *$p < .05$. **$p < .01$. ***$p < .001$. Abbreviations: LP, limbic-predominant.

**Supplementary Table 11 Summary statistics of LMMs predicting longitudinal PACC–5 scores using baseline atrophy subtype and stage in the preclinical AD sample, controlling for p-tau$_{217}$.**

| Predictor | Estimate | *SE* | df | *t* value | p |
|---|---|---|---|---|---|
| Intercept | –2.33 | 1.06 | 1350.95 | –2.21 | .028* |
| Years (spline 1) | 14.29 | 3.59 | 755.73 | 3.99 | < .001*** |
| Years (spline 2) | 3.76 | 2.34 | 784.22 | 1.60 | .109 |
| Age at MRI | 0.03 | 0.01 | 1345.13 | 1.89 | .059 |
| Sex (female) | 0.07 | 0.13 | 1298.23 | 0.51 | .612 |
| APOE ε4 carriership (carrier) | –0.03 | 0.13 | 1310.95 | –0.22 | .823 |
| Treatment (solanezumab) | 0.07 | 0.13 | 1308.15 | 0.56 | .574 |
| Years of education | –0.01 | 0.02 | 1355.58 | –0.28 | .776 |
| Version B: Martha Jackson | –0.25 | 0.04 | 4627.91 | –5.84 | < .001*** |
| Version SC: Anna Thompson | –0.76 | 0.17 | 5037.48 | –4.50 | < .001*** |
| Version SC: Robert Miller | –0.93 | 0.05 | 4657.16 | –19.35 | < .001*** |
| Baseline atrophy subtype (LP) | 0.14 | 0.15 | 1312.20 | 0.94 | .349 |
| Baseline atrophy stage | 0.08 | 0.05 | 1384.64 | 1.53 | .126 |
| p-tau$_{217}$ | 0.35 | 0.42 | 1288.57 | 0.84 | .404 |
| Years (spline 1) × age at MRI | –0.13 | 0.05 | 753.81 | –2.77 | .006** |
| Years (spline 2) × age at MRI | –0.03 | 0.03 | 798.83 | –0.88 | .377 |
| Years (spline 1) × sex (female) | 0.26 | 0.45 | 729.13 | 0.57 | .566 |
| Years (spline 2) × sex (female) | 0.34 | 0.28 | 718.80 | 1.21 | .228 |
| Years (spline 1) × APOE ε4 carriership (carrier) | –0.75 | 0.44 | 734.50 | –1.71 | .087 |
| Years (spline 2) × APOE ε4 carriership (carrier) | –0.68 | 0.28 | 730.97 | –2.45 | .015* |
| Years (spline 1) × treatment (solanezumab | 0.06 | 0.42 | 731.65 | 0.15 | .878 |
| Years (spline 2) × treatment (solanezumab) | 0.13 | 0.27 | 724.27 | 0.50 | .620 |
| Years (spline 1) × years of education | –0.05 | 0.08 | 748.81 | –0.57 | .566 |
| Years (spline 2) × years of education | 0.03 | 0.05 | 781.30 | 0.58 | .561 |
| Years (spline 1) × baseline atrophy subtype (LP) | –0.65 | 0.51 | 730.78 | –1.27 | .205 |
| Years (spline 2) × baseline atrophy subtype (LP) | –0.98 | 0.33 | 738.26 | –3.01 | .003** |
| Years (spline 1) × p-tau$_{217}$ | –0.75 | 0.17 | 753.31 | –4.39 | < .001*** |
| Years (spline 2) × p-tau$_{217}$ | –0.53 | 0.11 | 836.62 | –4.62 | < .001*** |
| Years (spline 1) × baseline atrophy stage | 0.15 | 0.14 | 1315.87 | 1.05 | .294 |
| Years (spline 2) × baseline atrophy stage | –9.29 | 1.39 | 727.14 | –6.69 | < .001*** |
| Baseline atrophy subtype (LP) × baseline atrophy stage | –8.77 | 0.90 | 777.61 | –9.75 | < .001*** |
| Years (spline 1) × baseline atrophy subtype (LP) × baseline atrophy stage | –1.82 | 0.45 | 730.46 | –4.02 | < .001*** |
| Years (spline 2) × baseline atrophy subtype (LP) × baseline atrophy stage | –1.21 | 0.30 | 800.55 | –4.11 | < .001*** |

*N* = 601. Formula: PACC-5 ~ ns(years, *df* = 2) × age at MRI + ns(years, *df* = 2) × sex + ns(years, *df* = 2) × APOE ε4 carriership + ns(years, *df* = 2) × treatment + ns(years, *df* = 2) × years of education + ns(years, *df* = 2) × baseline p-tau$_{217}$ + version + ns(years, *df* = 2) × baseline atrophy subtype × baseline atrophy stage + (years | participant). $R^2_{\text{marginal}}$ (REML/ML) = 0.34/0.22. **p* < .05. ***p* < .01. ****p* < .001. Abbreviations: LP, limbic-predominant.

**Supplementary Table 12 Summary statistics of LMMs predicting longitudinal CDR–SB scores using baseline atrophy subtype and stage in the mild symptomatic AD sample, controlling for p-tau$_{217}$/Aβ$_{42}$ ratios.**

| Predictor | Estimate | *SE* | df | *t* value | p |
|---|---|---|---|---|---|
| Intercept | 3.04 | 3.10 | 98.36 | 0.98 | .329 |
| Years | 1.04 | 1.23 | 223.34 | 0.85 | .399 |
| Age at MRI | 0.01 | 0.03 | 97.02 | 0.22 | .826 |
| Sex (female) | –0.46 | 0.48 | 100.93 | –0.95 | .346 |
| Years of education | –0.07 | 0.09 | 94.98 | –0.77 | .446 |
| APOE ε4 carriership (carrier) | –0.58 | 0.54 | 97.56 | –1.08 | .281 |
| Diagnosis (DAT) | 2.34 | 0.51 | 97.17 | 4.55 | < .001*** |
| Baseline atrophy subtype (LP) | –0.07 | 0.72 | 96.79 | –0.09 | .925 |
| Baseline atrophy stage | –0.06 | 0.11 | 90.42 | –0.55 | .587 |
| p-tau$_{217}$/Aβ$_{42}$ | 0.24 | 0.50 | 87.14 | 0.49 | .623 |
| Years × age at MRI | 0.00 | 0.01 | 222.01 | –0.36 | .716 |
| Years × sex (female) | –0.29 | 0.21 | 223.15 | –1.40 | .163 |
| Years × years of education | –0.01 | 0.03 | 219.08 | –0.42 | .678 |
| Years × APOE ε4 carriership (carrier) | –0.14 | 0.23 | 225.28 | –0.61 | .544 |
| Years × diagnosis (DAT) | 0.87 | 0.22 | 225.14 | 3.91 | < .001*** |
| Years × baseline atrophy subtype (LP) | 0.35 | 0.27 | 216.21 | 1.29 | .198 |
| Years × baseline atrophy stage | 0.14 | 0.04 | 219.22 | 3.57 | < .001*** |
| Baseline atrophy subtype (LP) × baseline atrophy stage | 0.14 | 0.14 | 91.41 | 1.04 | .303 |
| Years × p-tau$_{217}$/Aβ$_{42}$ | –0.17 | 0.17 | 215.04 | –0.96 | .339 |
| Years × baseline atrophy subtype (LP) × baseline atrophy stage | 0.04 | 0.07 | 226.31 | 0.54 | .589 |

*N* = 80. Formula: CDR-SB ~ years × age at MRI + years × sex + years × years of education + years × APOE ε4 carriership + years × diagnosis + years × p-tau$_{217}$/Aβ$_{42}$ + years × baseline atrophy subtype × baseline atrophy stage + (1 | participant). $R^2_{\text{marginal}}$ (REML/ML) = 0.48/0.51. **p* < .05. ***p* < .01. ****p* < .001. Abbreviations: LP, limbic-predominant.

**Supplementary Table 13 Summary statistics of LMMs predicting longitudinal CDR–SB scores using baseline MTA scores in the mild symptomatic AD sample with available MTA scores.**

| **Predictor** | **Estimate** | ***SE*** | **df** | ***t* value** | **p** |
|---|---|---|---|---|---|
| Intercept | –1.76 | 5.59 | 30.58 | –0.31 | .755 |
| Years | –2.27 | 1.79 | 55.81 | –1.27 | .210 |
| Years of education | –0.05 | 0.15 | 31.82 | –0.34 | .735 |
| Age at MRI | 0.04 | 0.06 | 30.90 | 0.67 | .510 |
| Sex (female) | 0.39 | 0.84 | 32.13 | 0.47 | .644 |
| APOE ε4 carriership (carrier) | 0.44 | 0.99 | 31.86 | 0.44 | .661 |
| MTA score | 0.60 | 0.63 | 32.06 | 0.95 | .348 |
| Years × years of education | 0.01 | 0.05 | 54.59 | 0.15 | .880 |
| Years × age at MRI | 0.02 | 0.02 | 56.04 | 0.86 | .392 |
| Years × sex (female) | 0.41 | 0.29 | 55.29 | 1.43 | .159 |
| Years × APOE ε4 carriership (carrier) | 0.76 | 0.34 | 55.57 | 2.27 | .027* |
| Years × MTA score | 0.56 | 0.21 | 54.86 | 2.72 | .009** |

*N* = 29. Formula: CDR-SB ~ years × age at MRI + years × sex + years × years of education + years × APOE ε4 carriership + years × baseline diagnosis + years × baseline MTA score + (1 | participant). $R^2_{\text{marginal}}$ (REML/ML) = 0.26/0.30. **p* < .05. ***p* < .01. ****p* < .001. Abbreviations: LP, limbic-predominant.

**Supplementary Table 14 Summary statistics of LMMs predicting longitudinal CDR–SB scores using baseline atrophy subtype and stage in the mild symptomatic AD sample with available MTA scores.**

| **Predictor** | **Estimate** | ***SE*** | **df** | ***t* value** | **p** |
|---|---|---|---|---|---|
| Intercept | 0.21 | 5.62 | 25.39 | 0.04 | .970 |
| Years | –1.78 | 1.84 | 51.02 | –0.97 | .339 |
| Years of education | 0.01 | 0.16 | 26.95 | 0.08 | .933 |
| Age at MRI | 0.00 | 0.07 | 24.47 | 0.07 | .945 |
| Sex (female) | –0.18 | 0.82 | 26.50 | –0.21 | .832 |
| APOE ε4 carriership (carrier) | –0.03 | 1.02 | 26.86 | –0.03 | .974 |
| Baseline atrophy subtype (LP) | 0.15 | 1.34 | 25.54 | 0.11 | .912 |
| Baseline atrophy stage (spline 1) | 2.36 | 2.91 | 25.19 | 0.81 | .425 |
| Baseline atrophy stage (spline 2) | –1.10 | 1.84 | 26.93 | –0.60 | .554 |
| Years × years of education | 0.05 | 0.05 | 49.92 | 1.00 | .325 |
| Years × age at MRI | 0.01 | 0.02 | 50.75 | 0.42 | .676 |
| Years × sex (female) | 0.10 | 0.28 | 50.89 | 0.35 | .726 |
| Years × APOE ε4 carriership (carrier) | 0.13 | 0.35 | 50.52 | 0.37 | .714 |
| Years × baseline atrophy subtype (LP) | 0.40 | 0.41 | 49.76 | 0.98 | .331 |
| Years × baseline atrophy stage (spline 1) | 2.77 | 0.87 | 49.74 | 3.17 | .003** |
| Years × baseline atrophy stage (spline 2) | –1.57 | 0.64 | 49.77 | –2.44 | .018* |
| Baseline atrophy subtype (LP) × baseline atrophy stage (spline 1) | 0.88 | 3.57 | 25.26 | 0.25 | .808 |
| Baseline atrophy subtype (LP) × baseline atrophy stage (spline 2) | 0.45 | 2.97 | 28.19 | 0.15 | .881 |
| Years × baseline atrophy subtype (LP) × baseline atrophy stage (spline 1) | –1.52 | 1.12 | 49.81 | –1.36 | .179 |
| Years × baseline atrophy subtype (LP) × baseline atrophy stage (spline 2) | 1.16 | 1.08 | 49.31 | 1.08 | .288 |

*N* = 29. Formula: CDR-SB ~ years × age at MRI + years × sex + years × years of education + years × APOE ε4 carriership + years × baseline diagnosis + years × baseline atrophy subtype × ns(baseline atrophy stage, knot = 3) + (1 | participant). $R^2_{\text{marginal}}$ (REML/ML) = 0.38/0.47. **p* < .05. ***p* < .01. ****p* < .001. Abbreviations: LP, limbic-predominant.
